# Nationwide Functional Profiling Enables Clinical Reclassification of *KCNH2* Variants in the French LQT2 Population

**DOI:** 10.64898/2026.11.10.26367605

**Authors:** Barbara Ribeiro de Oliveira, Malak Alameh, Sylvain Feliciangeli, Jérôme Montnach, Brieuc Quemeneur, Thomas Gabillon, Nawel Aoujaj, Aline Rio, Floriane Bibault, Elea Lecointe, Ngone Ba, Elouan Voisin, Isabelle Baró, Richard Redon, Aurélie Thollet, Jean-Jacques Schott, Alban Gaignard, Isabelle Denjoy, Vincent Probst, Rupamanjari Majumder, Florian Lesage, Gildas Loussouarn, Michel De Waard

## Abstract

**Background:** The increasing use of genetic testing has identified thousands of *KCNH2* variants associated with long QT syndrome type 2 (LQT2), yet most remain variants of uncertain significance (VUS). Translating functional abnormalities into clinically actionable evidence remains a major challenge. We established a nationwide functional genomics strategy to systematically characterize *KCNH2* variants identified in the French LQT2 population and improve their clinical interpretation.

**Methods:** We characterized 303 *KCNH2* variants, including 201 VUS, using standardized high-throughput automated patch-clamp electrophysiology and complementary membrane trafficking assays. Nine electrophysiological parameters were integrated using MeTAL to predict variant-specific repolarization phenotypes under homozygous and heterozygous conditions. Modeled QTc values were compared with patient electrocardiographic data and variant-specific penetrance estimates. Functional evidence was calibrated against established benign and pathogenic variants using ClinGen-recommended categorical OddsPath and continuous likelihood-ratio approaches.

**Results:** Multidimensional functional profiling revealed abnormalities not captured by current amplitude alone, identifying additional variants with clinically relevant repolarization defects. Among variants assessed under heterozygous conditions, MeTAL predictions showed 82.9% concordance with patient QTc phenotypes for ACMG class 5 variants, with apparent discordances partly consistent with incomplete penetrance. Continuous likelihood-ratio calibration demonstrated strong discrimination between established benign and pathogenic variants (AUC=0.936) and provided graded functional evidence for 76.1% of VUS. Integration of calibrated functional evidence into the ACMG/AMP framework enabled reclassification of 64 of 201 VUS (31.8%), including 7 as pathogenic, 46 as likely pathogenic, and 11 as likely benign.

**Conclusions:** This nationwide study establishes a scalable framework integrating high-throughput electrophysiology, computational modeling, and clinical phenotyping to translate complex *KCNH2* channel dysfunction into clinically interpretable evidence. The resulting reclassification of nearly one-third of VUS, together with the open-access HAMMER database, provides a resource to improve molecular diagnosis and risk assessment in LQT2.

## Introduction

Variants of *KCNH2*, the gene encoding the cardiac hERG potassium channel, have been implicated in congenital Long QT Syndrome type 2 (LQT2) and Short QT syndrome type 1 (SQT1), rare inherited arrhythmic disorders characterized by delayed or abbreviated ventricular repolarization and an increased risk of sudden cardiac death ^1–4^. The clinical relevance of LQT2 is further underscored by its distinct arrhythmic risk profile compared with LQT1, the comparatively limited protection afforded by beta-blocker therapy, and the particular susceptibility of hERG channels to pharmacological blockade by numerous drugs ^5–7^. Early identification of individuals at risk is essential for optimizing clinical management and implementing preventive strategies. A major challenge in the era of genomic medicine is the rapid accumulation of *KCNH2* variants in public clinical databases, from approximately 1,000 entries in 2021 to more than 3,900 in 2026 (ClinVar: https://www.ncbi.nlm.nih.gov/clinvar), driven by widespread adoption of high-throughput sequencing technologies. Among missense variants, approximately 80% remain classified as variants of uncertain significance (VUS) ^8^ according to the recommendations of the American College of Medical Genetics and Genomics and the Association for Molecular Pathology ^9^, and consequently represent a major focus of ongoing efforts toward clinical reclassification. Determining the pathogenicity of these VUS is a critical priority for accurate diagnosis and management of patients with inherited arrhythmic disorders, as well as for appropriate risk stratification within affected families ^10^.

In the context of *KCNH2*, patch-clamp electrophysiology is considered the gold standard for assessing the functional impact of genetic variants. Beyond functional characterization, electrophysiological studies provide important insights into genotype-phenotype relationships ^8^. Importantly, the ClinGen Sequence Variant Interpretation Working Group has established recommendations for the use of well-validated functional assays as PS3/BS3 evidence, emphasizing validation against established pathogenic and benign controls and calibration of the resulting evidence strength ^10^. Although functional interpretation of VUS has historically lagged behind the pace of variant discovery, coordinated international initiatives are now addressing this gap. Large-scale efforts to systematically characterize *KCNH2* variants are underway, with hundreds of variants already assessed using standardized high-throughput *in vitro* platforms ^11–16^. These initiatives represent an important step toward translating variant detection into clinically relevant interpretation.

Despite their considerable value, these approaches remain subject to important limitations. First, electrophysiological studies lack standardized experimental protocols, making direct and reliable comparisons across studies challenging ^8^. Second, because *KCNH2*-related disorders are inherited in an autosomal dominant manner, expression systems that strictly ensure equal expression of the wild-type (WT) and variant alleles are essential for accurately assessing variant pathogenicity in the heterozygous state. Third, functional analyses often focus on a restricted subset of biophysical parameters ^11,16^. Although hERG channel expression is an established determinant of channel function and disease risk, our recent systematic investigation demonstrated that other biophysical parameters, including channel inactivation, also play a role in determining the pathogenicity of individual variants ^17^. Assessing channel conductance alone therefore fails to capture the full spectrum of functionally relevant properties and may lead to inaccurate estimates of the clinical pathogenicity of hERG VUS. Indeed, alterations in other hERG channel biophysical properties may either exacerbate or compensate for the pathogenic effects of altered channel expression levels. Individual variants often produce a mixture of gain- (GoF) and loss-of-function (LoF) features, rendering their clinical interpretation particularly challenging in the absence of an integrative analytical framework ^18^. To address this complexity, we previously developed MeTAL ^17^, a computational package specifically designed to integrate the full spectrum of hERG biophysical properties that provides QTc evaluations as clinically-relevant assessment of variant pathogenicity. MeTAL establishes a rationale linking channel phenotypes – including gating kinetics and voltage-dependence for activation, inactivation, recovery from inactivation and deactivation besides current density – to patient-level clinical manifestations.

Herein, using a standardized phenotyping pipeline, we systematically phenotyped, at the national level, 303 French variants including 201 VUS (corresponding to 56% of the 358 *KCNH2* variants referenced in the national Cardiogen network (https://www.filiere-cardiogen.fr/)), 21 ACMG class 1 and 2 variants, 43 out of 78 ACMG class 4 and 38 out of 58 class 5 variants. 101 variants characterized were in common with an earlier large-scale phenotyping effort ^11^. We further applied MeTAL as a binary classifier to identify functionally disruptive variants under homozygous conditions across all ACMG classes and subsequently to assess their predicted impact on ventricular repolarization under strict heterozygous conditions. Using this systematic and standardized approach, we stratified the pathogenicity of French *KCNH2* VUS, including variants for which channel expression was preserved. Whenever clinical data were available, MeTAL-based pathogenicity predictions were compared with electrocardiographic phenotypes from patients carrying the corresponding variants. Functional phenotypes were subsequently calibrated against established benign and pathogenic reference variants according to ClinGen recommendations, enabling the assignment of PS3/BS3 evidence strength for clinical variant interpretation. Integration of this functional evidence into the ACMG/AMP classification framework supported the reclassification of 58 VUS as likely pathogenic. Building on these complementary functional, computational, and clinical efforts, we established an open-access, web-based database integrating detailed genetic, clinical, functional, and patient-level data for all 303 variants. This resource provides a comprehensive systematic characterization of *KCNH2* VUS undertaken at the scale of a national cohort and represents an important step toward resolving the clinical interpretation and management of *KCNH2* VUS. It will support clinicians, researchers, and variant curators in the accurate diagnosis and risk stratification of LQT2 and will remain open to the integration and ACMG-based reclassification of newly identified VUS by other research groups adopting our standardized protocol.

## Methods

### Patient variant selection and national Cardiogen database

Clinical and genetic data for 358 *KCNH2* variants were obtained from the Bamacœur database, a national registry from the French Cardiogen network. It compiles detailed clinical and genetic information from patients with inherited cardiac arrhythmias, contributed by a network of French expert centers, including the Inherited Cardiac Arrhythmia Reference Center at Nantes University Hospital, the Inherited Cardiac Diseases Reference Center of Assistance Publique-Hôpitaux de Paris (AP-HP), the Eastern and South-Eastern France Reference Center for Hereditary and Rare Cardiac Rhythm Disorders (CERA, Lyon), and the Competence Centers for Hereditary and Rare Cardiac Rhythm Disorders in Grenoble and Amiens. For this study, we prioritized 303 KCNH2 variants for comprehensive functional characterization (**Table S1**), including 201 variants of uncertain significance (VUS) and 102 previously classified variants (81 pathogenic or likely pathogenic and 21 benign or likely benign), which served as reference controls for benchmarking functional effects and supporting variant reclassification. The remaining 55 variants (35 ACMG class 4 and 20 class 5) were considered of lower priority by clinicians. All patients registered in the Bamacœur database provided written informed consent for the use of their clinical and genetic data for research purposes.

### KCNH2 site-directed mutagenesis

Site-directed mutagenesis of *the KCNH2* coding sequence (NCBI reference sequence: NM_000238.4), cloned between the *Hind*III and *Xho*I restriction sites of the pcDNA5/FRT/TO plasmid, was performed using the Gibson assembly method, as previously described ^18^. For electrophysiological recordings, we used a non-tagged hERG sequence (*Opti-hERG)* to generate the full panel of 303 missense variants analyzed in this study. For trafficking analyses, a pHluorin tag preceded by an additional transmembrane segment was fused to the 3’ end of the hERG coding sequence, yielding an extracellularly exposed pHluorin moiety (Opti-hERG-pHluorin), and used to generate a subset of 73 missense variants. Mutagenic primers were designed to introduce specific single-nucleotide substitutions (**Table S2**). All variants were confirmed by Sanger sequencing. Plasmids were purified using the endotoxin-free Qiagen Plasmid Plus Kit (Qiagen, 12945, Hilden, Germany), according to the manufacturer’s protocol, and quantified prior to electroporation (automated patch-clamp) or transfection (confocal microscopy).

### Cell culture and transient expression of hERG variants

HEK-293T cells (CRL-3216, ATCC) were used for transient expression of *KCNH2* variants. Cells were cultured in Dulbecco’s Modified Eagle’s Medium 4.5 g/L glucose (DMEM; Gibco, France) supplemented with 10% fetal calf serum (Eurobio, France), 2 mM L-glutamine, 100 U/mL penicillin, and 100 µg/mL streptomycin (Corning, France). Cultures were maintained at 37 °C in a humidified incubator with 5% CO₂ and regularly tested for mycoplasma contamination (MycoAlert, Lonza, France). Each of the 303 *Opti-hERG* plasmids was introduced into HEK-293T cells *via* electroporation using OC-100 cassettes and the MaxCyte STx system (MaxCyte Inc., MD, USA), as previously described ^19^. In heterozygous condition, care was taken that an exact 1:1 ratio for WT and variant hERG expression occurred by mixing equal quantities of WT and variant *opti-hERG* plasmids. Thirty hours post-electroporation, cells were detached using Accutase (Sigma-Aldrich, St. Louis, MO, USA) and resuspended in an external NMDG solution (in mM: 80 NaCl, 4 KCl, 2 CaCl₂, 1 MgCl₂, 5 glucose, 60 N-Methyl-D-glucamine (NMDG), 10 HEPES; pH 7.4 adjusted with NaOH; 280 ± 3 mOsm) for automated patch-clamp assays. For membrane expression assays using confocal microscopy, MDCK cells (ATCC CCL-34) were cultured on glass coverslips and transfected with either the WT Opti-hERG-pHluorin plasmid or one of the 73 *Opti-hERG-pHluorin* plasmids using Lipofectamine 2000 (Thermo Fisher Scientific, Grand Island, NY, USA) according to the manufacturer’s protocol, as described previously^18^.

### Protein trafficking assessment by confocal microscopy

Twenty-four hours after transfection, MDCK cells were imaged using an Ultraview Vox spinning disk confocal microscope (PerkinElmer) with a 60×/1.40 oil objective, as previously described ^18^. Cells were perfused with extracellular buffer (in mM: 140 NaCl, 5 KCl, 3 MgCl₂, 1 CaCl₂, 10 HEPES; pH 7.4), followed by acidification buffer (HEPES replaced with 2-(N-morpholino)ethanesulfonic acid (MES), pH 6.0) and returned to neutral pH. The cycle was performed twice in total. Fluorescence intensity was quantified using ImageJ (National Institutes of Health, Bethesda, MD, USA), and surface expression was expressed as the ratio (F₇.₄ − F₆.₀)/F₇.₄. Ionophores (nigericin and monensin, 10 µM) were applied at the end of the protocol to assess total fluorescence, which was found to be nearly identical to that measured at neutral pH (7.4). Three independent transfections were performed for each construct, and 38 to 65 cells were analyzed per variant (median, 47 cells).

### Standardized high-throughput automated patch-clamp recording

The evaluation of hERG variants was performed using an optimized fast-track protocol on the SyncroPatch 384PE system (Nanion), adapted from previously established manual patch-clamp sub-protocols ^18,20^. Key modifications were introduced to accommodate the specific requirements of Nanion’s PatchControl software and to optimize seal quality under high-throughput conditions. The final fast-track protocol consists of six defined sub-protocols (**Figure S1**). Notably, extracellular Ca²⁺ concentration was increased from 1 to 2.88 mM, and intracellular fluoride concentration reached 110 mM to facilitate gigaseal formation. Holding potential was set at -80 mV. Step durations were carefully adjusted to capture the voltage-dependent kinetics of channel gating ^20^. The complete protocol duration was 74 seconds per cell.

Recordings were performed at 22-23 °C using the external NMDG-based solution and an internal solution containing (in mM): 110 KF, 10 NaCl, 10 KCl, 10 EGTA, 10 HEPES (pH 7.2, NaOH), 280 ± 3 mOsm ^20^. Each variant was systematically analyzed the same day along with the WT hERG condition for thorough comparison. Two technical replicates were performed each time in order to record from 192 cells per variant as a starting point.

### Automated electrophysiological data analysis and statistical analyses

All R scripts used to analyze the automated patch-clamp (APC) data are available at https://github.com/jerome-montnach/hERGvariants. Cells with not respecting the quality control criteria – whenever resistance (Ra) > 10 MΩ, seal resistance (Rseal) < 600 MΩ (measured at -86 mV), leak current < -200 pA at -60 mV, or current run-down > 20% over the course of the experiment - were excluded from analysis. Current amplitudes were expressed as square root (sqrt) as recommended for normalization of data distribution ^14^. For extraction of biophysical properties, were considered only recordings with current amplitudes above 500 pA to avoid interference of native K^+^ channels, and below 10 nA to prevent poor voltage-clamp ^21^. To minimize variability, data from channel variants were normalized and compared to WT hERG values obtained on the same day of acquisition. Statistical analyses were performed using parametric t-tests for current amplitudes and single-point biophysical parameters, and ANOVA for kinetic parameters measured across multiple voltages. Significance thresholds were set as follows: *p* < 0.05 (*), *p < 0.01* (**), and *p < 0.001* (***).

### Computational modeling of variant-induced repolarization phenotypes

To translate the multiparametric functional consequences of *KCNH2* variants into a predicted cardiac repolarization phenotype, we used MeTAL (Multiscale-enriched Transformation and Analysis for Long-QT), a previously developed computational framework that integrates experimentally measured hERG biophysical properties into a human ventricular electrophysiological model to generate simulated pseudo-ECGs ^17^. MeTAL incorporates all nine hERG channel electrophysiological parameters, encompassing current amplitude, voltage-dependence, and gating kinetic properties. It generates variant-specific pseudo-ECGs from which QTc intervals were extracted (by convention: pseudo-QTc and modeled-QTc values for homozygous and heterozygous conditions, respectively). Pseudo-QTc values were used as a binary classifier to categorize variants requiring full investigation in heterozygous condition. Only modeled-QTc values were used to reproduce the genetic context of affected individuals. They were categorized as SQT-like (<340 ms), normal (340-470 ms), or LQT-like (>470 ms).

### Z-score normalization

Z-scores were calculated using two different reference strategies depending on the intended analysis. For the radar plots, biophysical parameters were normalized relative to the WT control. For each biophysical parameter, except current amplitude, the Z-score was calculated as: 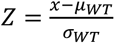, where x is the value measured for the variant, and *μ*_W*T*_ and *σ*_W*T*_ are the mean and standard deviation of the WT measurements. Radar plots were expressed in units of WT standard deviation, such that ±1 corresponded to ±1 Z-score relative to the WT distribution. Since current amplitudes lack normal distribution, they were square-root transformed to reach a normal distribution. For the comparative analyses of current amplitude, and MeTAL-generated pseudo and modeled QTc values, Z-scores were calculated using the distribution of benign variants (21 benign or likely benign variants; ACMG classes 1-2). For each parameter, the Z-score was calculated as: 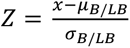, where *μ_B_*_/*LB*_ and *σ_B_*_/*LB*_ correspond to the mean and standard deviation of the benign/likely benign reference group.

### Calibration of functional evidence for ACMG/AMP variant interpretation

Variants with established ACMG classifications were used as reference controls for calibration of functional evidence based on MeTAL modeled-QTc, in accordance with ClinGen recommendations and the approach previously applied to *KCNH2* functional data ^11^. Benign/likely benign variants (B/LB; ACMG classes 1-2; 20 variants) constituted the benign reference group, whereas pathogenic variants (P; ACMG class 5; 38 variants) constituted the pathogenic reference group. Likely pathogenic variants (LP; ACMG class 4) were subsequently evaluated using the calibrated models along with VUS (ACMG class 3). A190T (ACMG class 1) was not used as a benign reference for calibration because of conflicting classifications reported in ClinVar. Functional evidence was calibrated using two complementary approaches: a categorical Odds of Pathogenicity (OddsPath) framework and a continuous log-likelihood ratio (LLR) framework.

OddsPath was calculated for the MeTAL modeled-QTc values according to ClinGen recommendations for the application of ACMG/AMP PS3 and BS3 functional evidence, using the approach established for *KCNH2* ^11^. Reference variants were cross-classified according to their clinical classification and functional phenotype into four categories: B/LB variants with a normal (A) or abnormal (B) functional result, and P variants with a normal (C) or abnormal (D) functional result. All 20 B/LB reference variants displayed a normal functional phenotype (A = 20, B = 0), whereas 7 of the 38 P variants displayed a normal phenotype and 31 displayed an abnormal phenotype (C = 7, D = 31). Because no B/LB reference variant displayed an abnormal functional result (B = 0), an add-one correction was applied to avoid a zero cell when estimating the posterior probability associated with an abnormal result, as recommended for OddsPath calibration ^11^. This yielded OddsPath values of 16.32 for an abnormal functional result and 0.184 for a normal functional result (**Table S3**). According to ClinGen Bayesian evidence thresholds, these values corresponded to PS3_moderate and BS3_moderate evidence, respectively. These assay-specific evidence strengths were subsequently assigned to variants outside the reference set according to their categorical phenotype. An independent OddsPath calibration was not performed for the trafficking assay because the number of available reference variants, particularly B/LB controls, was insufficient to support robust assay-specific calibration. Instead, trafficking results were interpreted using the previously established continuous LLR calibration reported for *KCNH2* ^11^.

The continuous log-likelihood ratio model was developed using MeTAL-modeled QTc with the same reference set of 58 variants. Because MeTAL generates pseudo and modeled QTc values on a common phenotypic scale, measurements obtained under either homozygous (for variants with a normal functional phenotype) or heterozygous conditions were included in the calibration. Because the pathogenic reference group comprises LoF variants associated with QT prolongation, the LLR model quantifies functional evidence only along the LoF/LQT spectrum. Accordingly, benign evidence derived from this model was not considered applicable to variants exhibiting an SQT-like phenotype (<340 ms), as marked QT shortening may instead reflect a pathogenic GoF mechanism. SQT-like variants were therefore evaluated separately. Logistic regression was used to model the probability of belonging to the pathogenic reference group as a function of MeTAL-modeled *QTc*(*x*): *logitPP* ∣ *x* = *β*0 + *β*1*x*. The odds predicted by the logistic model were normalized to the prior odds of pathogenic *versus* B/LB variants in the reference dataset to derive a continuous likelihood ratio: 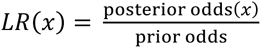, with 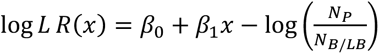 A. The resulting LLR values were converted into graded strengths of functional evidence using Bayesian LLR thresholds of 2.08, 4.33, 18.7, and 350, corresponding to supporting, moderate, strong, and very strong evidence toward pathogenicity, respectively. Reciprocal thresholds were used to assign the corresponding strengths of evidence toward benignity. LLR values falling between the supporting pathogenic and benign thresholds were considered functionally uninformative. Model discrimination was assessed by receiver operating characteristic (ROC) analysis and quantified using the area under the ROC curve (AUC). Following calibration, the continuous LLR model was first applied to ACMG class 4 variants as an independent group and subsequently to ACMG class 3 variants to estimate variant-specific functional evidence strength. Statistical analyses were performed using an R script adapted from O’Neill et al. ^11^, with modifications for the present *KCNH2* dataset and MeTAL-modeled QTc values.

### Variant-specific penetrance estimation

Variant-specific penetrance estimates were obtained using the Bayesian framework previously developed to account for incomplete penetrance in inherited cardiac channelopathies and subsequently applied to variant-specific risk estimation in *KCNH2*-associated LQT2 ^11,22^. The previously reported estimates were updated by incorporating clinical data from the present French cohort (**Tables S1** and **S4**). Analyses were performed using the Bayesian beta-binomial framework and the publicly available code provided by O’Neill et al. ^11^. Data from the literature and the independent Italian and Japanese cohorts included in the original analysis were retained, and affected and unaffected heterozygous carriers from the present French cohort were added to update the variant-specific posterior penetrance estimates. Briefly, variant-specific Bayesian priors were derived from the literature-based *KCNH2* penetrance model ^11^, in which prior penetrance estimates were generated using an expectation-maximization framework incorporating variant-level features. For each variant *i*, the prior distribution was represented by the parameters *α*_prior,*i*_ and *β*_prior,*i*_ of a beta-distribution. These priors were combined with the carrier data described above.

The French cohort comprised 277 unique patient-variant records. Individuals clinically classified as affected were considered affected heterozygous carriers, whereas those classified as normal were considered unaffected heterozygous carriers. Individuals with an unclear phenotype, no available ECG, or missing clinical information were considered non-informative and excluded from penetrance estimation. For each variant, the posterior beta-distribution parameters were calculated by combining the variant-specific prior with the numbers of affected and unaffected carriers across the literature, Italian, Japanese, and French cohorts: α_post,*i*_ = α_prior,*i*_ + *N*_affected,lit,*i*_ + *N*_affected,Italy,*i*_ + *N*_affected,Japan,*i*_ + *N*_affected,French,*i*_, and β_post,*i*_ = β_prior,*i*_ + *N*_unaffected,lit,*i*_ +*N*_unaffected,Italy,*i*_ + *N*_unaffected,Japan,*i*_ + *N*_unaffected,French,*i*_. Variant-specific penetrance was estimated as the mean of the posterior beta distribution as: 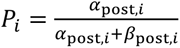. Uncertainty was represented using the 90% Bayesian credible interval, defined by the 5th and 95th percentiles of the posterior beta distribution. Variant-specific Bayesian priors were available for 184 of the 185 variants represented in the French cohort, the remaining variant was assigned a non-informative prior.

### ACMG/AMP reclassification of variants of uncertain significance (VUS) using a Bayesian point-based framework

The 201 *KCNH2* VUS were subjected to systematic re-evaluation using the ACMG/AMP framework ^9^, converted into the quantitative Bayesian point-based system proposed by Tavtigian et al. ^23,24^ (**Table S5**). The objective of this analysis was to determine whether incorporation of the functional evidence generated in the present study was sufficient to resolve the original VUS classification.

For each variant, the ACMG/AMP evidence criteria underlying the original classification were retrieved and retained at their previously assigned strength. Pre-existing functional evidence (PS3 or BS3), when present, was excluded before recalculation to avoid double counting with the newly generated functional evidence. The remaining evidence criteria were converted into points according to the naturally scaled system of Tavtigian et al. ^23^: pathogenic supporting, moderate, strong, and very strong evidence contributed +1, +2, +4, and +8 points, respectively, whereas benign supporting, moderate, and strong evidence contributed −1, −2, and −4 points, respectively (**Tables S5** and **S6**). The resulting sum was considered the baseline non-functional evidence score for each variant.

New functional evidence obtained from MeTAL-modeled QTc was incorporated according to the ClinGen Sequence Variant Interpretation Working Group recommendations for application of the PS3/BS3 criteria ^10^, using two calibration approaches. In the categorical OddsPath-based approach, the strength of functional evidence was assigned on the basis of the previously determined OddsPath values. Accordingly, variants displaying a deleterious functional phenotype were assigned PS3_moderate (+2 points), whereas variants displaying a functionally normal phenotype were assigned BS3_moderate (−2 points). In parallel, continuous LLR calibration was used to assign variant-specific functional evidence strengths, ranging from supporting to very strong for pathogenic evidence (PS3: +1, +2, +4, or +8 points) and from supporting to strong for benign evidence (BS3: −1, −2, or −4 points). Variants falling within the uninformative LLR interval received no additional functional evidence, while variants displaying SQT-like phenotypes were not assigned LLR-based evidence because the calibration was not applicable to QT shortening.

For each of the 201 VUS, final point scores were calculated independently for the OddsPath- and LLR-based approaches by algebraically summing the baseline non-functional evidence score and the corresponding newly assigned functional evidence points. Final classification was determined using the Tavtigian et al. ^23^ thresholds: benign (≤−7 points), likely benign (−6 to −1), VUS (0 to 5), likely pathogenic (6 to 9), and pathogenic (≥10 points). Variants with final scores within the 0-5-point interval were retained as VUS, whereas variants reaching the corresponding pathogenic or benign thresholds were considered reclassified (**Table S6**).

### Development of an open-access KCNH2 variant database

The database was implemented in Python using the Streamlit framework and deployed at https://hammer.bird.glicid.fr. A file-based pipeline processes electrophysiological, clinical, and *in silico* prediction data (SPARC, MeTAL) from CSV files. We used the Pandas library to normalize variant labels and functional summaries, to filter records with missing positions or ACMG classifications, and to produce standardized, variant-level records. The interface integrates Plotly interactive charts, a py3Dmol 3D hERG channel viewer (PDB 5VA1), and an external Protter 2D rendering link. Local sources include variant and clinical tables, SPARC/MeTAL CSV scores, ClinGen and ECG spreadsheets, and the PDB structure; ClinVar and gnomAD data appear as embedded fields or links rather than API queries. The source code is available at https://github.com/Herg-Database/Hammer.

## Results

### Variant classification and clinical dataset

Clinical and genetic data for 358 *KCNH2* variants were retrieved from the Bamacœur database ^20^. Clinical data were available for 277 patients (**Figure 1*a*, Table S4**). According to ACMG guidelines, approximately 56% (n = 201) of variants were classified as VUS, 38% (n = 136) as LP and P (ACMG classes 4 & 5), and 6% (n = 21) as B and LB (ACMG classes 1 & 2) (**Figure 1*b***). This distribution highlights the predominance of VUS among *KCNH2* variants encountered in clinical diagnostic testing in France. We next compared QTc values among patients according to ACMG variant classification (**Figure 1*c***). Patients carrying LP (class 4) and P (class 5) variants predominantly exhibited QTc durations above the normal clinical range, with 72% and 81% of patients, respectively, showing prolonged QTc values. Patients carrying VUS exhibited a broader and intermediate distribution of QTc values, underscoring the challenge of interpreting the clinical significance of these variants. Although patients carrying ACMG class 1 or 2 variants generally had lower QTc values than those carrying class 4 and 5 variants, 28.5% exhibited QTc values outside the physiological range. In these patients, the prolonged QTc values are not considered to be attributable to the identified *KCNH2* variants. We then examined the distribution of variants within the cryo-EM structure of the hERG channel according to their ACMG classification (**Figure 1*d***). Pathogenic variants, including LP variants, tended to cluster within key functional domains, including the pore region, the voltage-sensing domain (VSD), and the C-linker/cyclic nucleotide-binding homology domain (CNBHD). The distribution of variants across hERG structural domains (**Figure 1*e***) further illustrated this pattern, with pathogenic variants predominantly located within these domains, whereas VUS were more broadly distributed across the channel structure. These findings are consistent with a previous study that used SPARC, a structural pathogenicity algorithm, as a binary classifier of hERG variant pathogenicity ^20^.

**Figure 1:**
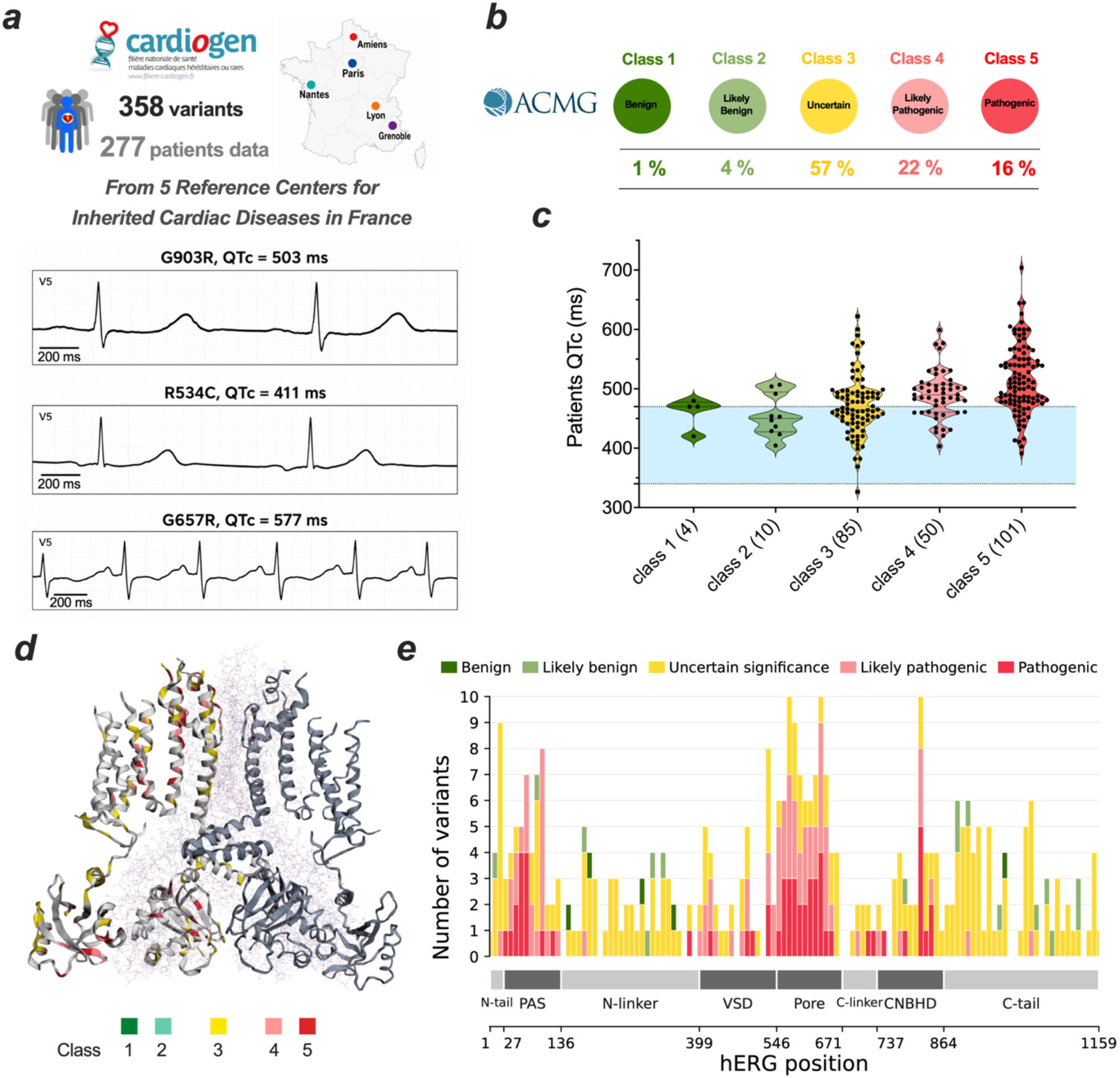
Distribution and clinical impact of 358 *KCNH2* variants identified in the French Cardiogen cohort. ***a***, Summary of variant and patient data retrieved from the Bamacœur national registry. Representative lead V5 ECG recordings from patients carrying the G903R, R534C, and G657R variants illustrate the range of observed QTc intervals. ***b***, Classification of the 358 variants according to ACMG guidelines; percentages as shown. ***c***, Violin plot showing the distribution of patient QTc values according to ACMG variant classification. The number of patients with available ECG data in each class is indicated below the corresponding group. The light blue shaded area indicates the normal QTc range (340 - 470 ms). ***d***, Structural mapping of all French variants onto the cryo-EM structure of the hERG channel, color-coded according to ACMG classification. ***e***, Distribution of variants across hERG structural domains, stratified by ACMG classification. Structural domains are indicated below the plot.

### Current amplitude alone does not fully capture hERG variant dysfunction under homozygous conditions

As a first step toward assessing how the 201 French VUS may disrupt channel function, we characterized 303 *KCNH2* variants using high-throughput APC (**Figure 2, Table S1**). This dataset included all ClinVar reported ACMG class 1 & 2 variants (21) and a large fraction of the ACMG class 4 & 5 variants (81 of 136; 60%), which served to validate our approach and identify the subset of variants requiring characterization under heterozygous conditions. An optimized patch-clamp protocol designed for APC conditions enables the extraction of nine essential hERG channel biophysical parameters (**Figure S1**)^20^. All variant parameters were normalized to WT values recorded on the same day to minimize batch fluctuations (**Figure S2**). The distribution of the square root (sqrt) of normalized current amplitude for the 303 characterized variants is shown (**Figure 2*a***). ACMG class 1 and 2 variants were used to define the benign reference distribution and the normal range corresponding to a Z-score between −2 and +2. Among the 201 VUS, 53 (26.4%) fell outside this normal Z-score range. VUS falling within the normal range cannot necessarily be considered benign because of the pathogenic importance of other biophysical properties. In agreement, several ACMG class 4 and 5 LP and P variants also displayed normal current-amplitude Z-scores: 15 of 43 class 4 variants (34.9%) and 1 of 38 class 5 variants (2.6%). This finding confirms that dysfunction cannot be attributed solely to defects in current amplitude ^17^. Overall, based on current amplitude measurements, 114 of the 303 variants (37.6%) exhibited a LoF phenotype, whereas 7 (2.3%) exhibited a GoF phenotype (**Figure 2*b***). We next examined the distribution of these functional phenotypes across the different structural domains of hERG (**Figure 2*c***). LoF variants were concentrated in the structured PAS, VSD, pore, and CNBHD domains. In contrast, variants located in the less well-characterized regions of the N-tail, N-linker and C-tail predominantly displayed normal current amplitudes. Notably, the few benign class 1 and 2 variants are located predominantly within these less structured regions, supporting the notion that functionally disruptive variants are enriched within structured domains of hERG ^20^.

**Figure 2.**
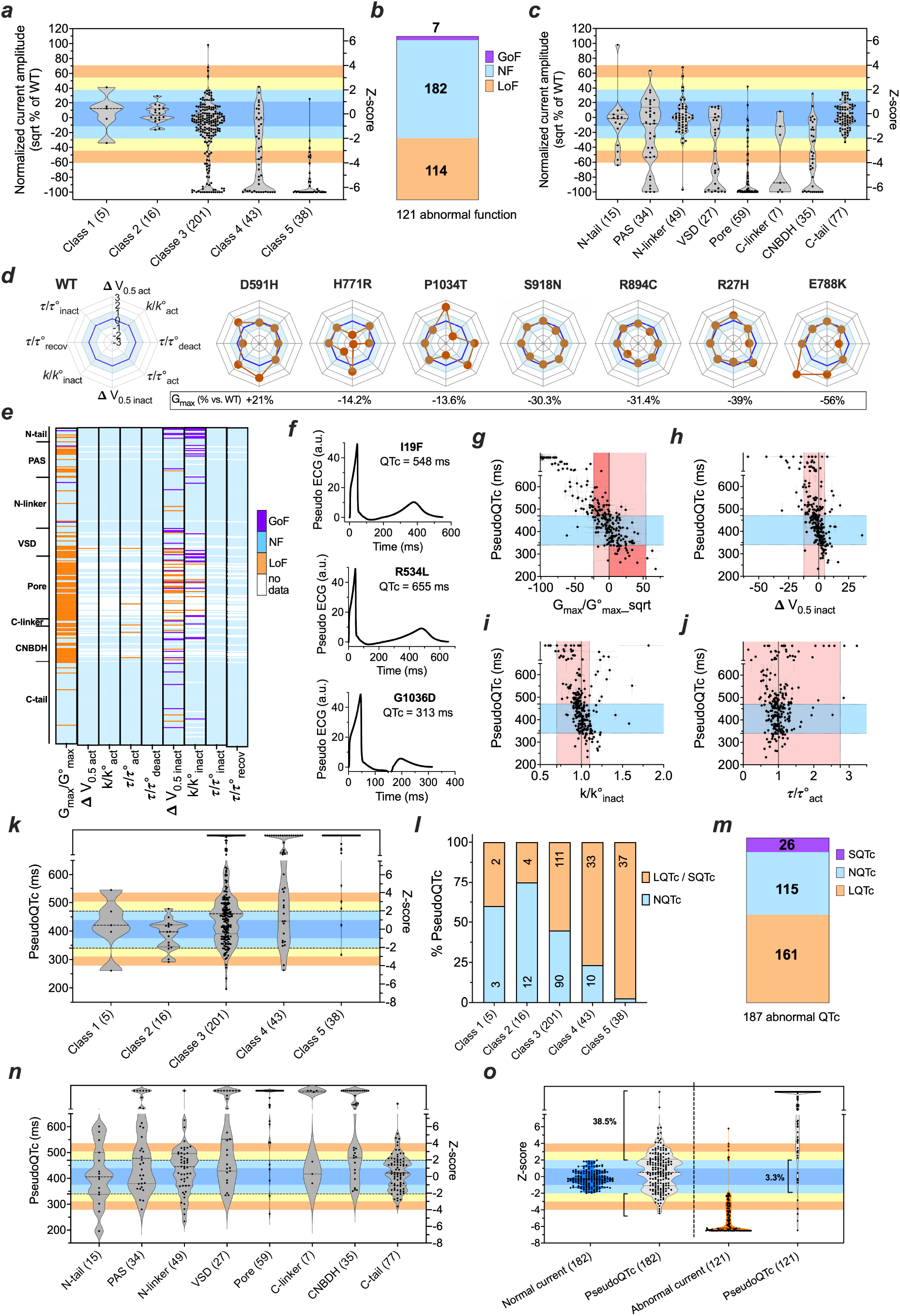
Integrated functional characterization of French hERG variants under homozygous conditions. ***a,*** Distribution of normalized current amplitude (square-root-transformed Z-scores) for 303 variants. The normal range (−2 ≤ Z ≤ 2) was defined based on ACMG class 1-2 variants. ***b,*** Classification of hERG variants based on square-root current amplitude (% of WT), identifying 114 LoF, 182 normal function (NF), and 7 GoF. ***c,*** Distribution of current amplitudes across hERG structural domains, highlighting an enrichment of LoF variants in structured regions, including the PAS, VSD, pore, and CNBHD. The number of variants in each domain is indicated in parentheses. ***d,*** Representative radar plots illustrating variant-specific biophysical fingerprints across eight gating parameters. Each axis represents a normalized biophysical parameter, and concentric lines indicate Z-score deviations relative to WT. The blue shaded area represents the WT range (±1 Z-score), allowing direct visualization of the magnitude and direction of changes (GoF or LoF) for each parameter. The variation in current amplitude relative to WT (G_max_, % vs. WT) is indicated below each radar plot. The plots illustrate three representative categories of variant effects: (i) no significant change in current amplitude despite significant alterations in biophysical properties (D591H, H771R, P1034T); (ii) minor changes in current amplitude combined with other minor alterations in biophysical properties, illustrating cumulative channel defects (S918N, R894C, R27H); and (iii) major alterations in current amplitude compensated by GoF changes in other biophysical properties (E788K). ***e,*** Heatmap showing GoF, NF, and LoF predictions generated by MeTAL for alterations in each individual biophysical parameter for each variant (n = 303 for current amplitude; n = 245 for all other parameters with measurable currents), organized according to hERG structural domains. MeTAL-defined pathogenicity thresholds are: (i) current amplitude (LQT: < −30% vs. WT; SQT: > 120% vs. WT); (ii) τ_act_ (τ/t^0^_act_ > 2.75 for LQT); (iii) V_0.5,inact_ (LQT: ΔV_0.5,inact_ < −12.5 mV; SQT: > 5 mV); and (iv) k_inact_ (k/k ^0^_inact_ < 0.7 for LQT; > 1.1 for SQT). ***f,*** Representative simulated ECG traces generated by MeTAL using the variant-specific biophysical parameters of the I19F, R534L and G1036D variants, corresponding to the radar plots shown in ***d***. All nine biophysical parameters were integrated by MeTAL to generate the simulated ECG traces. ***g-j,*** Plots of predicted pseudoQTc values as a function of individual biophysical parameters for all variants. PseudoQTc values were plotted against the square-root current amplitude (***g***, n=303 variants), inactivation parameters (***h-i***, n=245 variants), and activation time constant (τ_act; ***j***, n=245 variants). The rose/red-shaded areas indicate parameter ranges within which minor changes in the individual parameters are predicted by MeTAL to result in normal pseudoQTc values. ***k,*** Distribution of pseudoQTc values across ACMG classes. Z-scores were calculated based on the QTc values of variants falling within the normal QTc range (340 to 470). ***l,*** Percentage of normal and dysfunctional variants according to pseudoQTc classification for the five ACMG classes. Dysfunctional variants include those predicted to produce long pseudoQTc values (LQTc) or short pseudoQTc (SQTc). NQTc = normal pseudoQTc value. ***m,*** Number of variants classified according to their pseudoQTc category (NQTc, LQTc, or SQTc). ***n,*** Distribution of pseudoQTc values across hERG structural domains, highlighting increased detection of dysfunction in previously underappreciated regions, including the N-linker and C-tail. ***o,*** Comparison of current amplitude and pseudoQTc Z-scores for variants with normal versus abnormal current amplitudes. Percentages indicate the proportion of variants misclassified by the square-root current amplitude (% of WT) approach.

### Membrane trafficking defects explain most, but not all, loss-of-function phenotypes

A subset of 73 variants was further characterized for membrane-trafficking defects using chimeric constructs containing a pHluorin tag, which allows discrimination between the pool of channels expressed at the plasma membrane and total channel expression ^18^ (**Figure S3*a-c***). This subset comprised one ACMG class 1 variant, 27 VUS (2 GoF and 19 LoF based on current amplitude measurements), and 45 class 4 and 5 variants (**Figure S3*e***). Based on current-amplitude Z-scores, eight of these variants displayed normal current amplitudes, whereas 65 exhibited either a GoF (n=2) or LoF (n=63) phenotype (**Figure S3*d***). Among the 63 variants with LoF current amplitudes (Z-score < −2), 55 (87.3%) also showed impaired membrane trafficking (Z-score < −2), whereas 8 retained trafficking within the normal range. All variants with normal current amplitude, mostly located beyond residue 846, showed no marked trafficking defects, consistent with previous observations that missense variants in the distal C-tail are generally well tolerated with respect to hERG trafficking ^11^. Interestingly, the two GoF VUS, both located in the N-linker domain, showed a slight increase in trafficking, although this difference did not reach statistical significance. The only LoF variants that displayed normal trafficking were located in the pore or PAS domains, suggesting that their LoF phenotype may result primarily from altered gating rather than impaired membrane expression (**Figure S3*d***). Variants located in the VSD, C-linker and CNBHD showed the most pronounced trafficking defects, whereas variants in the PAS and pore domains exhibited milder effects (**Figure S3*b-d***). Overall, membrane trafficking was strongly associated with current amplitude (p < 0.0001), indicating that defective membrane expression accounts for the majority, but not all, of the LoF phenotypes observed in this variant set. Trafficking defects were also enriched among variants with higher ACMG classifications (**Figure S3*e***). Class 4 and 5 variants showed a high proportion of reduced membrane trafficking, whereas the only benign variant analyzed, P347S (class 1), displayed normal trafficking. Trafficking measurements were significantly correlated with the O’Neill et al. MAVE trafficking scores reported previously ^11^ for the 73 variants common to both datasets (Pearson r = 0.804, R² = 0.647, p = 1.0 × 10⁻¹⁷; Spearman ρ = 0.685, p = 2.4 × 10⁻¹¹; **Figure S3*f***). Notably, eight variants classified by the MAVE dataset as having near-absent trafficking showed substantial membrane expression in our trafficking assay, consistent with measurable hERG currents as assessed by our APC recordings (**Figure S3*g***). Based on this strong concordance, the LLR-calibrated trafficking thresholds previously established ^11^ were applied to our pHluorin measurements to assign graded levels of functional evidence (**Figure S3*h***). Among the 19 LoF VUS, 17 showed impaired trafficking based on Z-score classification. Of these, 12 reached LLR-calibrated levels of pathogenic evidence, ranging from PS3_supporting to PS3_very strong (**Figure S3*e,h***; **Table S1**).

### Multidimensional biophysical profiling reveals complex functional effects of hERG variants

Since pathogenic variants may display normal expression levels (**Figure 2*a***), their intrinsic dysfunction should result from alterations in other biophysical properties. We therefore investigated the complete set of biophysical properties of variants for which high-quality recordings and measurable currents were obtained (245 of the 303 variants; **Table S1**). For each variant, the eight essential biophysical parameters were represented as radar plots, indicating GoF and LoF features and compared with the WT condition and its standard deviation (±1 Z-score) (**Figure 2*d***, **Figure S4**). Typical examples include variants with no change in current amplitude compared with WT but with significant alterations in biophysical properties (D591H, H771R, P1034T); variants with minor alteration in current amplitude combined with other minimal changes in biophysical properties, potentially explaining synergistic effects (S918N, R894C, R27H); and, finally, variants with major alteration in current amplitude that is counteracted by GoF changes in other biophysical properties (E788K) (**Figure 2*d***). These examples also illustrate that many variants display both LoF and GoF changes in their biophysical properties (i.e., D591H, H771R, P1034T, R27H and E788K). As clearly demonstrated by these examples, a given variant can modify several biophysical properties in a complex manner, making it difficult to derive a global LoF or GoF phenotype. Among the 232 radar plots presented, which include only variants with sufficient current for analysis (**Figure S4**), 72 (31%) display combinations of LoF and GoF features, further complicating the interpretation of channel dysfunction.

MeTAL provides a single clinically-relevant functional readout capable of integrating all nine hERG biophysical properties of a given variant ^17^. Under homozygous conditions, the resulting pseudoQTc values are not directly comparable with clinical QTc but have the advantage to classify the intrinsic pathogenicity of hERG variants. For each individual parameter taken in isolation, we first assessed whether the corresponding pseudoQTc values fell within the LoF, NF or GoF range defined by MeTAL for all 303 variants. The results are presented as a heat map showing all nine parameters for the 303 variants, together with their corresponding structural domains **(Figure 2*e***). While all 303 variants could be represented for current amplitude, only 245 variants were investigated for each other property since 58 variants had no measurable currents at all. This analysis confirms that current amplitude is a major determinant of whether a variant produces LoF or GoF, as previously demonstrated ^17^ (LQT: < −30% vs. WT; SQT: > 120% vs. WT). Interestingly however, several other biophysical parameters should not be neglected. Hence, inactivation parameters also contribute critically, including shifts in V_0.5,inact_ (LQT: ΔV_0.5,inact_ < −12.5 mV; SQT: > 5 mV) and alterations in slope of inactivation (k/k^0^_inact_ < 0.7 for LQT; > 1.1 for SQT). Individually, these two parameters are sufficient per se to produce either GoF or LoF values. Activation time constant also appears to be a critical parameter to assess pathogenicity in some variants.

We therefore used MeTAL as a binary classifier to identify variants predicted to cause severe channel dysfunction; this time by incorporating all nine biophysical alterations to assess the global phenotype. Examples of the predicted pseudo-ECGs generated by MeTAL are shown (**Figure 2*f***). To assess the contribution of individual biophysical parameters to the general predicted repolarization phenotype, pseudoQTc values were plotted against changes in each parameter. Notably, a substantial proportion of variants exhibited abnormal pseudoQTc values despite individual parameters remaining within the normal range, highlighting the contribution of other biophysical alterations to the overall functional phenotype (**Figure 2*g-j***, **Figure S5**). Among the 89 variants whose square-root-transformed current amplitude (% of WT) remained above −22.7%, corresponding to a reduction of less than 30% in normalized current amplitude, 35 (39.3%) nonetheless exhibited severe channel dysfunction. These variants (upper dark red rectangle, **Figure 2*g***) fell in a range in which current reduction alone would be considered insufficient to support pathogenicity ^17^. Conversely, 19 of 92 variants (20.7%) reached SQT-like pseudoQTc values despite amplitude increases below +54% (lower dark red rectangle, **Figure 2*g***). Many variants exhibited only minor alterations in the inactivation parameters V_0.5,inact_ and *k*_inact_ (**Figure 2*h,i***), which, when considered individually, would be insufficient to produce abnormal pseudoQTc values. Nevertheless, these variants were predicted to cause either prolonged pseudoQTc values (approximately 40% of these variants) or shortened pseudoQTc values (approximately 8%) if alterations in other biophysical properties are considered as well. Comparable proportions were obtained for the activation time constant (τ _act_), despite alterations that would not be expected to substantially affect repolarization when considered in isolation (**Figure 2*j***). For the five remaining biophysical parameters, even substantial individual alterations were not expected to produce out-of-range pseudoQTc values when considered independently (**Figure S5**). Together, these findings demonstrate that the functional consequences of *KCNH2* variants cannot be reliably inferred from individual biophysical parameters alone, highlighting the importance of integrating multiple biophysical alterations to predict their impact on ventricular repolarization.

### MeTAL improves the detection of dysfunctional hERG variants under homozygous conditions

The pseudoQTc values generated by MeTAL on the basis of all nine parameters were plotted according to ACMG classification (**Figure 2*k***). These data confirm that a large proportion of class 1 and 2 variants display normal pseudoQTc values (**Figure 2*l***). Interestingly, however, 6 of these 21 B and LB variants (28.6%) produce dysfunctional channels under homozygous conditions, suggesting that these variants should also be studied under heterozygous conditions, in which the dysfunction may be corrected. Among the 201 VUS, MeTAL classified 111 as producing dysfunctional hERG channels (55.2%), warranting further investigation under heterozygous conditions. Based on these results, 90 French VUS were predicted to exhibit normal channel function under homozygous conditions, providing functional evidence supporting benignity. Importantly, the proportion of variants predicted to produce dysfunctional channels increased to 76.7% for class 4 variants (33 of 43) and 97.4% for class 5 variants (37 of 38), demonstrating the strong concordance between pseudoQTc-based functional predictions and ACMG classification. Overall, 61.7% of the variants (187 of 303) were predicted to produce dysfunctional hERG channels based on pseudoQTc values, including 8.6% associated with short pseudoQTc and 53.1% with long pseudoQTc values (**Figure 2*m***). MeTAL identified 66 additional dysfunctional variants under homozygous conditions (**Figure 2*m***) compared with the classification based solely on the current amplitude (121, cf. **Figure 2*b***). Accordingly, based on current amplitude alone, we would have selected 39.9% of the 303 hERG variants for characterization under heterozygous conditions, compared with 61.7% when using the MeTAL classification.

The distribution of pseudoQTc values across the different hERG channel domains is shown (**Figure 2*n***) and can be compared with the corresponding distribution based on square-root current amplitude (% of WT) (**Figure 2*c***). In particular, many of the additional dysfunctional variants identified by pseudoQTc analysis are localized within the N-linker and C-tail domains, where 53% and 35% of variants, respectively, were abnormal, compared with only 14% and 1.3% based on square-root current amplitude (% of WT).

We also examined the relationship between variant location, SPARC-predicted structural impact, and functional phenotype (**Figure S6**). Among the 116 variants displaying normal pseudoQTc values, irrespective of their ACMG classification, 75 (64.6%) were located within intrinsically disordered regions of hERG, including 24 in the N-linker, 50 in the C-tail, and one within a disordered segment of the VSD (**Figure S6*a***). Variants located in structured domains, such as the PAS, pore, and CNBHD, were less frequent. This distribution is consistent with the greater tolerance of intrinsically disordered regions to amino acid substitutions, as previously predicted by the SPARC binary classifier ^20^. We next compared SPARC predictions directly with the experimentally measured functional phenotypes (**Figure S6*b,c***). Using a SPARC score >3.25 as the threshold for predicted structural impact, 36 of 50 variants (72%) above this threshold displayed abnormal current amplitudes (**Figure S6*b***). Concordance was even higher when considering the MeTAL-derived pseudoQTc phenotype, with 44 of 55 variants (80%) predicted by SPARC to have a structural impact displaying an abnormal pseudoQTc phenotype (**Figure S6*c***). Thus, variants predicted by SPARC to affect hERG structure were frequently associated with experimentally detectable functional abnormalities, particularly when channel dysfunction was integrated into the pseudoQTc phenotype.

We next compared square-root amplitude Z-scores with pseudoQTc Z-scores for all 303 variants under homozygous conditions (**Figure 2*o***). Variants with a normal square-root amplitude (−2 ≤ Z ≤ 2) displayed a broad distribution of pseudoQTc values, with 38.5% exhibiting abnormal pseudoQTc Z-scores (Z < -2 or Z > 2), indicating that pseudoQTc analysis substantially increases the identification of dysfunctional variants. Conversely, among variants with an abnormal square-root amplitude (Z < -2 or Z > 2), only a small fraction (3.3%) displayed pseudoQTc values within the normal range. This finding indicates that most variants exhibiting abnormal current amplitudes are also identified as dysfunctional by pseudoQTc analysis. This finding further supports a major contribution of biophysical parameters beyond conductance alone to the functional consequences of hERG variants.

### Functional screening of 187 dysfunctional hERG variants under heterozygous conditions

Based on the discriminative power of pseudoQTc values, 187 variants predicted to cause channel dysfunction under homozygous conditions (111 VUS, 2 class 1, 4 class 2, 33 class 4, and 37 class 5) were selected for further investigation under heterozygous conditions. In addition, seven variants with normal pseudoQTc values (1 class 2, 4 class 3, 1 class 4, and 1 class 5) were included as controls to validate our experimental approach. The distribution of normalized current amplitudes (square-root-transformed Z-scores) for the 187 selected variants under heterozygous conditions was compared with that of benign and likely benign variants (ACMG classes 1 and 2) characterized under the same experimental conditions (**Figure 3*a*, Table S1**). All six class 1 and 2 variants displayed normal Z-scores. Among the 111 class 3 variants, 1 displayed a clear GoF effect on current amplitude (Z-score > 2), whereas 44 displayed a LoF effect (Z-score < -2), resulting in a total of 45 dysfunctional variants (40.5%) under heterozygous conditions. Among the combined class 4 and 5 variants, 38 of 70 (54.3%) displayed a LoF effect on current amplitude (Z-score < -2; **Figure 3*a***). These data illustrate that current amplitude alone is insufficient to identify the full spectrum of dysfunctional class 4 and class 5 hERG variants. We also calculated Z-scores for the eight remaining biophysical parameters in 183 variants characterized under heterozygous conditions, excluding four variants with insufficient residual current for parameter extraction (I642L, R823P, G604S and W585C; **Figure S7**). Although alterations in channel gating were observed across ACMG classes, most individual deviations from WT hERG were relatively modest, with Z-scores generally ranging between −2 and +2. Radar plots for all variants characterized under heterozygous conditions are provided for detailed comparison (**Figure S8**). We again applied MeTAL to integrate and combine all nine biophysical parameters in assessing variant dysfunction. Representative modeled ECGs and corresponding mean QTc values for eight variants from each ACMG class are shown in **Figure 3*b***. Modeled ECGs and mean QTc values for characterized variants are provided (**Figure S9**, **Table S1**). The distribution of modeled QTc values according to ACMG class is shown (**Figure 3*c*, Table S1**). We next evaluated how QTc values predicted by MeTAL changed from the homozygous condition (pseudoQTc values) to the heterozygous condition (modeled QTc values) (**Figure 3*d-g***). Overall, coexpression of the WT channel tended to normalize QTc values: values above 470 ms generally decreased, whereas values below 340 ms increased. This effect was observed across all ACMG classes. Interestingly, markedly abnormal pseudoQTc values (> 650 ms) were frequently reduced substantially following WT coexpression. The vast majority remained however within the pathological long QTc range. Indeed, among the 95 variants with severely abnormal pseudoQTc values, only 11 (4 class 3, 3 class 4, and 4 class 5) were normalized under heterozygous conditions with modeled QTc values between 345 ms and 465 ms (11.6%). Conversely, a further 11 of these 95 variants remained severely abnormal according to their modeled QTc values under heterozygous conditions (3 class 3, 3 class 4, and 5 class 5). These variants therefore retained a marked functional defect despite WT coexpression, suggesting strong dominant-negative effects. They included P846A, P846L, G925A in class 3; P596T, G626R, E637Q in class 4; and W585C, A561V, W568C, G604S, N633S in class 5. Among class 4 and 5 variants, long-QT phenotypes predominated and generally shifted toward normalization under heterozygous conditions. In contrast, class 3 variants displayed a more heterogeneous phenotype, with long-QT variants predominating but with a substantial proportion of short-QT variants also observed (**Figure 3*h***). Interestingly, one class 1 variant, A190T, with a modeled QTc value of 513.7 ms, was predicted to produce a pathogenic LQT2 phenotype despite current amplitude and all other biophysical parameters remaining within the normal range. This observation illustrates how several small deviations in biophysical properties, each individually within the normal range, can combine to produce an abnormal repolarization phenotype. The finding is consistent with the conflicting ClinVar classification of A190T, with some submissions assigning it to class 1 and others to class 3. Among class 3 variants, MeTAL identified four short-QT and 59 long-QT variants, corresponding to 63 of 111 (56.7%) (**Figure 3*i***). Thus, among class 3 variants, MeTAL raised the proportion identified as dysfunctional from 40.5% to 56.7%. Among the combined class 4 and 5 variants, modeled-QTc Z-scores identified 52 of 70 variants as dysfunctional, raising the proportion from 54.3% to 74.3% (**Figure 3*i***).

**Figure 3.**
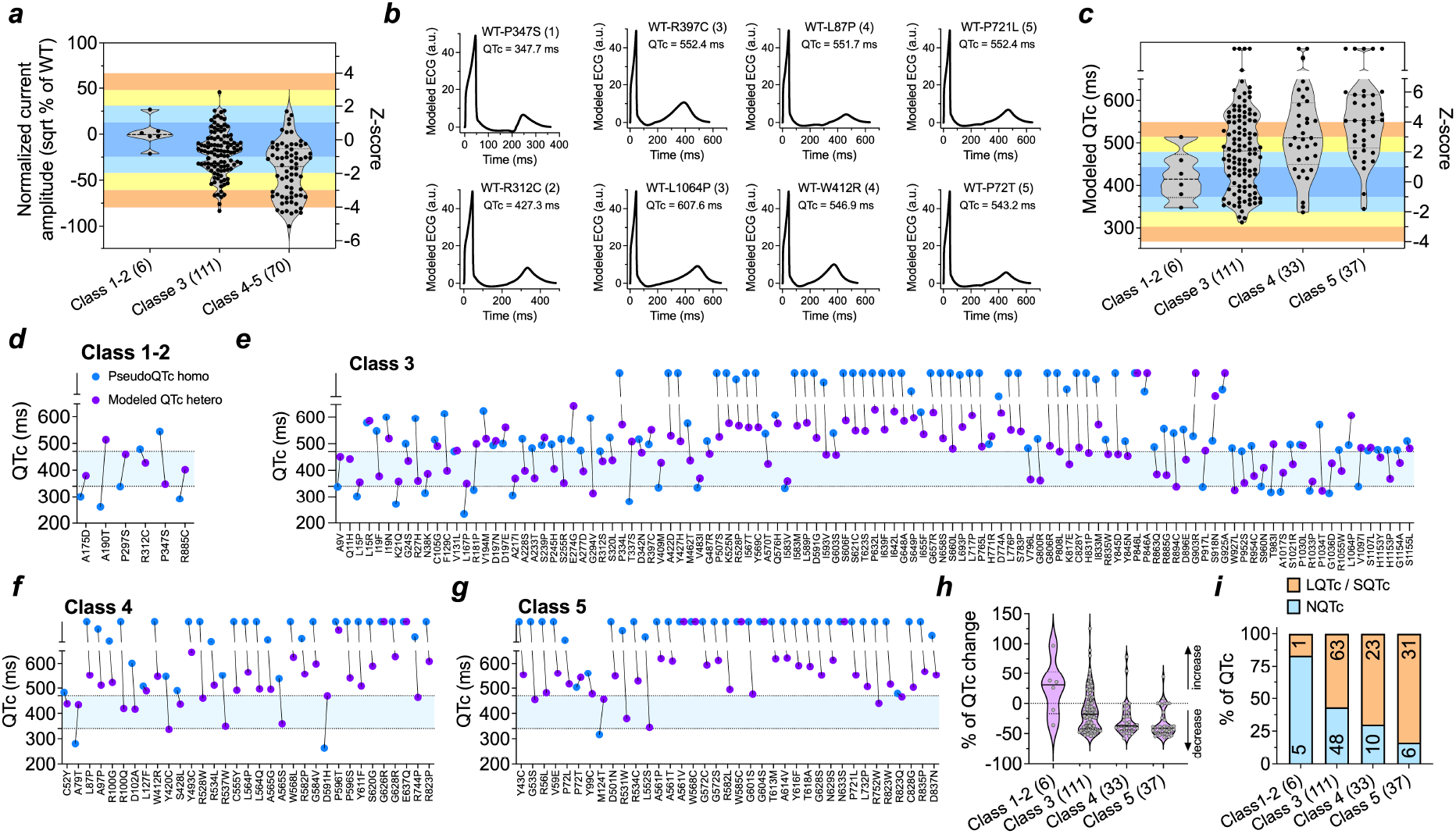
Refinement of predicted QTc phenotypes of dysfunctional *KCNH2* variants under balanced heterozygous conditions. ***a***, Distribution of normalized current amplitude (square root-transformed Z-scores) for 187 *KCNH2* variants under heterozygous conditions. The normal functional range (−2 ≤ Z ≤ 2) was defined using ACMG class 1-2 variants. ***b***, Representative MeTAL-modeled ECG traces for variants from different ACMG classes, illustrating the rescaled ECG morphology and corresponding mean modeled QTc values. ***c***, Distribution of modeled QTc values across ACMG classes. Most class 1-2 variants show QTc values within the normal range, whereas the proportion of prolonged QTc values progressively increases across classes 3, 4, and 5. ***d-g***, Comparison of QTc values predicted from homozygous functional data (pseudoQTc, blue dots) and heterozygous functional data (modeled QTc, purple dots) for ACMG class 1-2 variants (***d***), class 3 variants (***e***), class 4 variants (***f***), and class 5 variants (***g***). Heterozygous modeling markedly reduces the extreme QTc values observed under homozygous conditions, resulting in a narrower QTc distribution and a shift toward less severe phenotypes while preserving the overall class-dependent trend. ***h***, Relative change in QTc between homozygous (pseudoQTc) and heterozygous (modeled QTc) conditions. Class 1-2 variants show minimal changes in QTc and remain within the normal range, whereas class 4-5 variants show substantial QTc reductions that nevertheless frequently remain above the pathological threshold. ***i***, Proportion of variants classified in heterozygous condition as normal QTc (NQTc), long QTc (LQTc), or short QTc (SQTc) across ACMG classes. Most class 1-2 variants are associated with normal QTc values, whereas the proportion of abnormal QT phenotypes progressively increases from class 3 to class 5.

As a control, seven variants displaying normal pseudoQTc values were also examined under heterozygous conditions: one class 2 variant (R328C), four class 3 variants (R92L, R531Q, R887P, and P1093L), one class 4 and one class 5 variant. As expected, all modeled QTc values remained within the normal range (382–438 ms; **Figure S10**, **Table S1**)

### From bench to bedside: correlating the functional assessment of hERG variants by MeTAL with clinical data

Patient-level concordance between MeTAL-modeled QTc and the clinical phenotype decreased across the ACMG spectrum: 82.9% for class 5, 76.2% for class 4 and 45.5% for class 3, a gradient potentially influenced by differences in clinical penetrance. We first investigated whether MeTAL-modeled QTc values obtained under heterozygous conditions correlated with clinical QTc measurements (**Figure 4, Table S1**). Among the 187 *KCNH2* variants investigated under heterozygous conditions, clinical data were available for 108 variants (57.7%), corresponding to 172 individual patient records, with a maximum of seven patients available for a single variant (T613M). For the three ACMG class 1-2 variants, modeled QTc values consistently predicted a normal QTc interval, in agreement with the available clinical data (100% correspondence; **Figure 4*a,b***). Among the 19 ACMG class 4 variants, clinical data were available for 21 patients, including two patients each carrying R100G or Y493C. MeTAL predicted a long QT phenotype for 15 of the 19 variants, a normal QT interval for three variants, and one borderline short QT interval for one variant (**Figure 4*c,d***). At the patient level, 16 of the 21 clinical observations were concordant with the corresponding MeTAL prediction (76.2%; **Figure 4*d***). Among the 37 ACMG class 5 variants, clinical data were available for 36 variants, corresponding to 82 individual patient records (**Figure 4*e,f***). MeTAL predictions agreed with the clinical phenotypes in 68 of 82 cases (82.9% patient-level concordance), including five patients with normal QTc and 63 with long QTc values. Only three variants (R531W, V59E, R835P) showed a clear mismatch between prediction and clinical phenotype. For R531W, MeTAL predicted a normal QTc whereas the available patient had a long QT phenotype. Conversely, for V59E and R835P, MeTAL predicted long QT whereas the available patients had normal QTc values. However, only a single patient was available for each of these variants, precluding a robust assessment of these apparent discrepancies. For the remaining 11 patient-level observations in which the clinical phenotype did not initially match the MeTAL prediction (R752W, M124T, R823Q, G601S, R823W, R534C, D837N, A561T, T613M, A561V, G604S), the prediction was concordant with the phenotype of additional carriers of the same variant. Specifically, concordance was observed in one of two patients for R752W, R823Q, and R534C; in two of three patients for M124T, G601S, and R823W; in three of four patients for D837N; and in four of five patients for A561T, A561V, and G604S. For T613M, concordance was observed in six of seven patients (**Figure 4*e***). These observations support the robustness of MeTAL predictions and suggest that some apparent discrepancies between modeled and clinical QTc values may reflect incomplete clinical penetrance rather than inaccurate prediction of the functional consequences of the variant.

**Figure 4.**
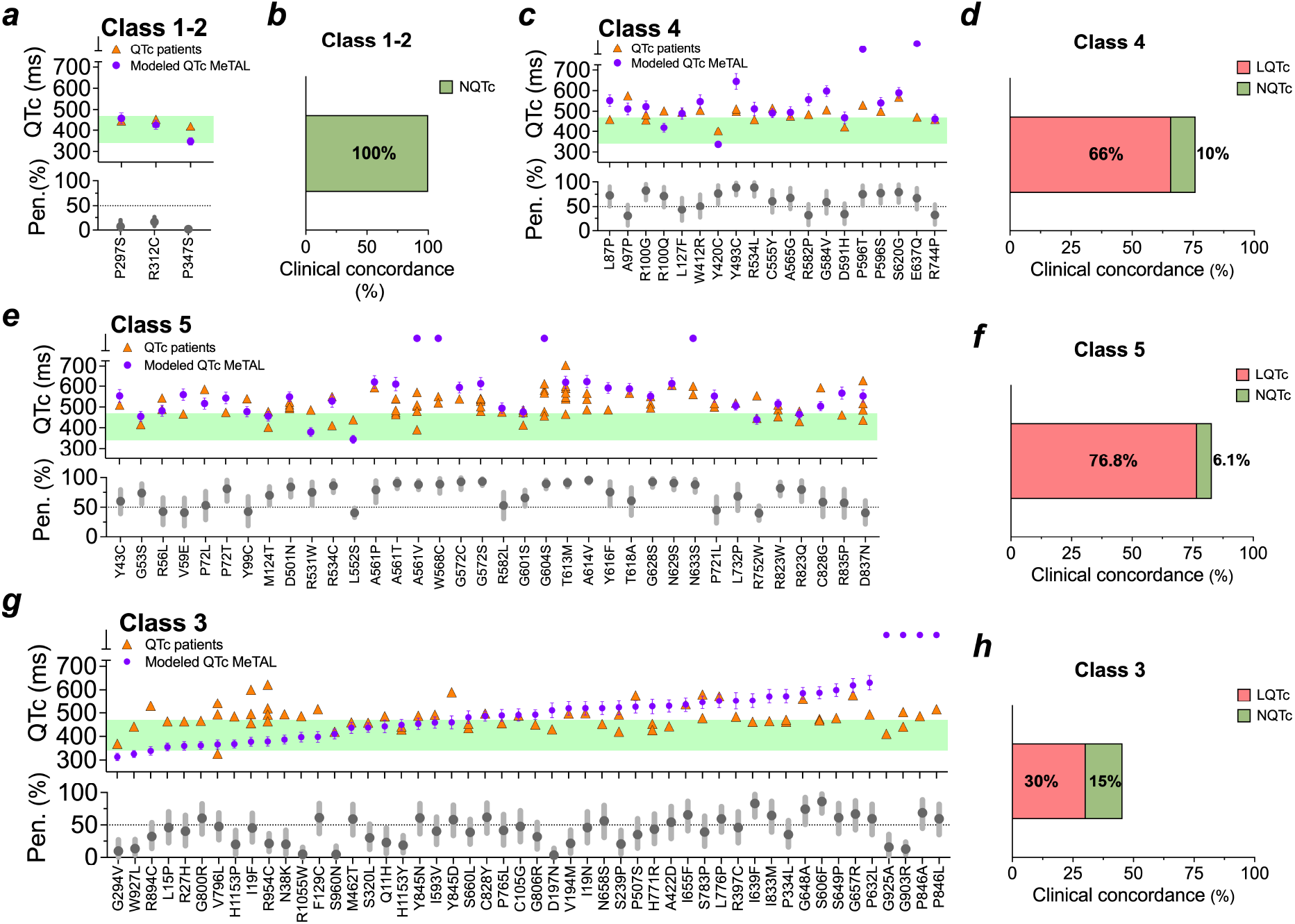
Clinical concordance between MeTAL-modeled QTc values, patient QTc values, and variant-specific penetrance. ***a,c,e,g,*** Comparison of MeTAL-modeled QTc values (mean ± SD) derived from heterozygous functional data (purple circles) with QTc values measured in individual patients carrying the corresponding *KCNH2* variants (orange triangles). The green shaded area indicates the normal QTc range (340-470 ms). Variant-specific LQTS penetrance estimates are shown in the lower panels (gray circles), with error bars representing the 90% credible intervals (CrI). The horizontal dotted line indicates 50% penetrance. Variants are grouped according to ACMG classification (Classes 1-2, 4, 5, and 3). ***b,d,f,h,*** Clinical concordance between the QT phenotype predicted by MeTAL (normal QT (NQtc), or long QT (LQTc)) and the QTc phenotype observed in individual patients. Bars indicate the percentage of patients whose clinical QTc phenotype was concordant with the corresponding MeTAL prediction, relative to the total number of patients evaluated within each ACMG class.

To further investigate discrepancies between MeTAL-predicted and clinical QTc values, we estimated variant-specific LQTS penetrance and its 90% credible interval using available carrier data (**Figure 4*a,c**,e,g*; Table S1**). While ACMG class 1–2 variants exhibited normal MeTAL-predicted QTc values and low penetrance (<50%), class 4 and 5 variants showed predominantly dysfunctional QTc phenotypes with variable penetrance, including unaffected carriers of variants predicted to prolong QTc. Among the 111 VUS characterized under heterozygous conditions, clinical data were available for 50 variants (66 patients), with a patient-level concordance of 45.5% (**Figure 4*g,h***). Notably, nine VUS predicted to have normal QTc values showed concordant clinical phenotypes despite exhibiting functional defects under homozygous conditions, whereas among those predicted to prolong QTc, only 20 of 36 patients (55.6%) exhibited prolonged clinical QTc values. The proportion of variants with penetrance >50% increased across ACMG classes, from 0% for class 1–2 to 38% for class 3, 72.2% for class 4, and 77.8% for class 5, suggesting that incomplete penetrance may partly explain the lower clinical concordance observed among VUS.

Among the 303 hERG variants characterized, 116 exhibited normal pseudoQTc values and were therefore not further investigated under heterozygous conditions (**Figure 2*k,l***). Clinical data were available for 26 of the 101 ACMG class 3–5 variants in this group, with 77% of patient QTc measurements falling within the normal range (**Figure S11, Table S1**). Notably, variants such as V115M, R892H, D712N, and E788K were associated with both normal and prolonged QTc values among different carriers, illustrating clinical heterogeneity (**Figure S11*a-c***). Although pseudoQTc values derived from homozygous functional data cannot be directly compared with clinical QTc measurements from heterozygous carriers, these observations support their use as a screening criterion while acknowledging the possibility of overlooking clinically relevant variants.

### Calibration of MeTAL-modeled QTc for functional evidence assignment

To establish the strength of functional evidence according to the ClinGen Sequence Variant Interpretation (SVI) recommendations ^10^, MeTAL-modeled QTc values were calibrated using variants with definitive ACMG classifications. The reference dataset comprised 58 variants, including 20 benign/likely benign (B/LB; ACMG classes 1-2) and 38 pathogenic variants (ACMG class 5) (**Figure 5*a***). Variants with normal functional phenotypes under homozygous conditions were represented by their pseudo-QTc values, whereas those with abnormal phenotypes were represented by their modeled-QTc values. This approach provided a common phenotypic readout for calibration across both experimental conditions.

**Figure 5.**
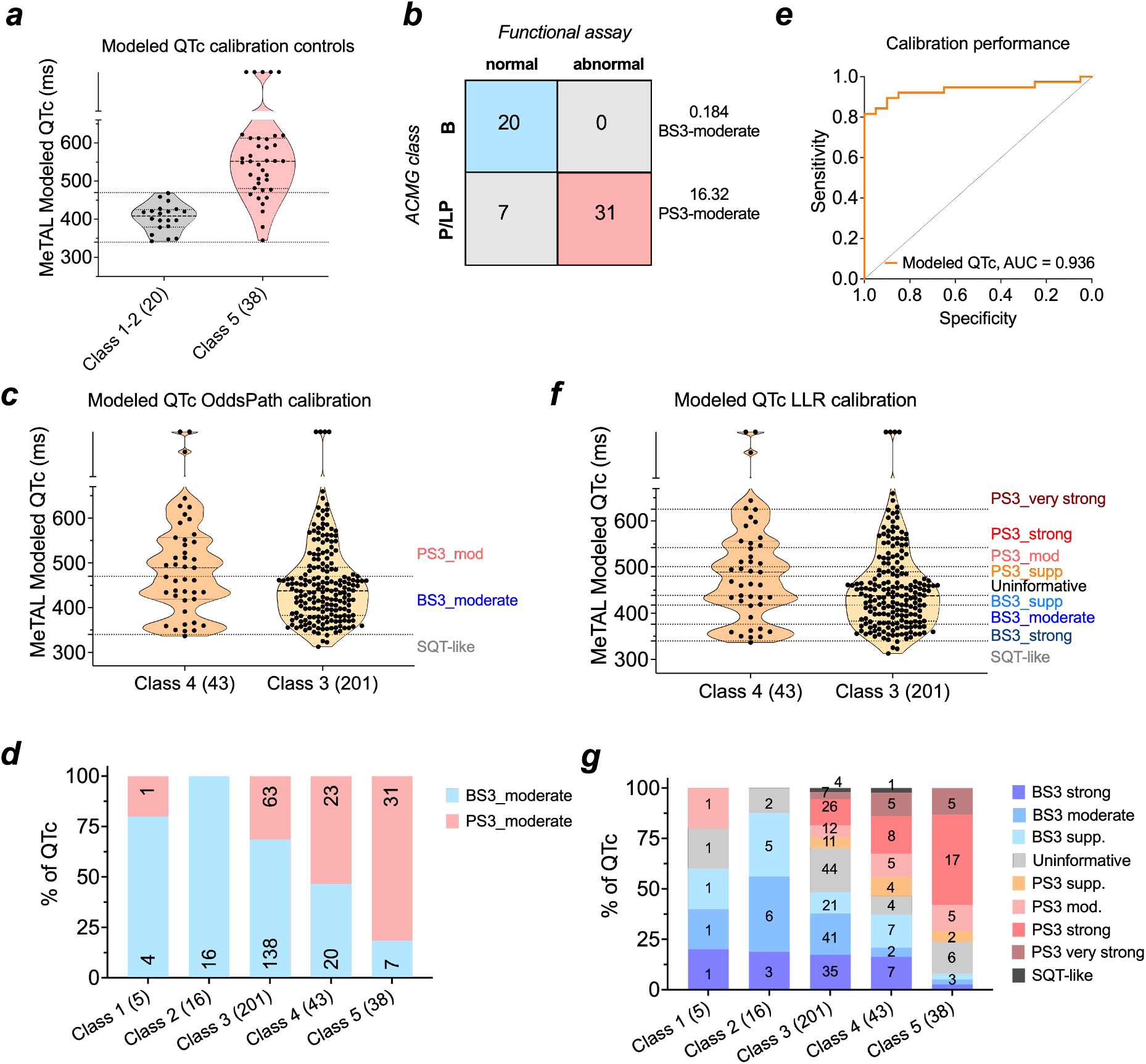
Calibration of MeTAL-modeled QTc for functional evidence assignment using categorical OddsPath and continuous likelihood-ratio approaches. ***a,*** Distribution of MeTAL-modeled QTc values among the reference variants used for calibration. Benign/likely benign variants (B/LB; ACMG classes 1-2, n = 20) and pathogenic variants (ACMG class 5, n = 38) constituted the benign and pathogenic reference groups, respectively. Dotted lines indicate the modeled QTc thresholds defining the normal range (340–470 ms). ***b***, Calibration of categorical functional evidence using the OddsPath framework. Among the reference variants, all 20 B/LB variants showed a normal modeled QTc, whereas 31 of 38 pathogenic variants showed an abnormal modeled QTc. Following application of the ClinGen add-one correction, OddsPath values were 0.184 for a normal functional result and 16.32 for an abnormal functional result, corresponding to BS3_moderate and PS3_moderate evidence, respectively. ***c***, Application of the categorical OddsPath calibration to ACMG class 4 variants (n = 43) and VUS (ACMG class 3, n = 201), which were not included in the calibration dataset. Horizontal dotted lines indicate the modeled QTc thresholds used to assign PS3_moderate or BS3_moderate evidence. Variants with modeled QTc <340 ms were considered separately as short-QT-like phenotypes. ***d***, Distribution of functional evidence strength across the five ACMG classes using the categorical OddsPath calibration. Bars represent the percentage of variants within each ACMG class assigned to each functional evidence category; numbers within bars indicate the corresponding number of variants. ***e***, Receiver operating characteristic (ROC) analysis of MeTAL-modeled QTc to discriminate between B/LB and pathogenic reference variants. MeTAL-modeled QTc showed an area under the curve (AUC) of 0.936; the diagonal line represents discrimination expected by chance. ***f***, Application of the continuous likelihood-ratio (LLR) calibration to ACMG class 4 variants and VUS. Horizontal dotted lines indicate LLR-derived thresholds corresponding to PS3_supporting, PS3_moderate, PS3_strong and PS3_very strong, and to BS3_supporting, BS3_moderate and BS3_strong evidence. Values not reaching the supporting threshold in either direction were considered functionally uninformative. Because marked QT shortening may represent a pathogenic GoF phenotype, variants with modeled QTc <340 ms were not assigned benign evidence despite falling beyond the benign LLR thresholds and were instead classified separately as short-QT-like. ***g***, Distribution of functional evidence strength across the five ACMG classes using the continuous LLR calibration. Bars represent the percentage of variants within each ACMG class assigned to each functional evidence category; numbers within bars indicate the corresponding number of variants.

We first calibrated MeTAL using the categorical OddsPath framework recommended by ClinGen ^10^. Functional results were classified as normal or abnormal according to the modeled QTc phenotype. All 20 B/LB reference variants displayed a normal modeled QTc, whereas 31 of 38 pathogenic reference variants displayed an abnormal modeled QTc. Following application of the conservative add-one correction, normal and abnormal functional readouts yielded OddsPath values of 0.184 and 16.32, respectively, corresponding to BS3_moderate and PS3_moderate evidence (**Figure 5*b*, Table S3**). Applying this categorical calibration to the complete dataset revealed a progressive enrichment of PS3_moderate evidence with increasing ACMG pathogenicity class (**Figure 5*c,d*, Table S1**). Among the 21 ACMG class 1-2 variants, 20 received BS3_moderate evidence, whereas A190T displayed an abnormal functional phenotype and received PS3_moderate evidence. Notably, the conflicting interpretations currently reported for A190T in ClinVar are consistent with its exclusion from the benign calibration group. Among the 43 ACMG class 4 variants, 20 (46.5%) received BS3_moderate and 23 (53.5%) received PS3_moderate evidence. Among the 38 ACMG class 5 variants, 31 (81.6%) received PS3_moderate, whereas 7 (18.4%) received BS3_moderate evidence. Finally, among the 201 VUS (ACMG class 3), 138 (68.7%) received BS3_moderate and 63 (31.3%) PS3_moderate evidence (**Figure 5*c,d***). The variant-level distribution of normal and abnormal MeTAL-modeled QTc phenotypes underlying these OddsPath evidence assignments is shown (**Figure S12)**. Thus, categorical OddsPath calibration provided functional evidence for all variants but, by design, assigned the same evidence strength to variants with substantially different degrees of functional alteration.

To exploit the continuous nature of MeTAL-modeled QTc, we next developed a likelihood-ratio (LLR) calibration using the same reference dataset. MeTAL-modeled QTc showed high discriminatory performance between B/LB and pathogenic reference variants, with an area under the ROC curve of 0.936 (**Figure 5*e***). The calibrated continuous LLR model translated modeled QTc values into graded strengths of evidence for pathogenicity or benignity, corresponding to PS3_supporting, PS3_moderate, PS3_strong and PS3_very strong, and to BS3_supporting, BS3_moderate and BS3_strong evidence. Modeled QTc values that did not reach the supporting threshold in either direction were considered functionally uninformative. The continuous calibration substantially refined functional evidence assignment compared with the categorical OddsPath approach (**Figure 5*f,g*, Table S1**). Among the 43 ACMG class 4 variants, 22 (51.2%) provided LLR-based evidence toward LQT pathogenicity, 13 of them reaching PS3_strong or PS3_very strong, whereas 16 (37.2%) provided evidence toward benignity and 4 (9.3%) were functionally uninformative (**Table S1**). One additional class 4 variant displayed an SQT-like modeled phenotype (<340 ms) and was therefore considered separately. Application of the continuous MeTAL-QTc calibration to the 201 VUS provided LLR-based evidence toward either pathogenicity or benignity for 153 variants (76.1%) (**Figure 5*f,g*, Table S1**). Fifty-six variants (27.9%) provided evidence toward pathogenicity, 33 of them reaching PS3_strong or PS3_very strong, whereas 97 (48.3%) provided evidence toward benignity, 76 of them reaching BS3_moderate or BS3_strong (**Table S1**). Forty-four variants (21.9%) fell within the LLR-uninformative interval and therefore did not reach the supporting evidence threshold in either direction. Four additional VUS (2.0%) displayed an SQT-like modeled phenotype (<340 ms), bringing the total to five SQT-like variants across ACMG classes 3 and 4. Because marked QT shortening may represent a pathogenic GoF phenotype of *KCNH2*, these variants were not assigned benign evidence despite falling on the low-QTc side of the calibration. The present LLR model was calibrated along the LoF/long-QT spectrum and therefore does not quantify pathogenic evidence in the short-QT direction. These variants were consequently considered separately from both the benign and LLR-uninformative categories. Overall, whereas categorical OddsPath calibration assigned a uniform moderate level of functional evidence based solely on a normal *versus* abnormal phenotype, continuous LLR calibration captured the magnitude of the functional alteration, distinguishing variants providing strong or very strong functional evidence from those providing only supporting evidence or yielding functionally uninformative results. This substantially increased the resolution of functional evidence assignment, particularly among VUS.

### Calibrated functional evidence enables reclassification of KCNH2 VUS

Finally, we assessed whether the calibrated functional evidence generated in this study could contribute to resolving the 201 *KCNH2* VUS according to the ACMG/AMP framework ^9^. To quantitatively integrate the different lines of evidence, ACMG/AMP evidence strengths were converted into the Bayesian point-based framework developed by Tavtigian and colleagues ^23,24^. Pre-existing PS3 or BS3 criteria were removed before calculation of the baseline score to avoid double counting functional evidence, and the newly generated MeTAL-modeled QTc evidence was subsequently incorporated using either categorical OddsPath or continuous likelihood-ratio (LLR) calibration^10^ (**Tables S5** and **S6**). Notably, pre-existing functional evidence was available for only 2 of the 201 VUS; our systematic characterization therefore provided new functional evidence for 199 variants (99.0%). Using categorical OddsPath calibration, 63 of the 201 VUS (31.3%) fulfilled the criteria for PS3_moderate, whereas 138 (68.7%) met the criteria for BS3_moderate (**Figure 5*c,d***). Within the quantitative framework, moderate evidence contributes +2 points toward pathogenicity or −2 points toward benignity ^23,24^, allowing the calibrated functional evidence to be integrated with the remaining ACMG/AMP evidence originally associated with each variant ^9^. Incorporation of OddsPath-calibrated functional evidence reclassified 58 of the 63 variants assigned PS3_moderate (92.1%) from VUS to likely pathogenic, corresponding to 28.9% of the 201 VUS, whereas the remaining five retained their VUS classification (**Figure 6*a,b***). Conversely, all 138 variants assigned BS3_moderate remained classified as VUS, despite a two-point shift toward benignity. Overall, 143 of the 201 variants (71.1%) therefore remained classified as VUS. These results illustrate the additive nature of the Bayesian point-based framework, in which the impact of functional evidence depends both on its empirically supported strength and on the evidence already available for each variant ^10,23,24^. This asymmetric effect largely reflected the distribution of pre-existing non-functional evidence. Most variants were already positioned close to the likely pathogenic threshold, with 147/201 (73.1%) having a baseline score of +5 before incorporation of the newly calibrated functional evidence. Consequently, the addition of PS3_moderate (+2 points) was sufficient to move most functionally abnormal variants beyond the ≥6-point likely pathogenic threshold, whereas BS3_moderate (−2 points) was generally insufficient to reach the ≤−1-point likely benign threshold (**Figure 6*a***). Thus, the differential impact of PS3_moderate and BS3_moderate on VUS resolution reflects their integration with the existing evidence profile of each variant, rather than differences in their numerical weight.

**Figure 6.**
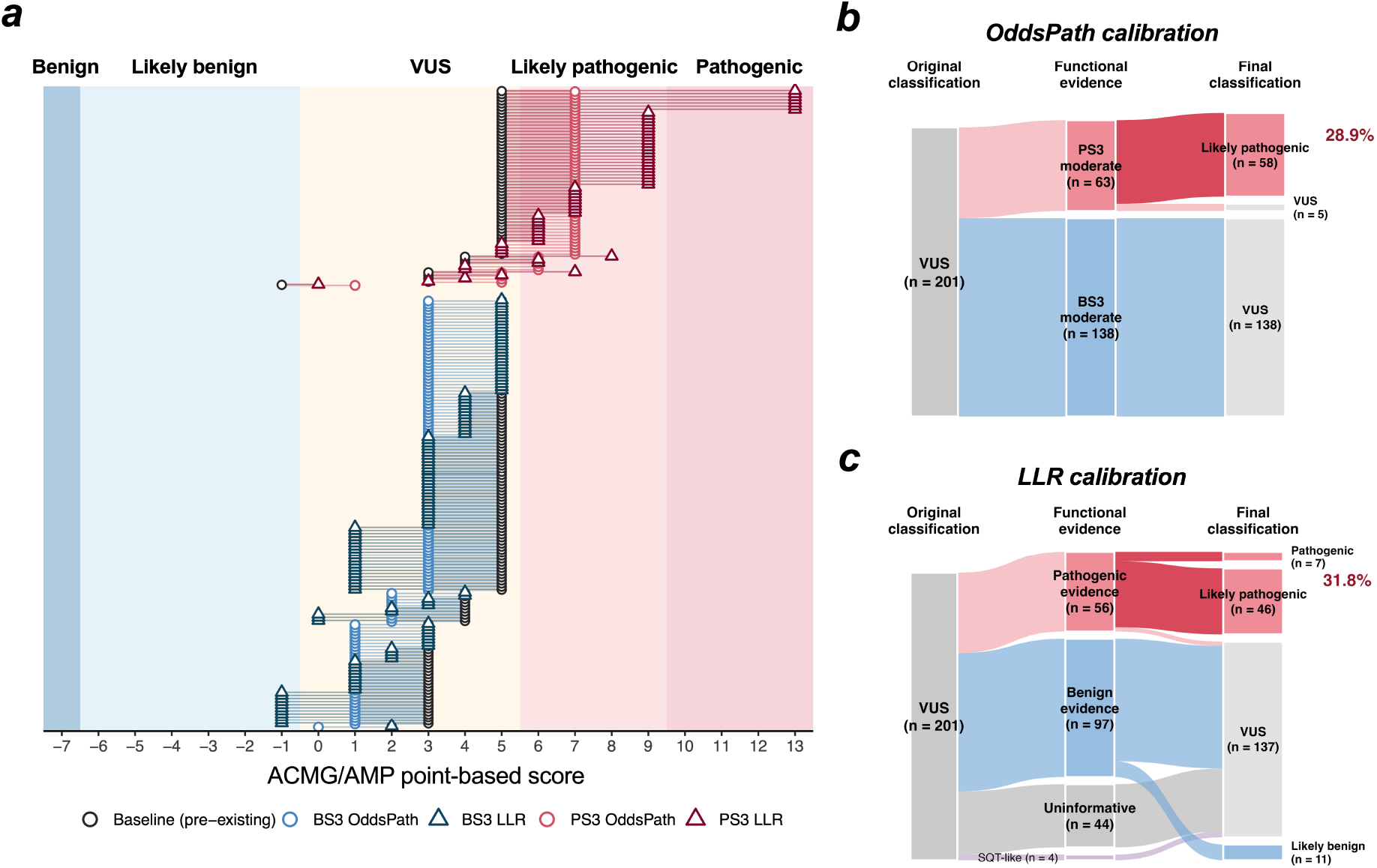
Calibrated functional evidence enables reclassification of *KCNH2* variants of uncertain significance. ** *a*,** ACMG/AMP point-based scores for the 201 *KCNH2* variants initially classified as variants of uncertain significance (VUS), before and after incorporation of functional evidence calibrated using categorical OddsPath or continuous likelihood-ratio (LLR) approaches. Open black circles indicate baseline scores derived from pre-existing ACMG/AMP criteria, while colored circles and triangles represent final scores obtained using OddsPath and LLR calibration, respectively. Red and blue indicate evidence supporting pathogenicity (PS3) and benignity (BS3), with lighter shades representing OddsPath and darker shades representing LLR. Lines connect baseline and final scores for each variant. Shaded backgrounds indicate the ACMG/AMP classification categories according to the Bayesian point-based framework: benign (≤−7), likely benign (−6 to −1), VUS (0–5), likely pathogenic (6–9), and pathogenic (≥10). ***b*,** Sankey diagram illustrating variant reclassification following categorical OddsPath calibration. Among the 201 VUS, 63 received PS3_moderate evidence (+2 points), of which 58 were reclassified as likely pathogenic and five remained VUS. The remaining 138 variants received BS3_moderate evidence (−2 points) and retained their VUS classification. Overall, 58 variants (28.9%) were reclassified. ***c*,** Sankey diagram illustrating variant reclassification following continuous LLR calibration. Among the 56 variants receiving evidence supporting pathogenicity, seven were reclassified as pathogenic, 46 as likely pathogenic, and three remained VUS. Of the 97 variants receiving evidence supporting benignity, 11 were reclassified as likely benign and 86 remained VUS. An additional 44 variants received uninformative evidence, while four variants displaying SQT-like phenotypes were not assigned LLR-based evidence because the calibration was not applicable to QT shortening. Overall, LLR calibration enabled the reclassification of 64 variants (31.8%), including seven pathogenic, 46 likely pathogenic, and 11 likely benign classifications.

We next investigated whether the graded functional evidence provided by continuous LLR calibration could overcome the limitations of assigning uniformly moderate evidence and enable more definitive ACMG/AMP classifications (**Figure 6*a,c*, Table S6**). This approach enabled the reclassification of 64 of the 201 VUS (31.8%), including seven as pathogenic, 46 as likely pathogenic, and 11 as likely benign. Among the 56 variants receiving evidence supporting pathogenicity, 53 were reclassified (seven as pathogenic and 46 as likely pathogenic), while three remained VUS. Notably, all seven variants receiving very strong pathogenic evidence reached the pathogenic classification, and all 26 variants receiving strong pathogenic evidence were reclassified as likely pathogenic. Conversely, among the 97 variants receiving evidence supporting benignity, 11 were reclassified as likely benign, all supported by strong benign evidence, while 86 remained VUS. An additional 44 variants received uninformative evidence, while 4 variants displaying SQT-like phenotypes were not assigned LLR-based evidence because the calibration was not applicable to QT shortening. These 48 variants retained their baseline ACMG/AMP classification. Although the overall reclassification rate was only modestly higher with LLR than with OddsPath calibration (31.8% versus 28.9%), the distribution of final classifications differed substantially (**Figure 6*b,c***). Whereas OddsPath calibration exclusively reclassified variants as likely pathogenic, continuous LLR calibration enabled reclassification toward both pathogenicity and benignity, including seven pathogenic and 11 likely benign classifications. This demonstrates the added value of quantitatively graded functional evidence in refining ACMG/AMP classification outcomes beyond those achievable with uniformly moderate evidence.

Importantly, the non-functional ACMG/AMP criteria were incorporated at their originally assigned strengths and were not independently reassessed in this study ^20^. Therefore, the resulting classifications reflect the impact of adding calibrated functional evidence to existing variant assessments rather than comprehensive *de novo* ACMG/AMP classifications.

### Cross-platform benchmarking establishes the added value of MeTAL for functional characterization of KCNH2 variants

An important consideration for the clinical use of functional data is their reproducibility across experimental platforms. Comparison of the 101 variants shared with O’Neill et al. ^11^ revealed several advantages of the MeTAL-based approach (**Figure 7**). First, MeTAL identified repolarization abnormalities in variants with normal current amplitudes that may be missed by approaches relying primarily on trafficking and conductance measurements. Indeed, 6 of 7 variants with normal current amplitudes but prolonged MeTAL-predicted QTc (G903R, R582P, R397C, V194M, A190T, T983I, P917L) exhibited significant hyperpolarizing shifts in voltage-dependent inactivation (−6.5 to −15.6 mV), highlighting the contribution of gating properties to functional interpretation (**Figure 7*a***). Conversely, 14 variants with reduced (n=10) or increased (n=4) current amplitudes in O’Neill et al. had normal MeTAL-predicted QTc. Notably, none of the 10 loss-of-current variants (R752W, F129C, L552S, I593V, G53S, Y845D, R744P, R531W, R823Q, R863Q) showed significantly reduced current amplitudes under our heterozygous conditions. This discrepancy may reflect expression-dose-dependent effects amplified by the preferential expression of the variant over WT hERG in the pIRES system used in previous studies ^25^. By contrast, our co-electroporation strategy, using independent WT and variant plasmids at a 1:1 ratio, was designed to approximate balanced allelic expression in the heterozygous state ^26^, a critical consideration when assessing dominant-negative effects (**Figure 7b**). The four gain-of-current variants (H1153Y, P297S, M124T, R528W) displayed only modest changes in activation and deactivation kinetics, without significant alterations in current amplitude or inactivation, insufficient to shorten predicted QTc (**Figure 7a**).

**Figure 7.**
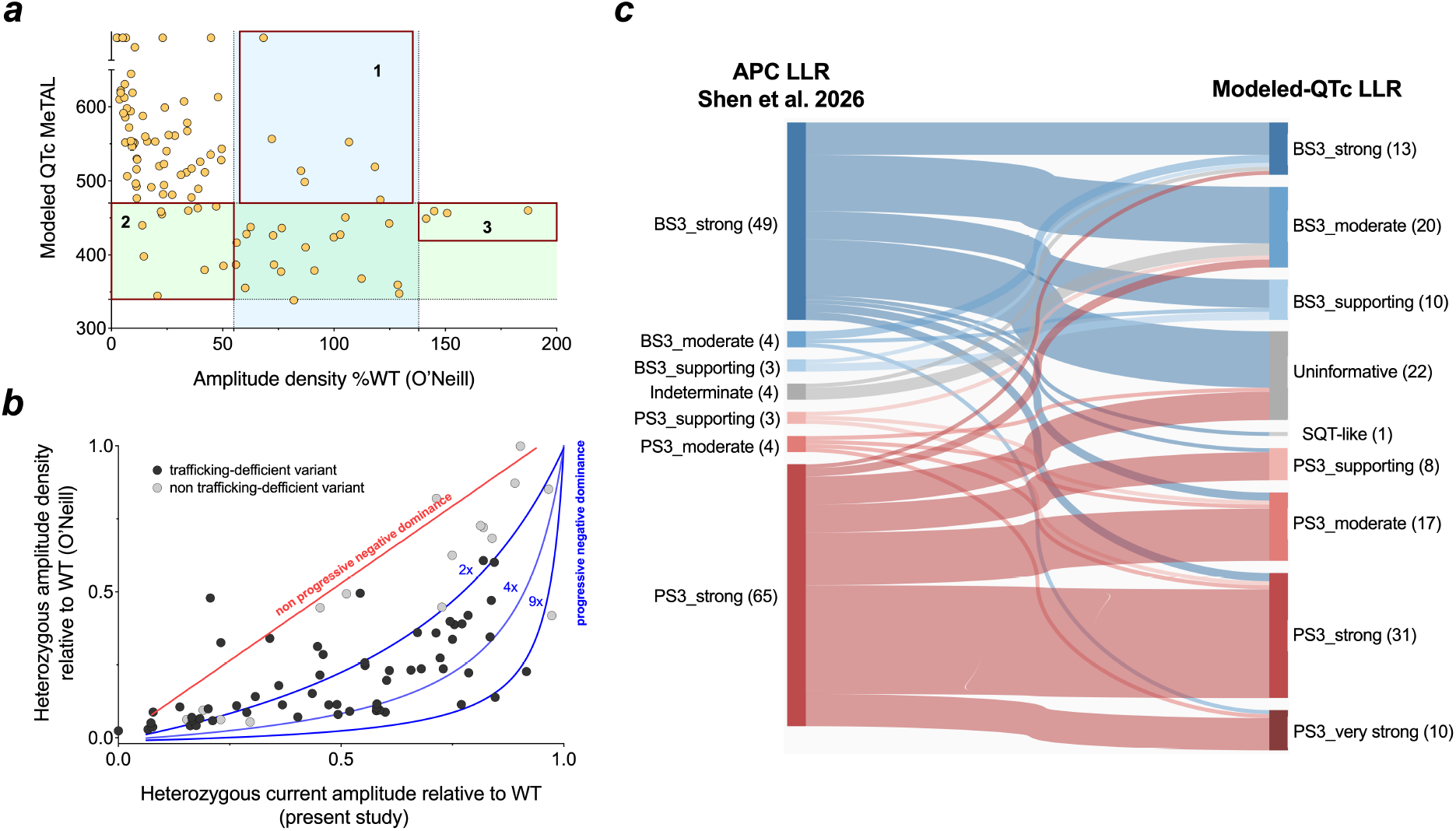
Cross-platform comparison highlights the impact of multidimensional functional assessment and heterozygous expression models on *KCNH2* variant interpretation. ***a,*** Comparison of current amplitude and MeTAL-predicted QTc in 101 shared *KCNH2* variants. Scatter plot comparing peak tail current amplitude (% WT) reported by O’Neill et al. (2024) with MeTAL-predicted QTc values obtained in the present study. Each dot represents an individual *KCNH2* variant (n = 101). Vertical dotted lines indicate the normal current amplitude range (55–138% WT; Z-scores between −2 and +2), as established by Thomson et al. ^16^, while horizontal dotted lines indicate the normal QTc reference range. Three groups of discordant variants are highlighted: (1) 7 variants with normal current amplitude (55–138% WT) but prolonged predicted QTc; (2); 10 variants with reduced current amplitude (<55% WT) but normal predicted QTc and (3) 4 variants with increased current amplitude (>138% WT) but normal predicted QTc. These discrepancies illustrate how alterations in channel biophysical properties may either compensate for changes in current amplitude or impair repolarization despite preserved current amplitude, while also highlighting the potential influence of experimental expression strategies, particularly the preferential expression of variant over WT subunits in the pIRES system, which may exacerbate apparent loss of function. ***b,*** Influence of heterozygous expression strategies on hERG current amplitude. Comparison of heterozygous current amplitudes relative to WT obtained using the bicistronic pIRES system (O’Neill et al., 2024; y-axis) and independent and balanced WT and variant plasmid co-expression (present study; x-axis). Trafficking-deficient variants (black dots) generally exhibit lower current amplitudes in the pIRES system, consistent with the amplification of progressive dominant-negative effects resulting from preferential variant expression. Blue curves represent theoretical models of progressive dominant-negative effects assuming 2-, 4-, or 9-fold higher variant than WT expression. Non-trafficking-deficient variants (grey dots) generally exhibit more comparable current amplitudes between expression systems, consistent with functional effects less sensitive to the relative proportion of variant subunits (red line). See Rio et al. ^30^ for details. ***c,*** Concordance between functional evidence assignments derived from APC-based LLR ^27^ and MeTAL modeled-QTc LLR (present study) for 132 shared *KCNH2* variants. Band widths are proportional to the number of variants transitioning between evidence categories, from Shen et al. (left) to MeTAL (right). Overall concordance in evidence direction was 72.0% (95/132), including 35 BS3→BS3 and 60 PS3→PS3 variants, while 10 variants (7.6%) received opposing evidence and 27 (20.5%) were uninformative in at least one approach.

We next compared APC-derived LLR evidence reported by Shen et al. ^27^ with MeTAL modeled-QTc LLRs for 132 shared variants (**Figure 7*c***). Functional evidence was concordant in 72.0% (95/132), while 10 variants (7.6%) received opposing PS3/BS3 assignments and 27 (20.5%) were indeterminate or uninformative in at least one approach. Notably, these discrepancies partly recapitulated those observed with O’Neill et al. Six variants classified as BS3 by Shen et al. (G903R, A190T, V194M, R397C, R582P, and T983I) received PS3 evidence from MeTAL, despite normal current amplitudes in O’Neill et al. ; most exhibited altered voltage-dependent inactivation. Conversely, four variants classified as PS3 by Shen et al. (F129C, R531W, L552S, and R863Q) received BS3 evidence from MeTAL. Although these variants showed reduced current amplitudes in O’Neill et al., their current amplitudes were preserved under our heterozygous conditions, suggesting that differences in expression systems and the integration of multiple biophysical parameters contribute to the divergent classifications. Together, these comparisons illustrate how MeTAL can refine functional interpretation by integrating multiple hERG biophysical properties and their combined impact on ventricular repolarization, including gating abnormalities that may occur independently of, or compensate for, changes in current density ^28,29^. This is particularly relevant for hERG, where altered gating can substantially modify I_Kr_ contribution to repolarization despite relatively preserved current amplitude.

Finally, the observed expression-system bias does not apply to trafficking, which was measured in the homozygous state in both studies. Nevertheless, agreement across the 73 shared variants remained incomplete (Pearson r = 0.804, Spearman ρ = 0.685, both p < 0.0001; **Figure S3*f***). Moreover, eight variants classified as having near-absent trafficking by MAVE retained substantial membrane expression in our assay, as supported by measurable hERG currents (**Figure S3*g***). These discrepancies, likely reflecting methodological differences – namely on tag identity / location – further emphasize the value of orthogonal experimental validation.

### A web-based KCNH2 variant database for researchers and clinicians

To organize the extensive dataset generated during this project comprising the characterization of 303 French *KCNH2* variants, and approximately 130,000 recorded and analyzed cells, we developed an interactive web-based database (https://hammer.bird.glicid.fr). The database integrates clinical information and aggregated functional data for each variant, facilitating data interrogation and interpretation by clinicians. For every variant, users can access experimentally measured electrophysiological parameters, SPARC-predicted pathogenicity score ^20^, MeTAL-predicted QT phenotype ^17^, a modeled ECG, and aggregated clinical data. It also reports OddsPath values and likelihood-ratio calibration scores, which quantify the strength of functional evidence supporting pathogenicity or benignity. Through the web interface, users can search for individual variants or filter them according to ACMG classification, each variant being presented on a dedicated page that integrates these data. To further place these findings in a structural context, the database incorporates a three-dimensional model of the hERG channel. Users can thus visualize the location of each variant within the channel alongside its functional and clinical data.

## Discussion

The increasing availability of clinical sequencing has revealed a large and growing number of *KCNH2* variants, while functional characterization has remained comparatively limited. Large-scale functional studies have begun to address this gap, demonstrating the feasibility of multiplexed trafficking assays and high-throughput electrophysiology for generating functional evidence at scale. Early studies established the value of massively parallel assays for identifying trafficking-deficient *KCNH2* variants, initially focusing on selected regions of the channel ^15,31^. More recently, automated patch-clamp approaches have enabled direct measurement of channel function for hundreds of variants, providing a complementary readout to multiplexed trafficking assays and demonstrating their potential for clinical variant interpretation ^11,14,32–34^. The recent study by O’Neill et al. ^11^ further demonstrated the value of combining large-scale functional measurements with clinical phenotyping and penetrance estimates for *KCNH2*-LQT2 variant classification and risk stratification.

A major strength of the present study is its population-based, national-scale design. Rather than selecting variants according to their location within the channel or their prior classification, we systematically characterized *KCNH2* variants identified through the French clinical network, including the complete set of VUS collected within this framework. The relevance of such a comprehensive national strategy extends beyond the French cohort, as variants identified in France account for approximately 10% of *KCNH2* variants currently reported in ClinVar, highlighting the substantial contribution of the French diagnostic network to the global landscape of *KCNH2* variation. This strategy provided functional evidence for all 201 VUS and, more broadly, established a standardized functional framework for 303 clinically identified variants.

Beyond its scale, the principal advance of this study is the integration of standardized multiparametric electrophysiology, balanced (50%WT/50%variant) heterozygous functional assessment, MeTAL-based transformation of channel-level dysfunction into modeled repolarization phenotypes, and formal calibration of PS3/BS3 evidence. This combination captures defects not detected by current amplitude alone, evaluates them in the genetic context of carriers, and translates complex and potentially opposing biophysical effects into clinically interpretable evidence. To facilitate translation of these findings into clinical practice, we developed HAMMER, an open-access database integrating functional, computational, classification, and available clinical data for each variant. By linking systematic experimental characterization with computational modeling and patient phenotypes, this approach provides a framework for connecting national genetic diagnostics with standardized functional interpretation. More broadly, it illustrates how a clinically ascertained national cohort can serve as a framework for systematic functional genomics of an inherited ion-channel disorder.

Comparisons with previously published functional datasets highlight the importance of integrating multiple biophysical properties when interpreting *KCNH2* variants. Discrepancies across platforms may arise from differences in experimental conditions, particularly WT-to-variant expression ratios, but also reflect the limitations of approaches relying primarily on current amplitude or trafficking measurements. Indeed, alterations in gating properties may reinforce or compensate for changes in current density, ultimately modifying their impact on ventricular repolarization. By integrating these combined effects into a modeled QTc phenotype, MeTAL provides a complementary framework for refining functional evidence and emphasizes the importance of orthogonal validation in variant interpretation.

Variants were evaluated in two genotype contexts. Homozygous expression measures the intrinsic capacity of a variant to perturb hERG function, whereas heterozygous expression reproduces the molecular context of LQT2 carriers and tests whether the defect persists in the presence of WT channels, where dominant-negative effects and allele dosage operate. The two conditions were not interchangeable in our dataset: 69 variants showed a marked defect under homozygous conditions while retaining near-normal function in the heterozygous state, consistent with haploinsufficiency rather than dominant-negative suppression. Extending this to patients, MeTAL predictions showed high patient-level concordance for class 5 variants (82.9%), and most apparent mismatches occurred for variants for which other carriers displayed the predicted phenotype, consistent with the incomplete penetrance of LQT2 and with recent large-scale penetrance analyses ^11,22^. A normal QTc in a carrier therefore does not contradict a pathogenic functional effect, nor does an abnormal QTc establish that the *KCNH2* variant is solely responsible: patient-level measurements contextualize functional predictions rather than serving as a gold standard against which they are scored. Penetrance itself increased with ACMG pathogenicity, from exclusively low estimates among class 1-2 variants to >50% penetrance for 72.2% and 77.8% of class 4 and class 5 variants, respectively, yet varied substantially within classes. ACMG classification estimates the probability that a variant is disease-causing; penetrance estimates the probability that carriers express the phenotype. The divergence is most visible among VUS, for which patient-level concordance was lower and clinical heterogeneity greater: an abnormal functional phenotype associated with a normal QTc in an individual patient cannot, by itself, exclude a pathogenic contribution of the variant. Interpreted together, MeTAL and penetrance provide a framework for functionally stratifying VUS beyond their current class 3 designation and for placing individual patient QTc values within the phenotypic spectrum associated with each variant.

Translating functional phenotypes into clinically usable evidence requires not only defining whether a variant is abnormal, but also quantifying the strength of that evidence. The ClinGen Sequence Variant Interpretation Working Group specifically recommends calibration of well-established functional assays using known pathogenic and benign controls to determine the appropriate strength of PS3/BS3 evidence ^10^. Calibration of MeTAL-modeled QTc according to this framework provided moderate PS3 or BS3 evidence across the complete dataset. However, binary OddsPath classification necessarily assigns equivalent evidence strength to variants with substantially different degrees of functional alteration. Taking advantage of the continuous nature of modeled QTc, our LLR calibration captured this magnitude and showed high discrimination between established benign and pathogenic variants (AUC = 0.936). Importantly, this approach provided graded evidence toward pathogenicity or benignity for 76.1% of VUS, ranging from supporting to very strong evidence, while explicitly identifying variants for which the functional phenotype remained uninformative. The clinical impact of this calibrated evidence is most apparent in the VUS category. Prior functional evidence was available for only 2 of the 201 variants analyzed here. By generating calibrated functional evidence across nearly the entire VUS dataset, our approach enabled 58 variants (28.9%) to be reclassified as likely pathogenic using categorical OddsPath calibration. Importantly, continuous LLR calibration extended this contribution by enabling the reclassification of 64 variants (31.8%), including seven as pathogenic, 46 as likely pathogenic, and 11 as likely benign. Although the overall reclassification rates were comparable, LLR calibration provided greater resolution of functional evidence strength, allowing variants to reach more definitive classifications in both pathogenic and benign directions. This reclassification did not result from functional evidence acting in isolation, but from its quantitative integration with the pre-existing evidence supporting each variant: most reclassified variants were already positioned close to the likely pathogenic threshold and crossed this boundary upon addition of PS3_moderate evidence.

Conversely, although BS3_moderate shifted functionally normal variants toward benignity, it was insufficient to reclassify them because their existing evidence profiles remained within the VUS range. The ability of LLR calibration to assign stronger evidence in either direction therefore partially overcame this limitation, particularly for variants receiving strong benign or very strong pathogenic evidence. These findings illustrate how systematically generated and quantitatively calibrated functional data can help resolve VUS while preserving the evidence-based logic of the ACMG/AMP framework ^10,23,24^.

Several limitations should be acknowledged. The expression system, although standardized and highly scalable, does not reproduce the molecular and electrophysiological environment of human cardiomyocytes, and the controlled WT/variant context used to model heterozygosity may not reproduce all aspects of endogenous allele-specific expression, subunit assembly or cellular regulation. MeTAL is an intermediate computational representation of ventricular repolarization and pseudo-ECG modelling rather than a complete model of ventricular repolarization, and therefore cannot capture all interactions among cardiac ionic currents. Clinical QTc is itself influenced by age, sex, heart rate, medications, autonomic state, and genetic and environmental modifiers, so a single ECG measurement may underestimate the functional contribution of a pathogenic variant. These limitations reinforce rather than diminish the importance of integrating functional and clinical evidence.

Together, our findings establish a scalable framework in which standardized multidimensional electrophysiology defines channel dysfunction and MeTAL translates this dysfunction into a clinically interpretable repolarization phenotype. Continuous calibration then quantifies its evidentiary strength, allowing it to contribute directly to variant reclassification within the ACMG/AMP framework, while patient phenotypes and penetrance provide complementary clinical context. Combined with open dissemination through HAMMER, this framework provides a direct path from high-throughput functional genomics to evidence-based interpretation of *KCNH2* variants in patients and their families.

## Data Availability

All data produced in the present study are available upon reasonable request to the authors

https://uncloud.univ-nantes.fr/index.php/s/TiXwNYwWXcPXmri

## Acknowledgements

The authors gratefully acknowledge the late Florence Kyndt (CHU Nantes), whose invaluable contributions to *KCNH2* variant database curation and ACMG classification were instrumental in the development of this study. Her dedication and expertise are deeply appreciated.

