## Supplementary data for "Nationwide Functional Profiling Enables Clinical Reclassification of *KCNH2* Variants in the French LQT2 Population"

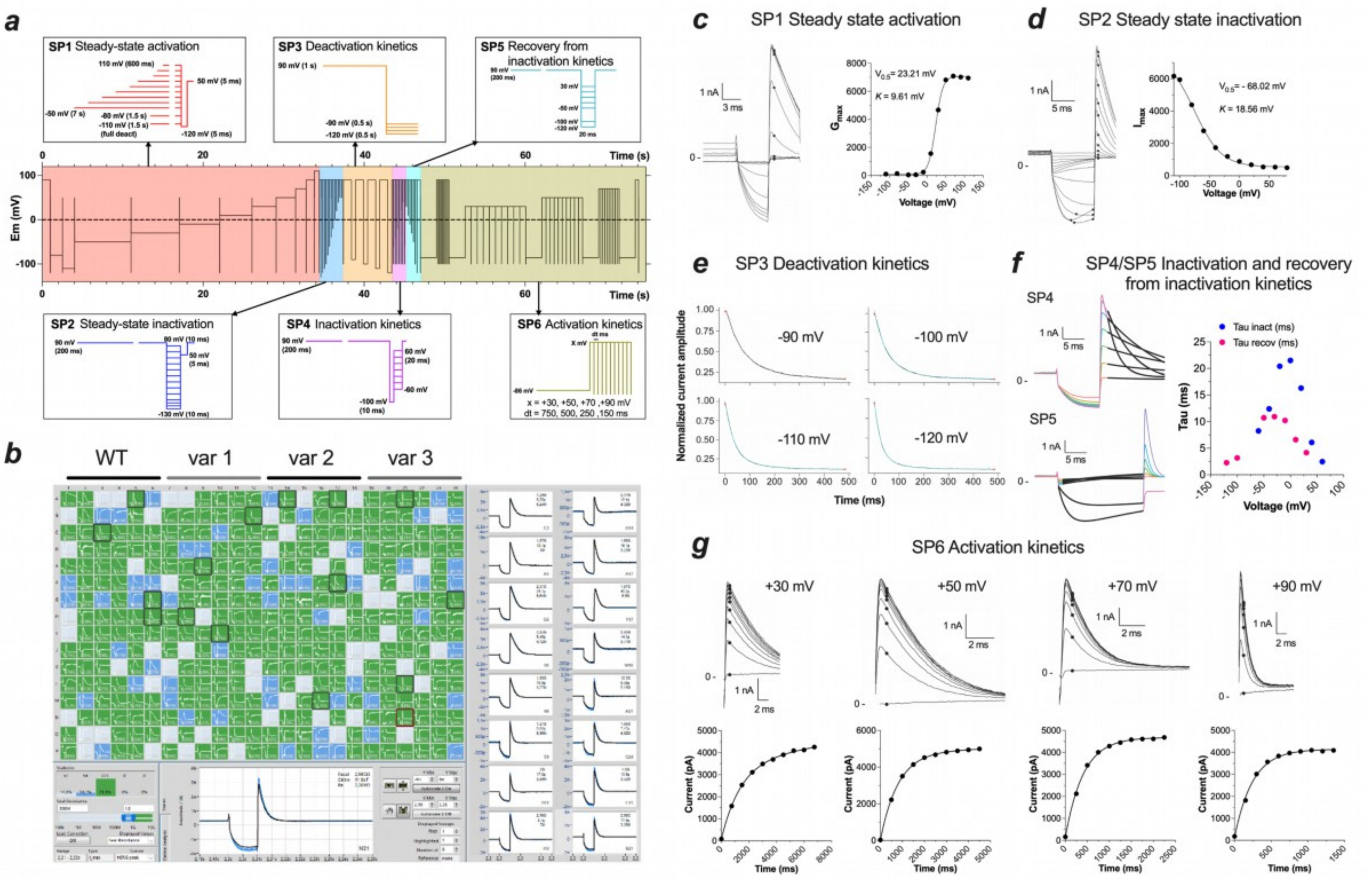
a
SP1 Steady-state activation
SP3 Deactivation kinetics
SP5 Recovery from inactivation kinetics
SP2 Steady-state inactivation
SP4 Inactivation kinetics
SP6 Activation kinetics
x = +30, +50, +70 ,+90 mV
dt = 750, 500, 250 ,150 ms
Time (s)
Em (mV)
b
WT
var 1
var 2
var 3
c
SP1 Steady state activation
V0.5= 23.21 mV
K = 9.61 mV
Voltage (mV)
d
SP2 Steady state inactivation
V0.5= - 68.02 mV
K = 18.56 mV
Voltage (mV)
e
SP3 Deactivation kinetics
Normalized current amplitude
-90 mV
-100 mV
-110 mV
-120 mV
Time (ms)
f
SP4/SP5 Inactivation and recovery from inactivation kinetics
SP4
SP5
Tau inact (ms)
Tau recov (ms)
Tau (ms)
Voltage (mV)
g
SP6 Activation kinetics
+30 mV
+50 mV
+70 mV
+90 mV
Current (pA)
Time (ms)

**Figure S1. Workflow for automated hERG characterization and data analysis pipeline.** ***a,*** Optimized fast-track voltage-clamp protocol for comprehensive extraction of hERG channel biophysical parameters [1]. The 74-s protocol is subdivided into six optimized subprotocols (SP). **SP1:** Steady-state activation protocol using depolarizing steps of variable duration, longer at lower voltages and shorter at higher voltages. **SP2:** Steady-state inactivation protocol, consisting of a single activating pulse to fully open activation gates prior to assessment of inactivation. **SP3:** Deactivation kinetics, with time constants extracted at −120, −110, −100, and −90 mV. **SP4:** Inactivation kinetics measured following channel activation at +90 mV. **SP5:** Recovery from inactivation measured following channel activation/inactivation at +90 mV. **SP6:** Activation kinetics measured at +30, +50, +70, and +90 mV using repeated depolarizing pulses separated by 750, 500, 250, and 150 ms hyperpolarizing steps to −100 mV to fully remove inactivation and quantify cumulative channel activation. Peak currents were measured for each activation step. ***b,*** Representative automated patch-clamp recordings acquired on the SyncroPatch platform, illustrating one plate containing hERG WT and three variant constructs. ***c–g,*** Output files generated by the automated R analysis pipeline for quantitative analysis of voltage-clamp data from the six optimized subprotocols (SP1-SP6).

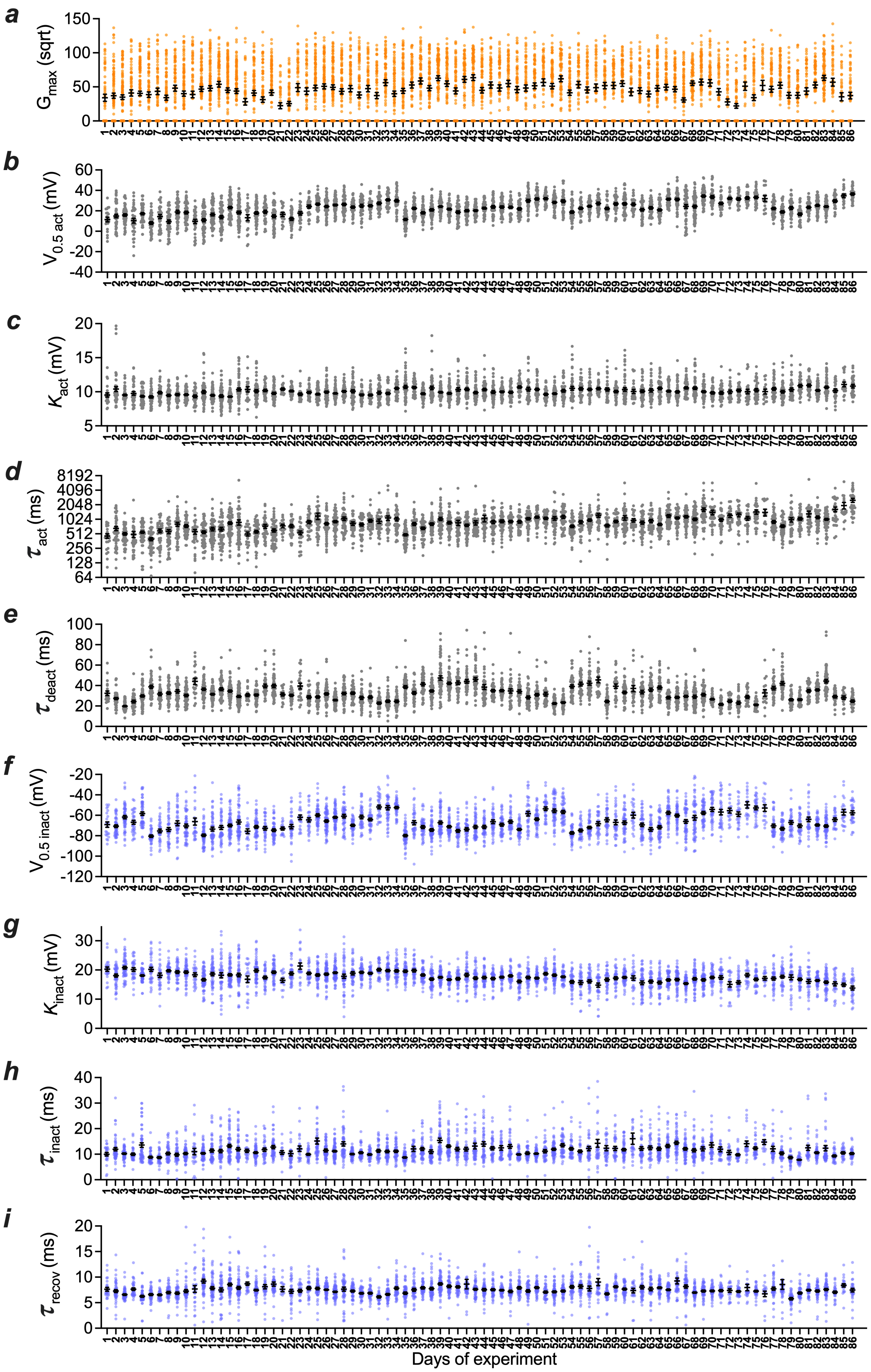


**Figure S2. Stability of nine hERG WT parameters across experimental days.** ***a-i*****,** WT hERG recordings were acquired daily throughout the experimental campaign to monitor assay stability and to enable direct comparison between WT and variant measurements obtained on the same day.

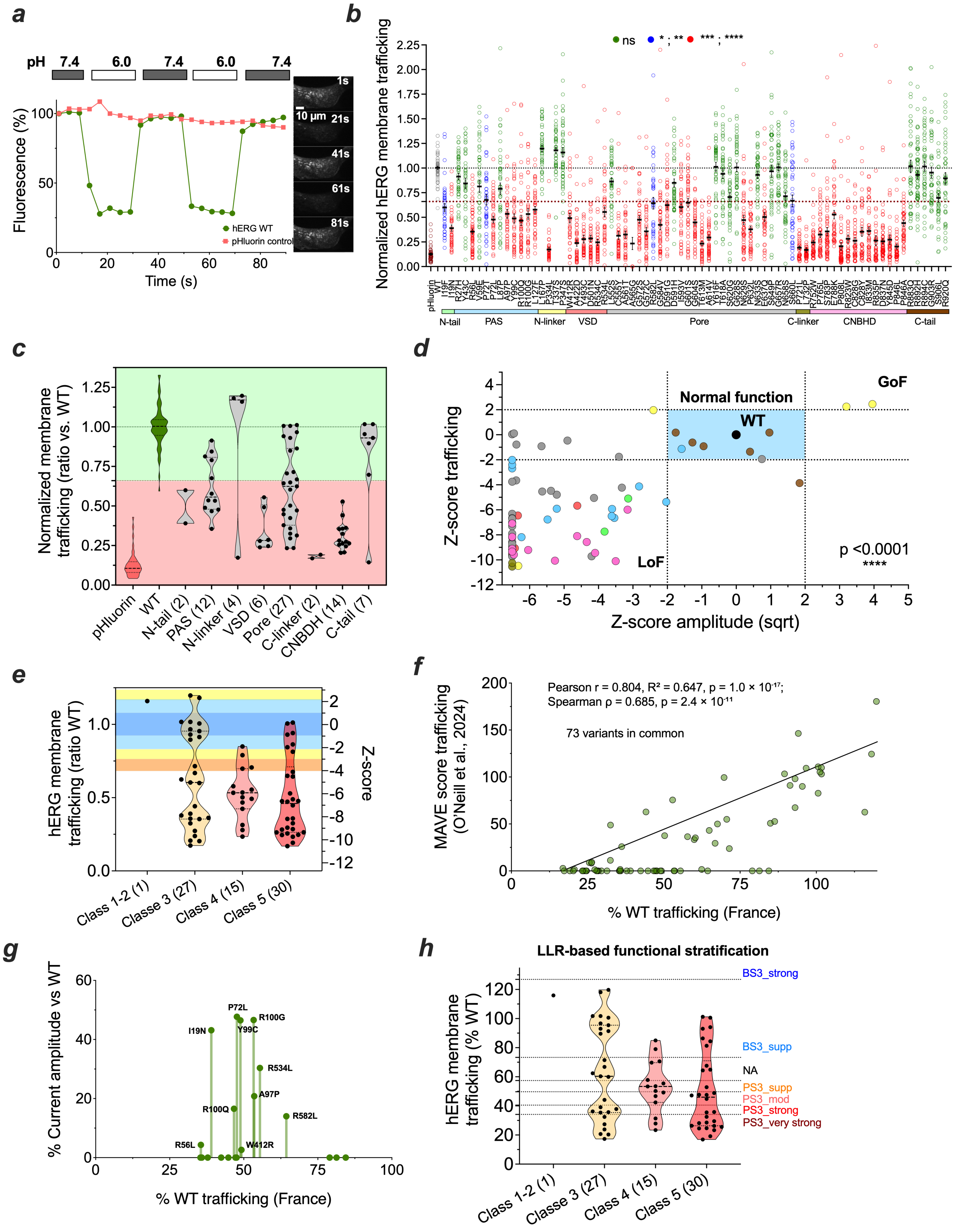

**Figure S3. Trafficking analysis of 73 French hERG variants using pHluorin-tagged chimeric constructs. *a,*** Representative confocal imaging recording of pHluorin fluorescence in MDCK cells transfected with WT-hERG-pHluorin plasmid and pHluorin control plasmid, showing fluorescence quenching at pH 6.0 relative to pH 7.4. Plasma membrane fluorescence was calculated as the difference between total fluorescence measured at pH 7.4 and intracellular fluorescence remaining after extracellular pHluorin quenching at pH 6.0. ***b,*** Quantification of plasma membrane expression and total expression of 73 French hERG variants using chimeric constructs carrying a pHluorin tag, allowing discrimination between surface-expressed channels and total channel expression (mean ± SEM of the percentage fluorescence variation [(pH 7.4 − pH 6.0) / pH 7.4]) [2]. Forty-eight variants exhibited loss-of-function trafficking defects (threshold ratio = 0.66). Basal pHluorin fluorescence was also recorded. Sample sizes ranged from n = 38 to 65 cells per condition (WT and variants). Statistical significance versus WT hERG: ****p < 0.0001, ***p < 0.001, **p < 0.01, *p < 0.05 (Kruskal-Wallis test). **c,** Domain-based analysis of trafficking defects across variants. Gain-of-function variants located in the N-linker domain showed a trend toward mildly increased surface trafficking. In contrast, loss-of-function variants located in the pore or PAS domains frequently exhibited preserved or reduced trafficking, indicating multiple mechanisms of loss of function, including gating-related and trafficking-related defects. Variants located in the voltage-sensing domain (VSD), C-linker, and cyclic nucleotide-binding homology domain (CNBHD) displayed the most pronounced trafficking impairments. ***d,*** Relationship between membrane trafficking and current amplitude for 73 hERG variants, expressed as Z-scores relative to WT. Current amplitude values were square-root transformed before Z-score calculation. Data points are color-coded according to the structural domain of each variant, as in panel **b**. Among the 63 variants exhibiting loss-of-function (LoF) in current amplitude (Z-score < −2), 55 (87.3%) also showed impaired membrane trafficking (Z-score < −2), whereas eight LoF variants displayed normal trafficking, suggesting functional impairment through mechanisms other than defective membrane trafficking. Among the eight variants with normal current amplitude, seven displayed trafficking levels within the normal range. Two gain-of-function (GoF) variants showed both increased current amplitude and increased membrane trafficking. The association between current amplitude and membrane trafficking was highly significant ($p < 0.0001$). ***e,*** Distribution of hERG membrane trafficking across ACMG classes, expressed as WT-normalized membrane trafficking ratio (left axis) and corresponding Z-score (right axis). Class 4 and 5 variants showed a high proportion of trafficking-deficient variants, whereas the only benign variant analyzed (P347S, class 1) displayed normal membrane trafficking. ***f,*** Correlation of membrane trafficking measurements obtained in the present study with MAVE trafficking scores reported by O'Neill et al. for the 73 variants common to both datasets. A significant correlation was observed between trafficking measurements from the two studies (Pearson $r = 0.804$, $R^2 = 0.647$, $p = 1.0 \times 10^{-17}$; Spearman $\rho = 0.685$, $p = 2.4 \times 10^{-11}$). ***g,*** Functional characterization of variants displaying discrepant trafficking measurements between the O'Neill et al. MAVE dataset [3] and the present study. Eight variants (I19N, P72L, A97P, Y99C, R100G, R100Q, R534L, and R582L) reported as having near-zero membrane trafficking in the MAVE dataset showed substantial residual trafficking in the present study, ranging from 39% to 64% of WT. Consistent with the presence of trafficking-preserved channels, electrophysiological characterization also revealed measurable residual hERG current for all eight variants, with current amplitudes ranging from 14% to 47% of WT. Each vertical line connects the trafficking and current-amplitude measurements obtained for the same variant. ***h,*** Functional stratification of hERG variants using LLR-calibrated trafficking thresholds. Membrane trafficking of hERG variants, expressed as a % relative to WT, is shown according to ACMG classification. Each dot represents an individual variant. Previously established log-likelihood ratio (LLR)-calibrated trafficking thresholds from O'Neill et al. were applied to the pHluorin trafficking measurements to define different strengths of functional evidence. Trafficking levels below 57.4%, 49.7%, 40.5%, and 34.12% of WT correspond to PS3_Supporting, PS3_Moderate, PS3_Strong, and PS3_Very Strong evidence, respectively, whereas trafficking levels above 73.3% and 126.9% of WT correspond to BS3_Supporting and BS3_Strong evidence, respectively. These thresholds were externally derived and were not recalibrated using the present dataset.

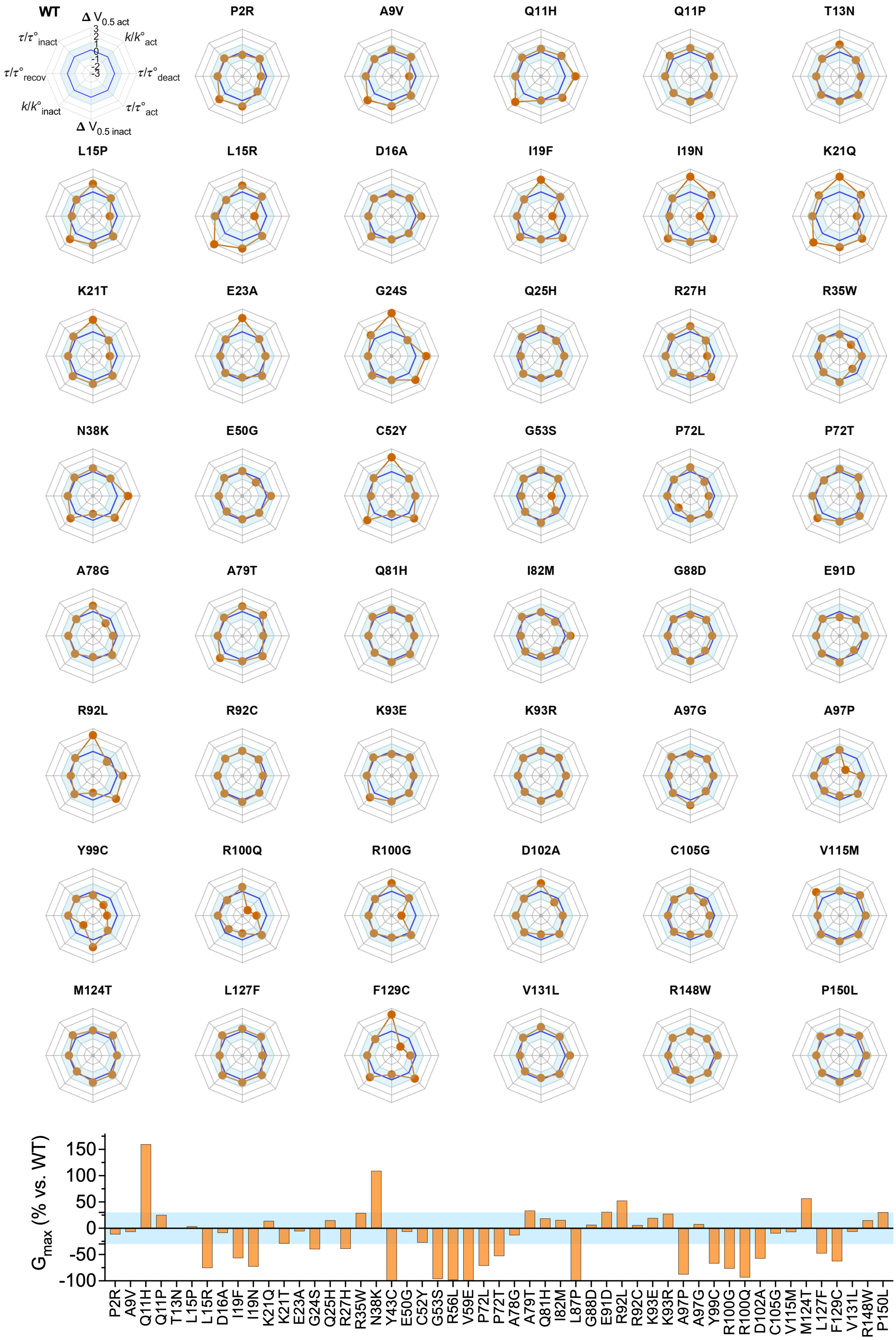
WT
Δ $V_{0.5\ act}$
$k/k°_{act}$
$\tau/\tau°_{deact}$
$\tau/\tau°_{act}$
Δ $V_{0.5\ inact}$
$k/k°_{inact}$
$\tau/\tau°_{recov}$
$\tau/\tau°_{inact}$
3
2
1
0
-1
-2
-3
P2R
A9V
Q11H
Q11P
T13N
L15P
L15R
D16A
I19F
I19N
K21Q
K21T
E23A
G24S
Q25H
R27H
R35W
N38K
E50G
C52Y
G53S
P72L
P72T
A78G
A79T
Q81H
I82M
G88D
E91D
R92L
R92C
K93E
K93R
A97G
A97P
Y99C
R100Q
R100G
D102A
C105G
V115M
M124T
L127F
F129C
V131L
R148W
P150L
$G_{max}$ (% vs. WT)
150
100
50
0
-50
-100
P2R
A9V
Q11H
Q11P
T13N
L15P
L15R
D16A
I19F
I19N
K21Q
K21T
E23A
G24S
Q25H
R27H
R35W
N38K
Y43C
E50G
C52Y
G53S
R56L
V59E
P72L
P72T
A78G
A79T
Q81H
I82M
L87P
G88D
E91D
R92L
R92C
K93E
K93R
A97P
A97G
Y99C
R100G
R100Q
D102A
C105G
V115M
M124T
L127F
F129C
V131L
R148W
P150L

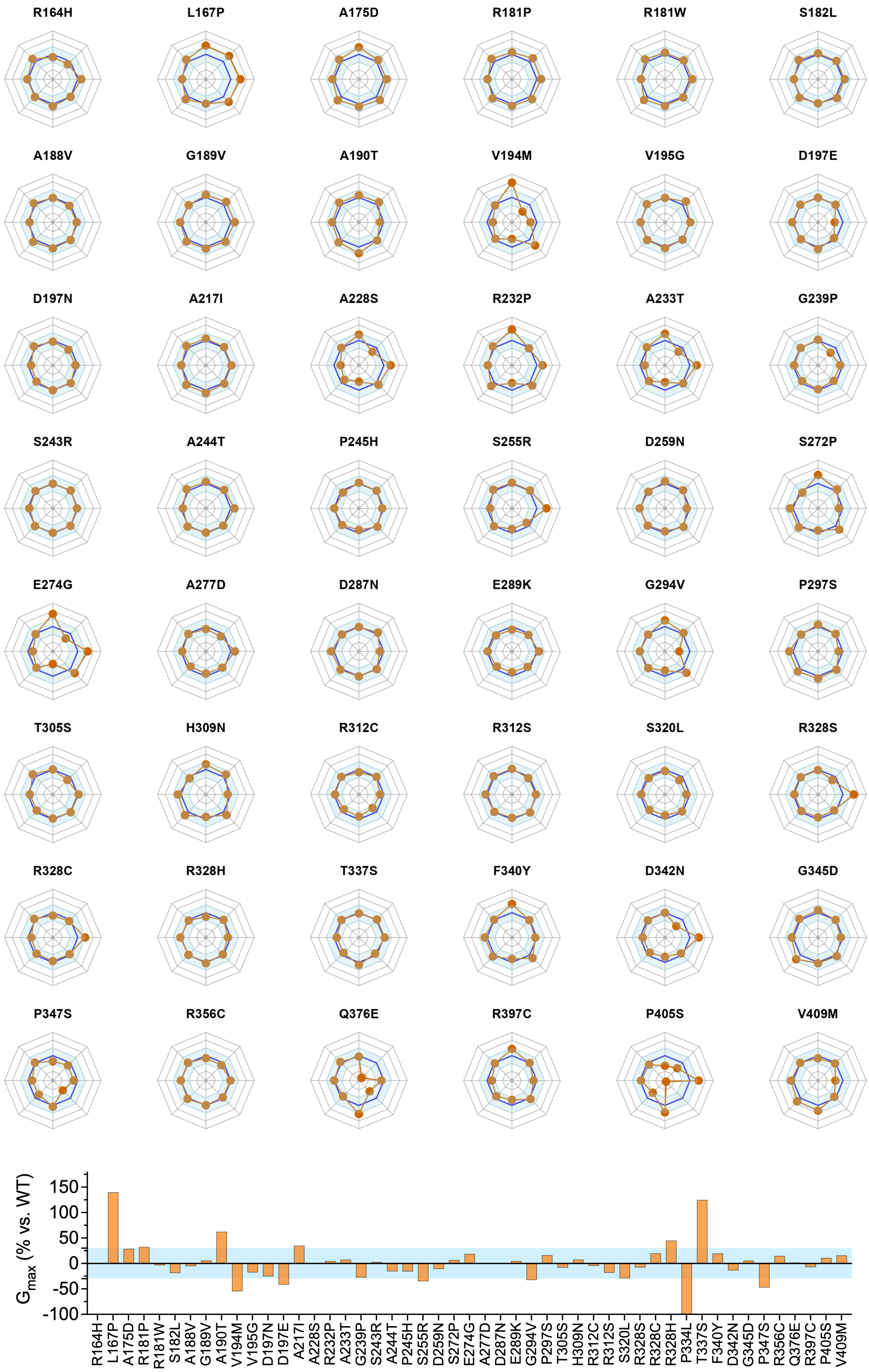
R164H
L167P
A175D
R181P
R181W
S182L
A188V
G189V
A190T
V194M
V195G
D197E
D197N
A217I
A228S
R232P
A233T
G239P
S243R
A244T
P245H
S255R
D259N
S272P
E274G
A277D
D287N
E289K
G294V
P297S
T305S
H309N
R312C
R312S
S320L
R328S
R328C
R328H
T337S
F340Y
D342N
G345D
P347S
R356C
Q376E
R397C
P405S
V409M
$G_{max}$ (% vs. WT)
150
100
50
0
-50
-100
R164H
L167P
A175D
R181P
R181W
S182L
A188V
G189V
A190T
V194M
V195G
D197N
D197E
A217I
A228S
R232P
A233T
G239P
S243R
A244T
P245H
S255R
D259N
S272P
E274G
A277D
D287N
E289K
G294V
P297S
T305S
H309N
R312C
R312S
S320L
R328S
R328C
R328H
P334L
T337S
F340Y
D342N
G345D
P347S
R356C
Q376E
R397C
P405S
V409M

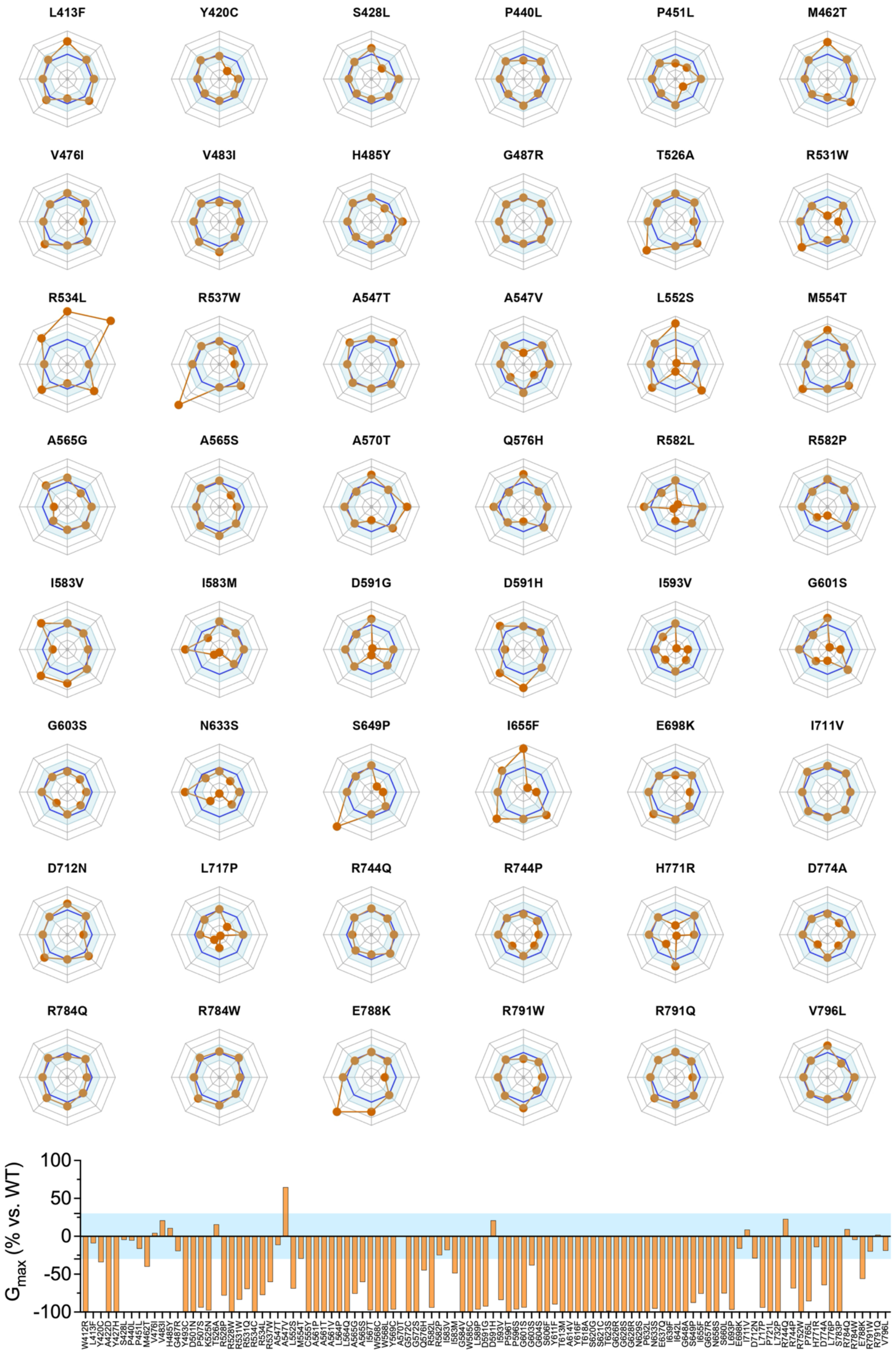

L413F
Y420C
S428L
P440L
P451L
M462T
V476I
V483I
H485Y
G487R
T526A
R531W
R534L
R537W
A547T
A547V
L552S
M554T
A565G
A565S
A570T
Q576H
R582L
R582P
I583V
I583M
D591G
D591H
I593V
G601S
G603S
N633S
S649P
I655F
E698K
I711V
D712N
L717P
R744Q
R744P
H771R
D774A
R784Q
R784W
E788K
R791W
R791Q
V796L
100
50
0
-50
-100
$G_{max}$ (% vs. WT)
W412R
L413F
Y420C
A422D
Y427H
S428L
P440L
P451L
M462T
V476I
V483I
H485Y
G487R
Y493C
D501N
P507S
K525N
T526A
R528P
R528W
R531W
R531Q
R534C
R534L
R537W
A547T
A547V
L552S
M554T
C555Y
A561P
A561T
A561V
L564P
L564Q
A565G
A565S
I567T
W568C
W568L
Y569C
A570T
G572C
G572S
Q576H
R582L
R582P
I583V
I583M
G584V
W585C
L589P
D591G
D591H
I593V
P596T
P596S
G601S
G603S
G604S
S606F
Y611F
T613M
A614V
Y616F
T618A
S620G
S621C
T623S
G626R
G628S
G628R
N629S
P632L
N633S
E637Q
I639F
I642L
G648A
S649P
I655F
G657R
N658S
S660L
L693P
E698K
I711V
D712N
L717P
P721L
L732P
R744Q
R744P
R752W
P765L
H771R
D774A
L776P
S783P
R784Q
R784W
E788K
R791W
R791Q
V796L

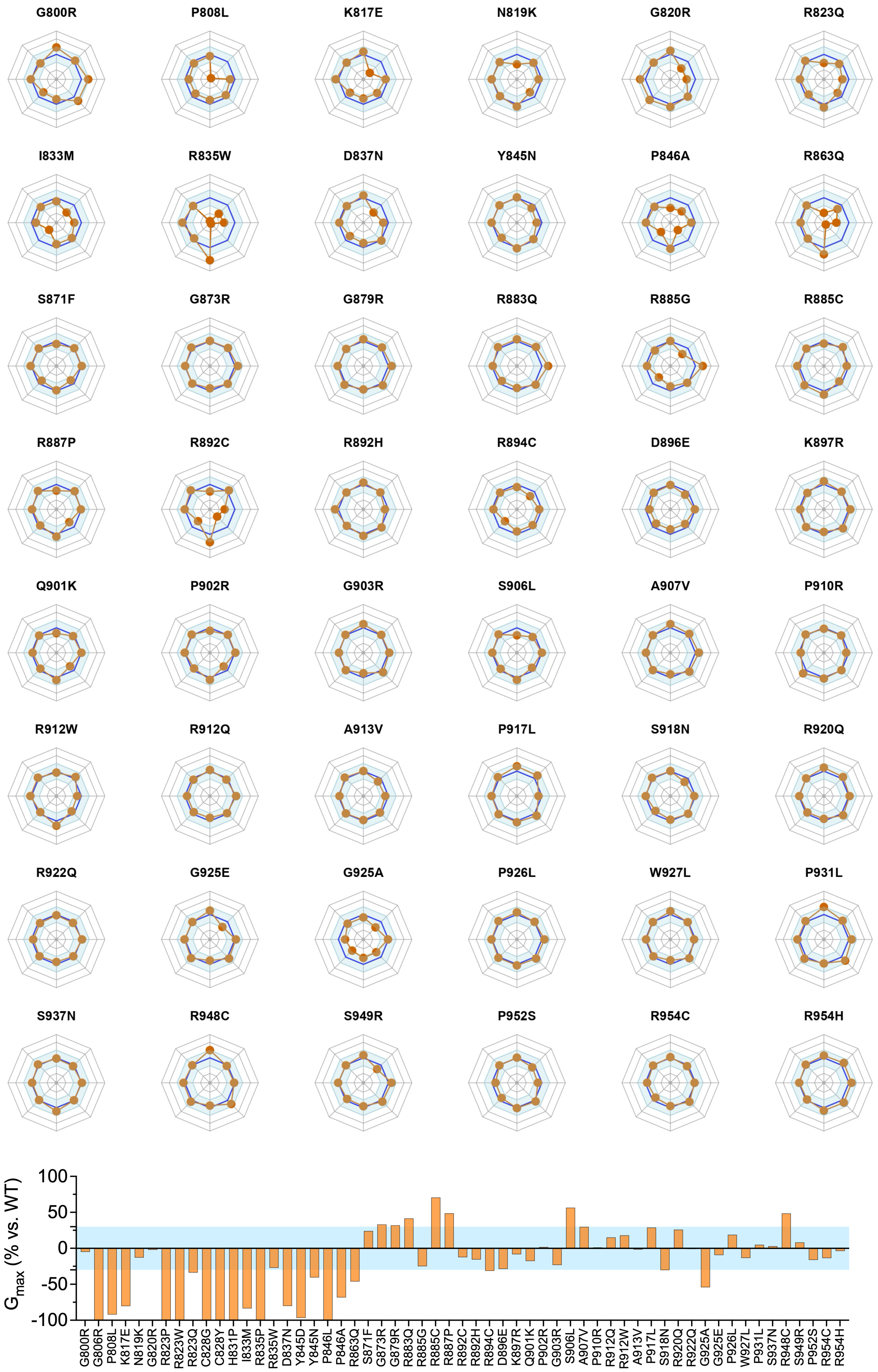
G800R
P808L
K817E
N819K
G820R
R823Q
I833M
R835W
D837N
Y845N
P846A
R863Q
S871F
G873R
G879R
R883Q
R885G
R885C
R887P
R892C
R892H
R894C
D896E
K897R
Q901K
P902R
G903R
S906L
A907V
P910R
R912W
R912Q
A913V
P917L
S918N
R920Q
R922Q
G925E
G925A
P926L
W927L
P931L
S937N
R948C
S949R
P952S
R954C
R954H
100
50
0
-50
-100
$G_{max}$ (% vs. WT)
G800R
G806R
P808L
K817E
N819K
G820R
R823P
R823W
R823Q
C828G
C828Y
H831P
I833M
R835P
R835W
D837N
Y845D
Y845N
P846L
P846A
R863Q
S871F
G873R
G879R
R883Q
R885G
R885C
R887P
R892C
R892H
R894C
D896E
K897R
Q901K
P902R
G903R
S906L
A907V
P910R
R912Q
R912W
A913V
P917L
S918N
R920Q
R922Q
G925A
G925E
P926L
W927L
P931L
S937N
R948C
S949R
P952S
R954C
R954H

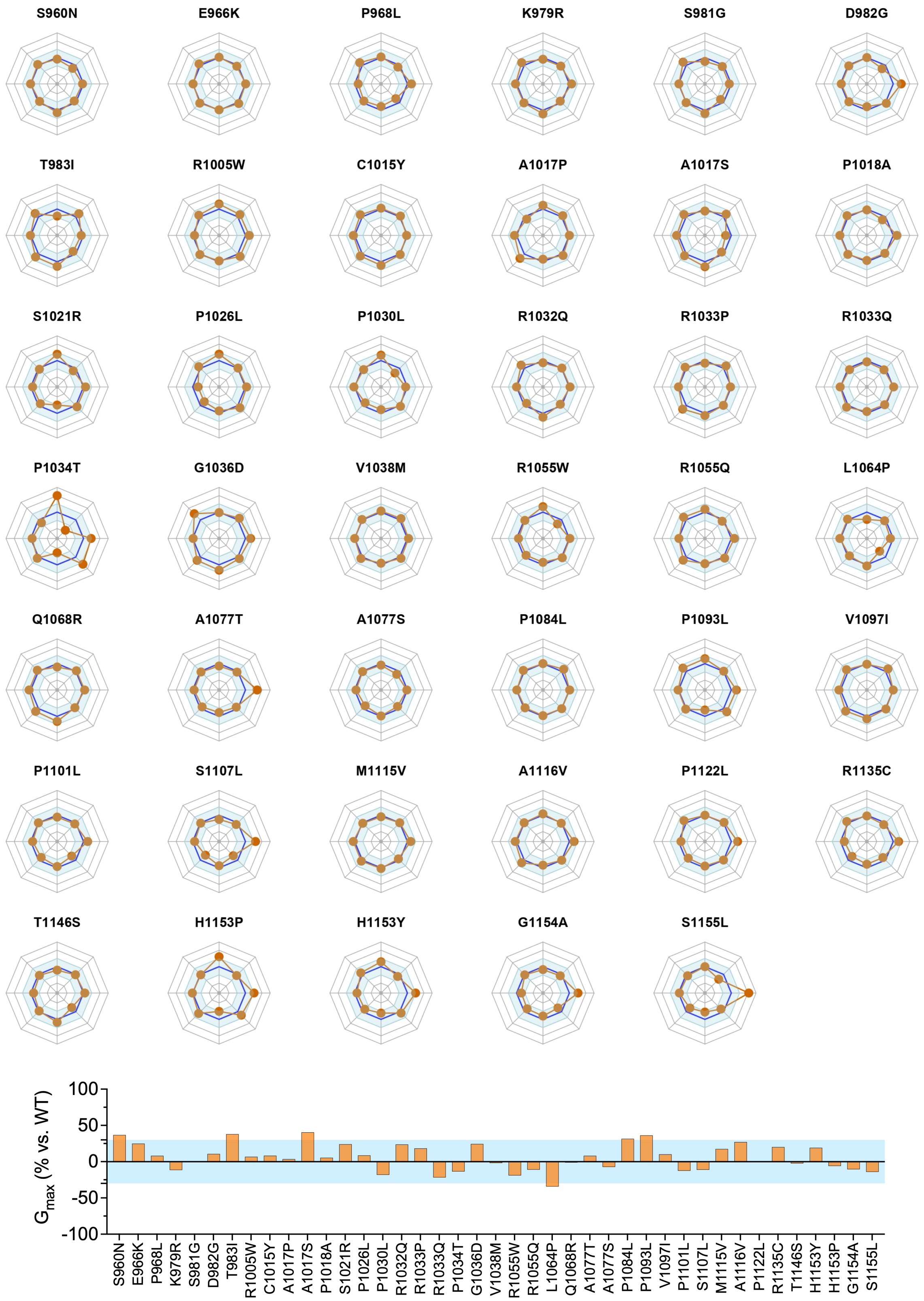


**Figure S4. Radar plots of 232 hERG variants under homozygous conditions.** Radar plots showing the variant-specific biophysical fingerprints of 232 hERG variants under homozygous conditions, including only variants with sufficient current for analysis (n ≥ 5 for each parameter). Each axis represents a normalized gating parameter, and concentric lines indicate Z-score deviations relative to wild type.

The blue shaded area denotes the wild-type range (±1 Z-score), enabling direct visualization of the magnitude and direction of variant-induced changes (outward: GoF or inward: LoF) across parameters. Orange dots indicate the Z-score of each variant parameter relative to wild-type. The current amplitude variation relative to wild type (Gmax, % vs. WT) is shown below radar plots, expressed as the percentage change in current amplitude for each variant. Together, these plots illustrate the broad diversity of biophysical phenotypes among variants, including isolated defects, combined alterations across multiple parameters, and mixed GoF/LoF profiles.

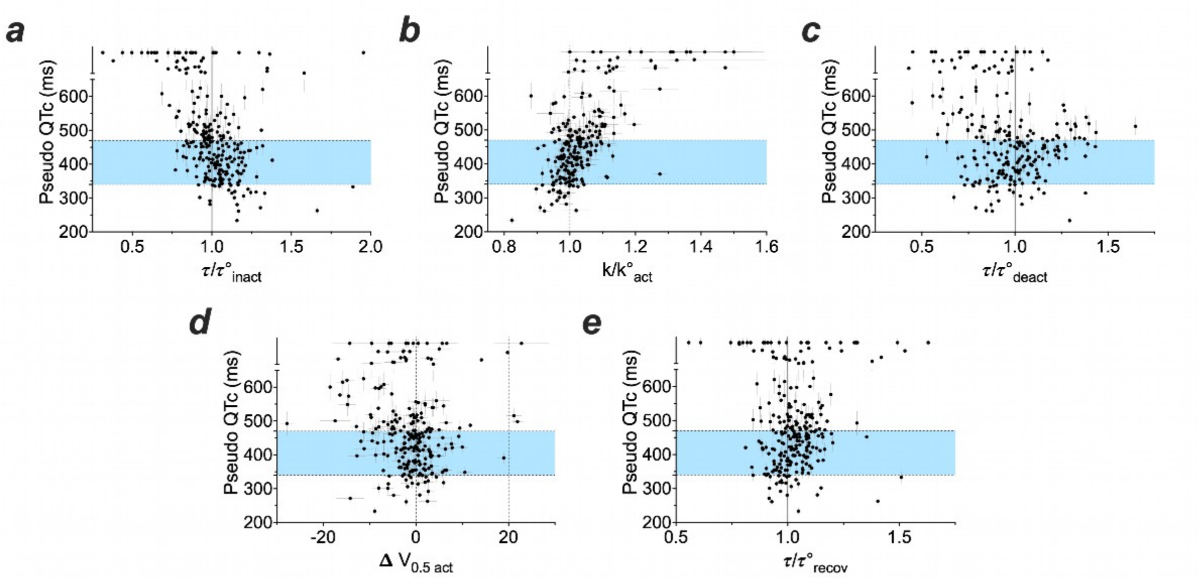


**Figure S5. Relationship between predicted QTc and individual hERG biophysical parameters in homozygous conditions**. ***a-e,*** Predicted QTc values under homozygous conditions were plotted as a function of individual biophysical parameters $\tau_{inact}$, $k_{act}$, $\tau_{deact}$, $V_{0.5\ act}$, $\tau_{recov}$. Although none of these parameters alone was identified as a major determinant by MeTAL, their combined alterations may have a greater impact than expected.

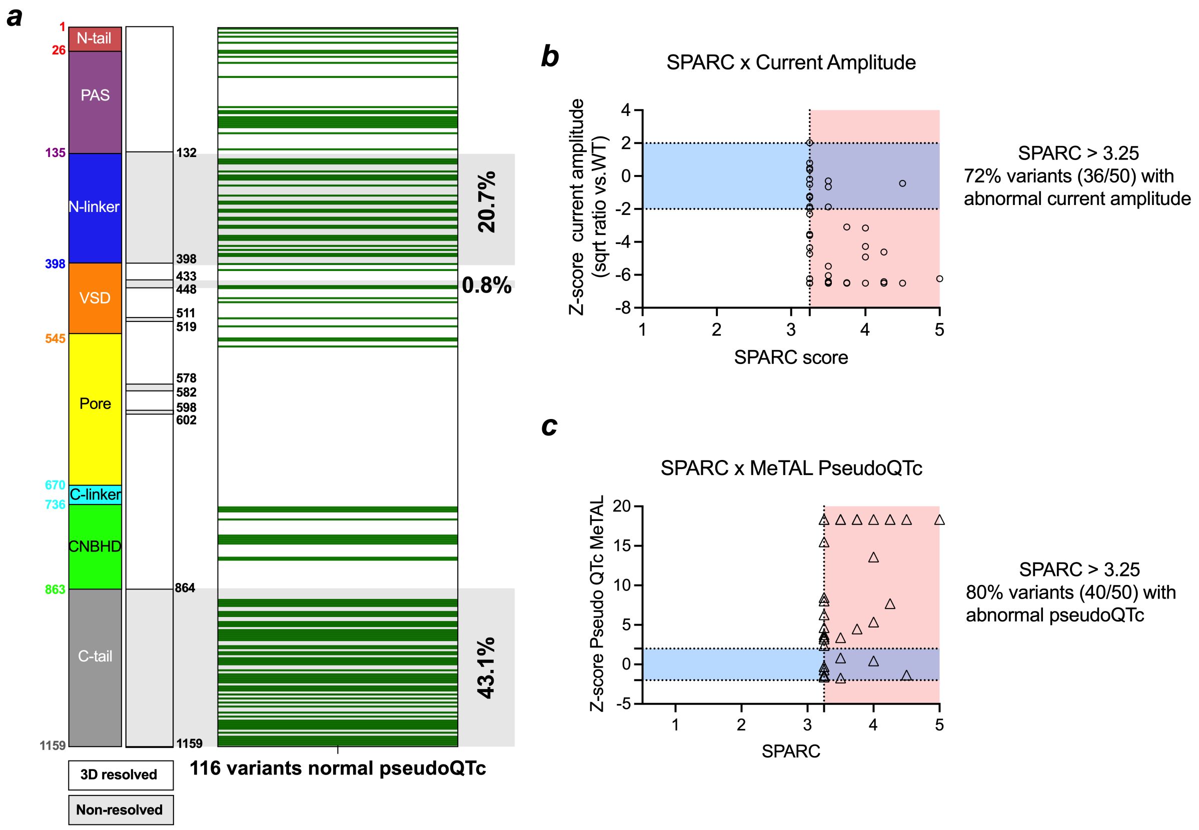


**Figure S6. Relationship between SPARC-predicted structural impact and functional phenotypes of hERG variants.** ***a,*** All 116 hERG variants displaying normal pseudoQTc values were mapped onto the hERG protein structure irrespective of their ACMG classification. A majority of benign variants (75/116, 64.6%) localized to intrinsically disordered regions (gray), consistent with these regions being more permissive to amino acid substitutions, as previously predicted by the SPARC binary classifier. Of these, 24 variants were found in the N-linker, 50 in the C-tail, and only one in the VSD. Variants located within structured domains (PAS, pore, and CNBHD) were less frequent. ***b,*** Relationship between SPARC score and current-amplitude Z-score. A SPARC score >3.25 was considered predictive of a structurally impactful variant. Among variants exceeding this threshold, 72% (36/50) displayed an abnormal current-amplitude phenotype, indicating substantial concordance between predicted structural impact and experimentally measured hERG dysfunction. The blue-shaded region indicates the normal range for current-amplitude Z-scores, whereas red-shaded regions indicate abnormal values. ***c,*** Relationship between SPARC score and MeTAL pseudoQTc. Among variants with SPARC scores >3.25, 80% (44/55) displayed an abnormal modeled pseudoQTc phenotype, further supporting the association between SPARC-predicted structural impact and functional impairment. The blue-shaded region indicates the normal pseudoQTc range, whereas red-shaded regions indicate abnormal values.

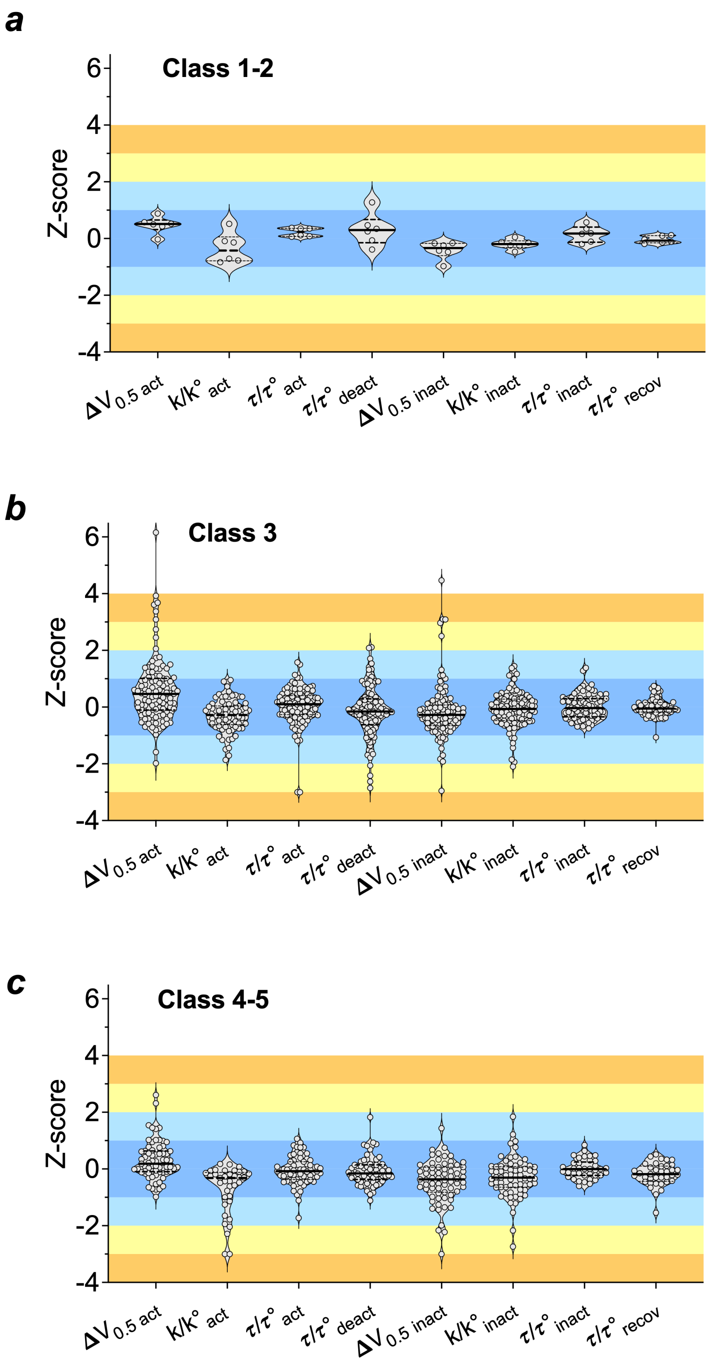


**Figure S7. Distribution of biophysical parameter alterations across ACMG classes in heterozygous conditions.** ***a-c*,** Violin plots showing the distribution of normalized biophysical parameters measured in heterozygous conditions for *KCNH2* variants classified as ACMG classes 1–2, class 3, and classes 4–5. Functional effects are represented as Z-scores calculated relative to the wild-type (WT) control recorded on the same experimental day. Parameters include voltage dependence of activation ($\Delta V_{0.5\ act}$), activation slope factor ($k/k°_{act}$), activation time constant ($\tau/\tau°_{act}$), deactivation time constant ($\tau/\tau°_{deact}$), voltage dependence of inactivation ($\Delta V_{0.5\ inact}$), inactivation slope factor ($k/k°_{inact}$), inactivation time constant ($\tau/\tau°_{inact}$), and recovery from inactivation time constant ($\tau/\tau°_{recov}$). The shaded blue area represents the normal functional range ($-2 \leq Z \leq 2$), while yellow and orange regions indicate moderate and severe deviations from WT function, respectively. Variants classified as

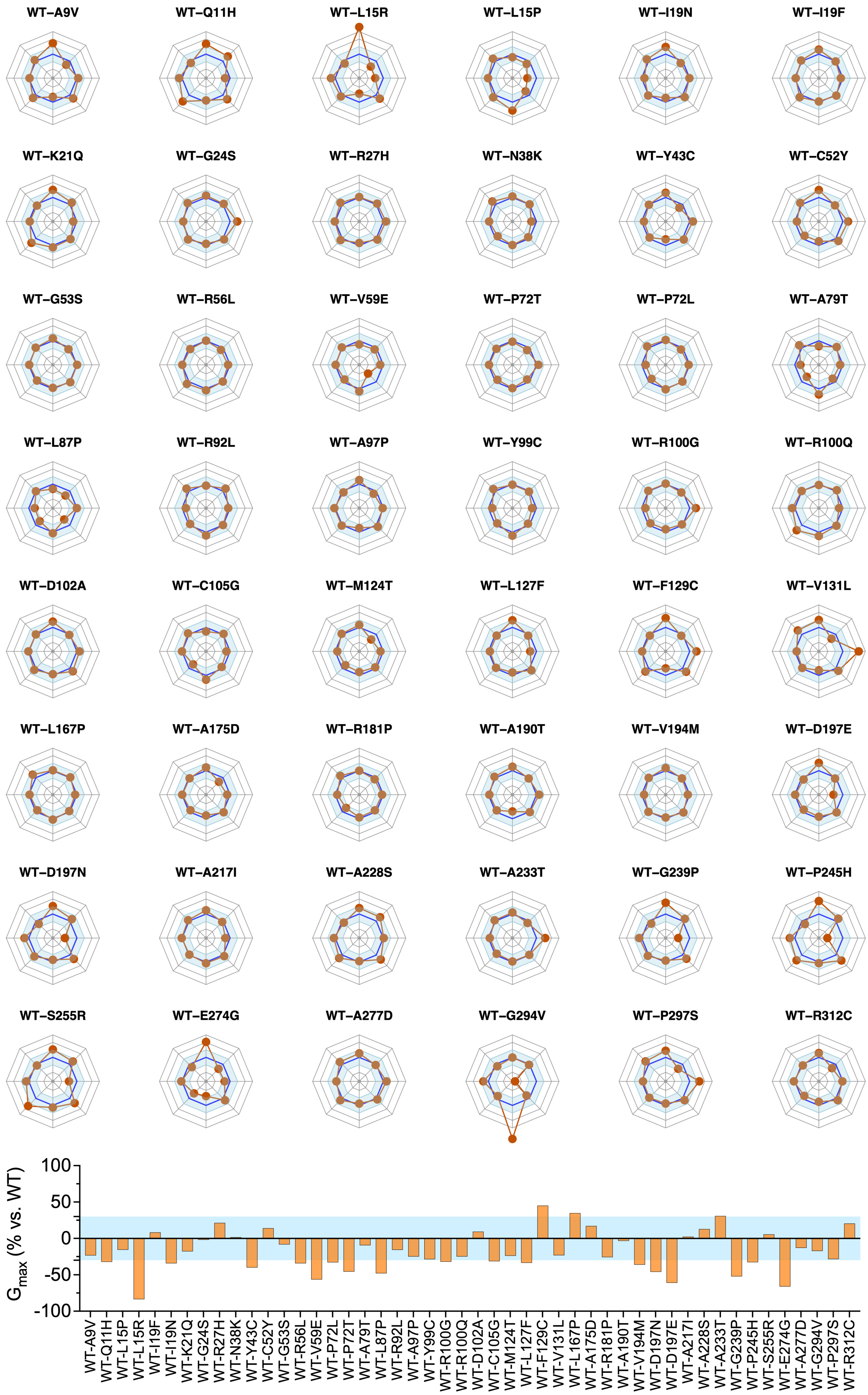

WT–A9V
WT–Q11H
WT–L15R
WT–L15P
WT–I19N
WT–I19F
WT–K21Q
WT–G24S
WT–R27H
WT–N38K
WT–Y43C
WT–C52Y
WT–G53S
WT–R56L
WT–V59E
WT–P72T
WT–P72L
WT–A79T
WT–L87P
WT–R92L
WT–A97P
WT–Y99C
WT–R100G
WT–R100Q
WT–D102A
WT–C105G
WT–M124T
WT–L127F
WT–F129C
WT–V131L
WT–L167P
WT–A175D
WT–R181P
WT–A190T
WT–V194M
WT–D197E
WT–D197N
WT–A217I
WT–A228S
WT–A233T
WT–G239P
WT–P245H
WT–S255R
WT–E274G
WT–A277D
WT–G294V
WT–P297S
WT–R312C
$G_{max}$ (% vs. WT)
100
50
0
-50
-100

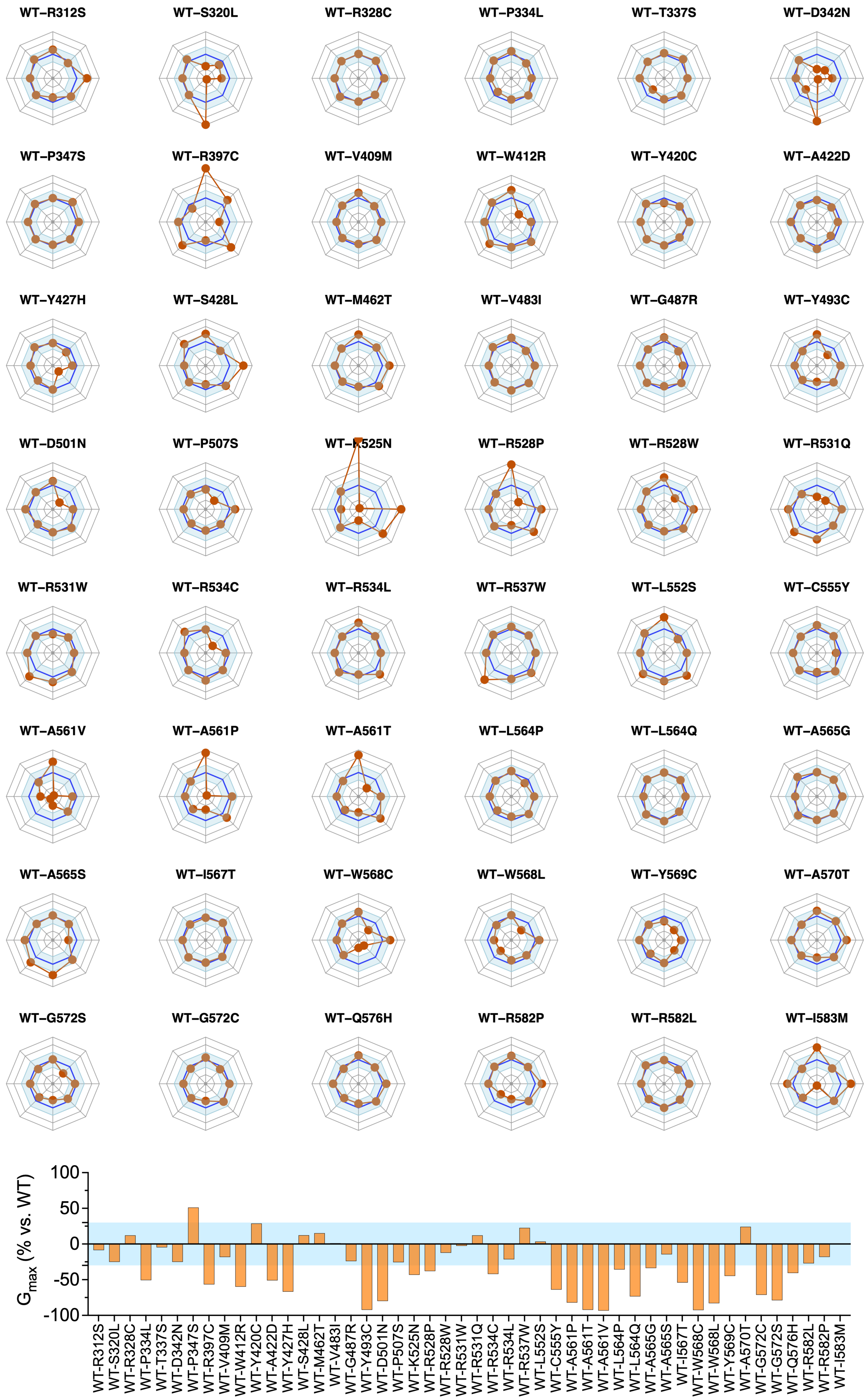
WT–R312S
WT–S320L
WT–R328C
WT–P334L
WT–T337S
WT–D342N
WT–P347S
WT–R397C
WT–V409M
WT–W412R
WT–Y420C
WT–A422D
WT–Y427H
WT–S428L
WT–M462T
WT–V483I
WT–G487R
WT–Y493C
WT–D501N
WT–P507S
WT–K525N
WT–R528P
WT–R528W
WT–R531Q
WT–R531W
WT–R534C
WT–R534L
WT–R537W
WT–L552S
WT–C555Y
WT–A561V
WT–A561P
WT–A561T
WT–L564P
WT–L564Q
WT–A565G
WT–A565S
WT–I567T
WT–W568C
WT–W568L
WT–Y569C
WT–A570T
WT–G572S
WT–G572C
WT–Q576H
WT–R582P
WT–R582L
WT–I583M
$G_{max}$ (% vs. WT)
100
50
0
-50
-100

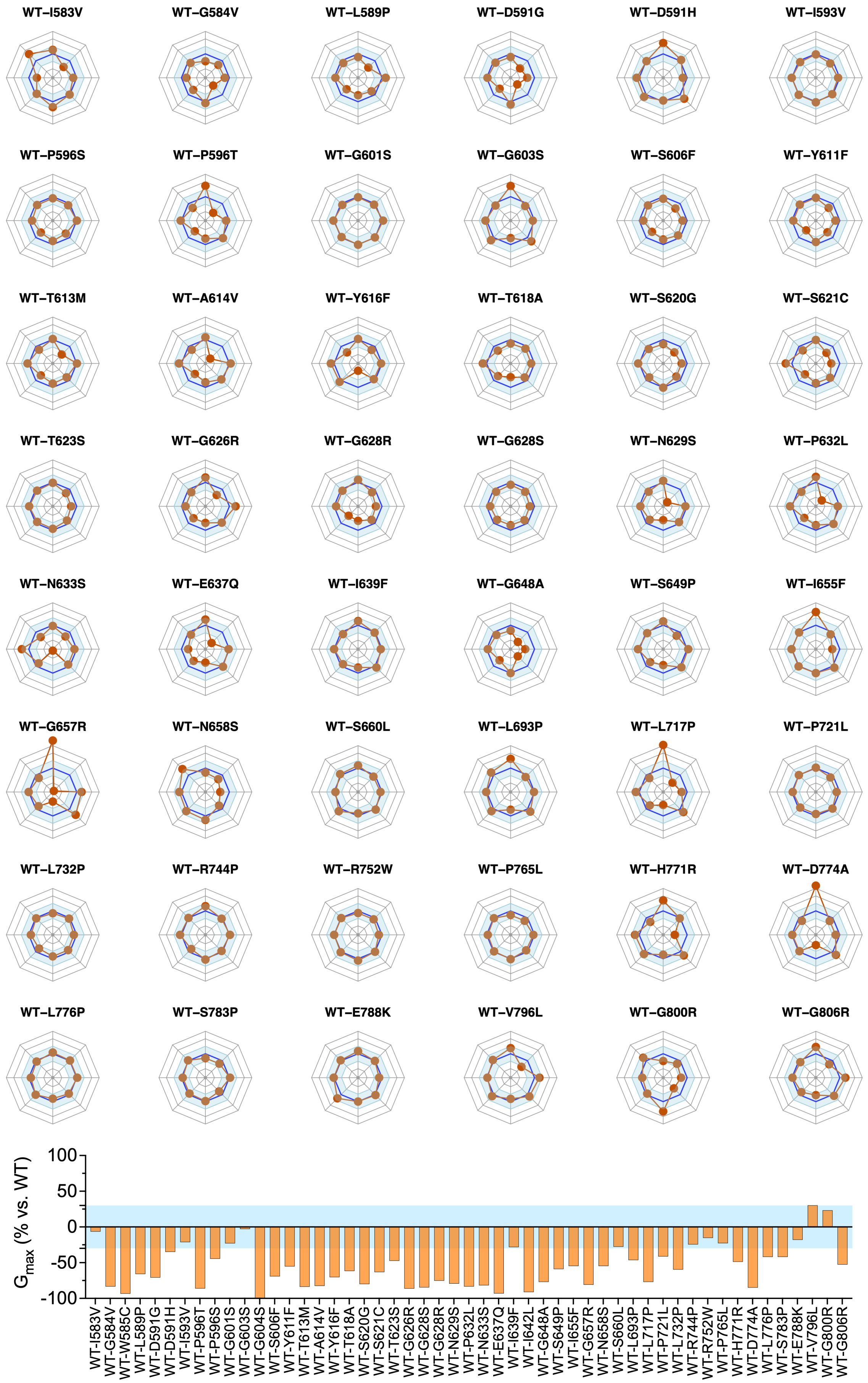

WT–I583V
WT–G584V
WT–L589P
WT–D591G
WT–D591H
WT–I593V
WT–P596S
WT–P596T
WT–G601S
WT–G603S
WT–S606F
WT–Y611F
WT–T613M
WT–A614V
WT–Y616F
WT–T618A
WT–S620G
WT–S621C
WT–T623S
WT–G626R
WT–G628R
WT–G628S
WT–N629S
WT–P632L
WT–N633S
WT–E637Q
WT–I639F
WT–G648A
WT–S649P
WT–I655F
WT–G657R
WT–N658S
WT–S660L
WT–L693P
WT–L717P
WT–P721L
WT–L732P
WT–R744P
WT–R752W
WT–P765L
WT–H771R
WT–D774A
WT–L776P
WT–S783P
WT–E788K
WT–V796L
WT–G800R
WT–G806R
G max (% vs. WT)
100
50
0
-50
-100
WT-I583V
WT-G584V
WT-W585C
WT-L589P
WT-D591G
WT-D591H
WT-I593V
WT-P596T
WT-P596S
WT-G601S
WT-G603S
WT-G604S
WT-S606F
WT-Y611F
WT-T613M
WT-A614V
WT-Y616F
WT-T618A
WT-S620G
WT-S621C
WT-T623S
WT-G626R
WT-G628S
WT-G628R
WT-N629S
WT-P632L
WT-N633S
WT-E637Q
WT-I639F
WT-I642L
WT-G648A
WT-S649P
WT-I655F
WT-G657R
WT-N658S
WT-S660L
WT-L693P
WT-L717P
WT-P721L
WT-L732P
WT-R744P
WT-R752W
WT-P765L
WT-H771R
WT-D774A
WT-L776P
WT-S783P
WT-E788K
WT-V796L
WT-G800R
WT-G806R

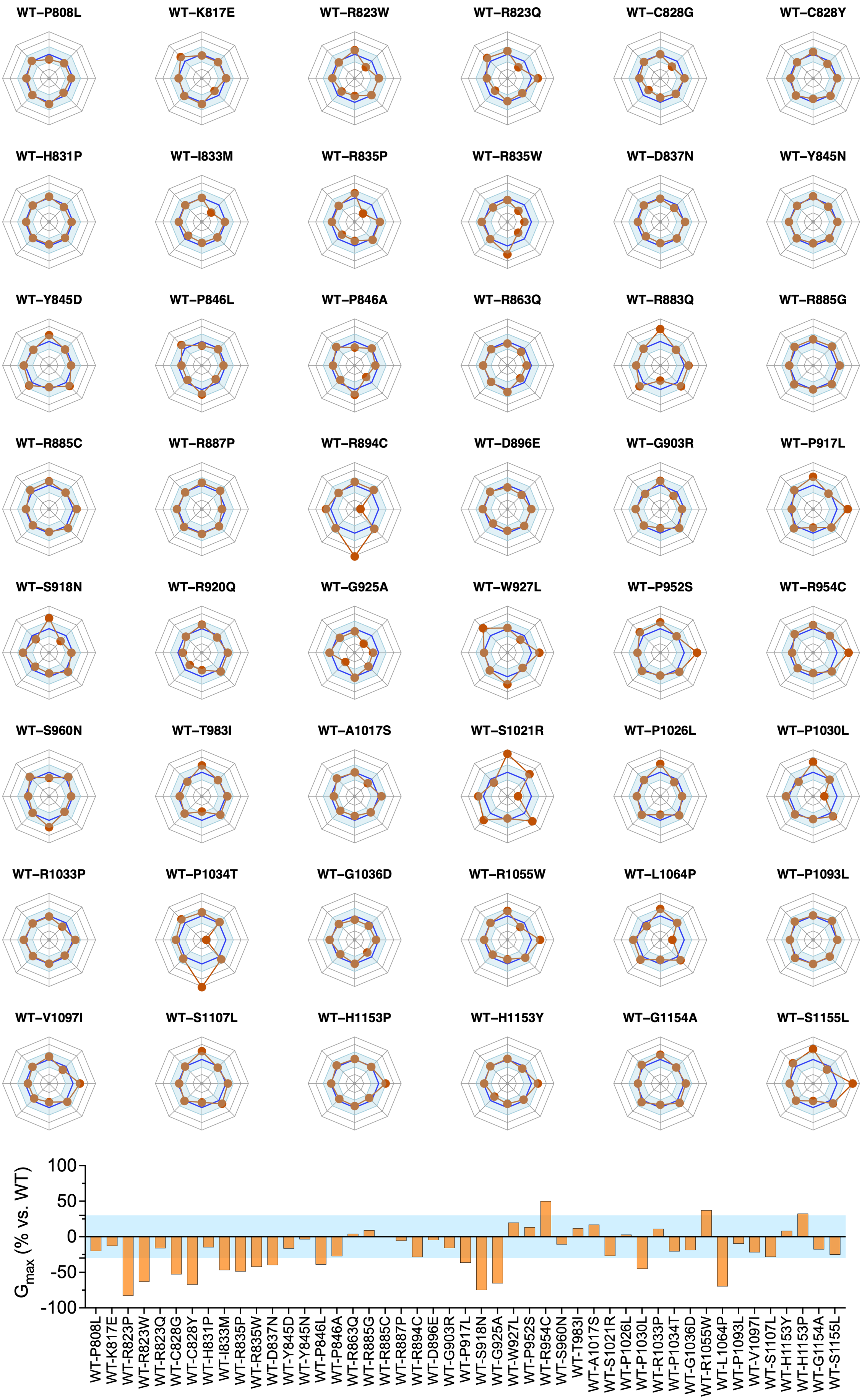
WT–P808L
WT–K817E
WT–R823W
WT–R823Q
WT–C828G
WT–C828Y
WT–H831P
WT–I833M
WT–R835P
WT–R835W
WT–D837N
WT–Y845N
WT–Y845D
WT–P846L
WT–P846A
WT–R863Q
WT–R883Q
WT–R885G
WT–R885C
WT–R887P
WT–R894C
WT–D896E
WT–G903R
WT–P917L
WT–S918N
WT–R920Q
WT–G925A
WT–W927L
WT–P952S
WT–R954C
WT–S960N
WT–T983I
WT–A1017S
WT–S1021R
WT–P1026L
WT–P1030L
WT–R1033P
WT–P1034T
WT–G1036D
WT–R1055W
WT–L1064P
WT–P1093L
WT–V1097I
WT–S1107L
WT–H1153P
WT–H1153Y
WT–G1154A
WT–S1155L
$G_{max}$ (% vs. WT)
100
50
0
-50
-100

**Figure S8. Radar plots of 192 hERG variants under heterozygous conditions.** Radar plots showing the variant-specific biophysical fingerprints of 191 hERG variants under heterozygous conditions, including only variants with sufficient current for analysis (**n ≥ 5** for each parameter). Each axis represents a normalized gating parameter, and concentric lines indicate Z-score deviations relative to wild type. The blue shaded area denotes the wild-type range (±1 Z-score), enabling direct visualization of the magnitude and direction of variant-induced changes (outward: GoF or inward: LoF) across parameters. Orange dots indicate the Z-score of each variant parameter relative to wild type. The current amplitude variation relative to wild type (Gmax, % vs. WT) is shown below radar plots, expressed as the percentage change in current amplitude for each variant.

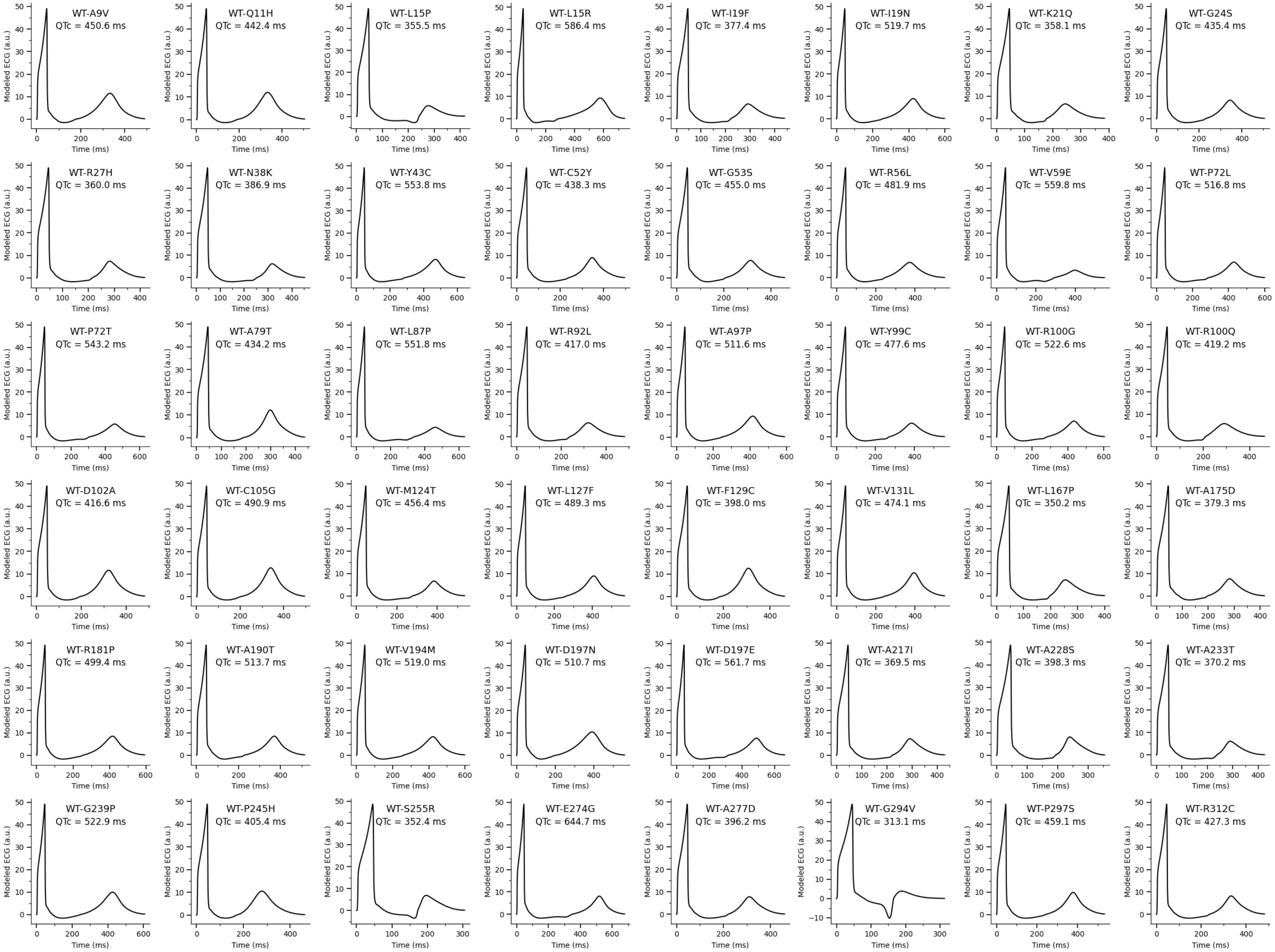
WT-A9V
QTc = 450.6 ms
WT-Q11H
QTc = 442.4 ms
WT-L15P
QTc = 355.5 ms
WT-L15R
QTc = 586.4 ms
WT-I19F
QTc = 377.4 ms
WT-I19N
QTc = 519.7 ms
WT-K21Q
QTc = 358.1 ms
WT-G24S
QTc = 435.4 ms
WT-R27H
QTc = 360.0 ms
WT-N38K
QTc = 386.9 ms
WT-Y43C
QTc = 553.8 ms
WT-C52Y
QTc = 438.3 ms
WT-G53S
QTc = 455.0 ms
WT-R56L
QTc = 481.9 ms
WT-V59E
QTc = 559.8 ms
WT-P72L
QTc = 516.8 ms
WT-P72T
QTc = 543.2 ms
WT-A79T
QTc = 434.2 ms
WT-L87P
QTc = 551.8 ms
WT-R92L
QTc = 417.0 ms
WT-A97P
QTc = 511.6 ms
WT-Y99C
QTc = 477.6 ms
WT-R100G
QTc = 522.6 ms
WT-R100Q
QTc = 419.2 ms
WT-D102A
QTc = 416.6 ms
WT-C105G
QTc = 490.9 ms
WT-M124T
QTc = 456.4 ms
WT-L127F
QTc = 489.3 ms
WT-F129C
QTc = 398.0 ms
WT-V131L
QTc = 474.1 ms
WT-L167P
QTc = 350.2 ms
WT-A175D
QTc = 379.3 ms
WT-R181P
QTc = 499.4 ms
WT-A190T
QTc = 513.7 ms
WT-V194M
QTc = 519.0 ms
WT-D197N
QTc = 510.7 ms
WT-D197E
QTc = 561.7 ms
WT-A217I
QTc = 369.5 ms
WT-A228S
QTc = 398.3 ms
WT-A233T
QTc = 370.2 ms
WT-G239P
QTc = 522.9 ms
WT-P245H
QTc = 405.4 ms
WT-S255R
QTc = 352.4 ms
WT-E274G
QTc = 644.7 ms
WT-A277D
QTc = 396.2 ms
WT-G294V
QTc = 313.1 ms
WT-P297S
QTc = 459.1 ms
WT-R312C
QTc = 427.3 ms
Modeled ECG (a.u.)
Time (ms)

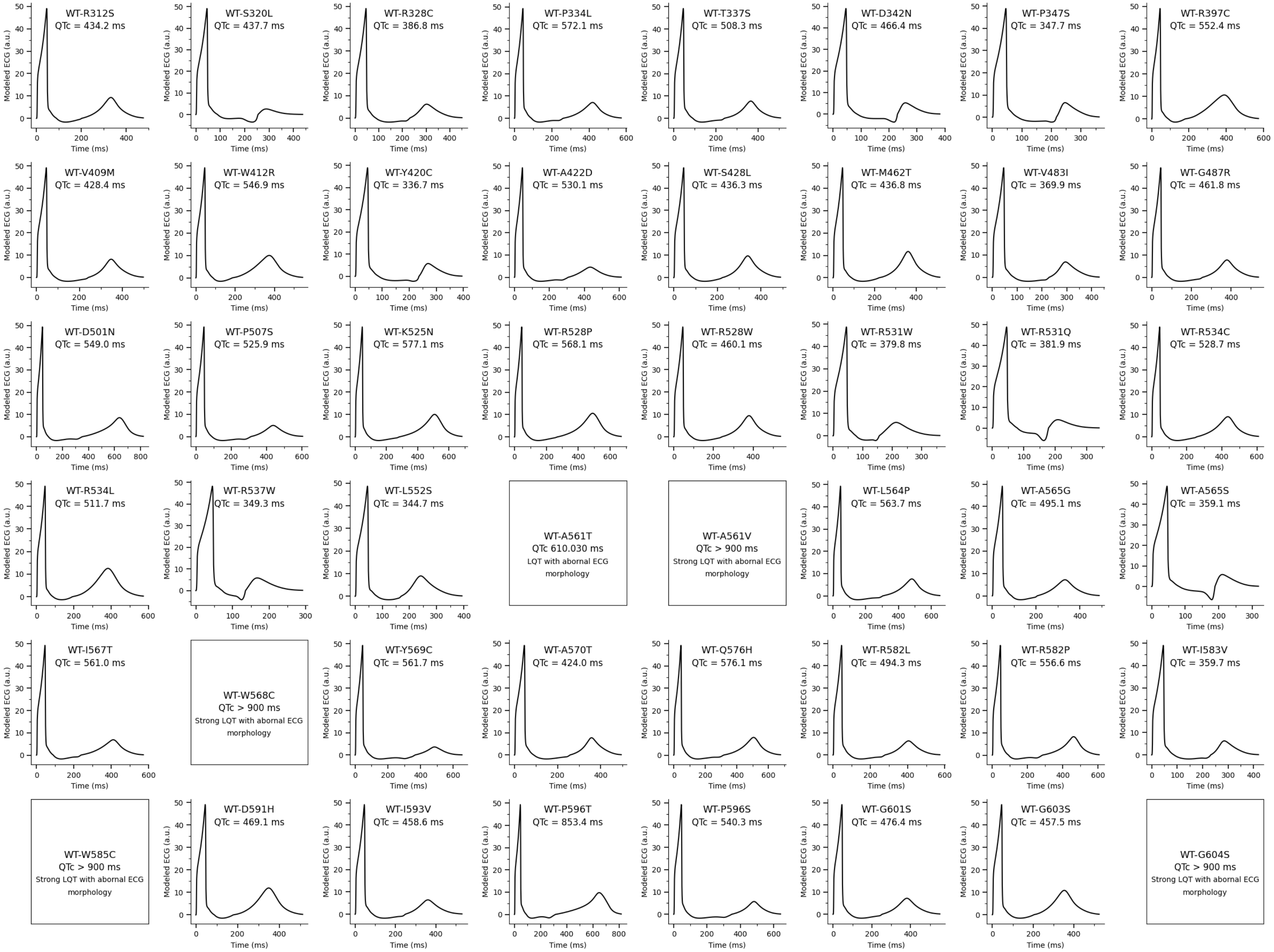
Modeled ECG (a.u.)
Time (ms)
WT-R312S QTc = 434.2 ms
WT-S320L QTc = 437.7 ms
WT-R328C QTc = 386.8 ms
WT-P334L QTc = 572.1 ms
WT-T337S QTc = 508.3 ms
WT-D342N QTc = 466.4 ms
WT-P347S QTc = 347.7 ms
WT-R397C QTc = 552.4 ms
WT-V409M QTc = 428.4 ms
WT-W412R QTc = 546.9 ms
WT-Y420C QTc = 336.7 ms
WT-A422D QTc = 530.1 ms
WT-S428L QTc = 436.3 ms
WT-M462T QTc = 436.8 ms
WT-V483I QTc = 369.9 ms
WT-G487R QTc = 461.8 ms
WT-D501N QTc = 549.0 ms
WT-P507S QTc = 525.9 ms
WT-K525N QTc = 577.1 ms
WT-R528P QTc = 568.1 ms
WT-R528W QTc = 460.1 ms
WT-R531W QTc = 379.8 ms
WT-R531Q QTc = 381.9 ms
WT-R534C QTc = 528.7 ms
WT-R534L QTc = 511.7 ms
WT-R537W QTc = 349.3 ms
WT-L552S QTc = 344.7 ms
WT-A561T QTc 610.030 ms LQT with abornal ECG morphology
WT-A561V QTc > 900 ms Strong LQT with abornal ECG morphology
WT-L564P QTc = 563.7 ms
WT-A565G QTc = 495.1 ms
WT-A565S QTc = 359.1 ms
WT-I567T QTc = 561.0 ms
WT-W568C QTc > 900 ms Strong LQT with abornal ECG morphology
WT-Y569C QTc = 561.7 ms
WT-A570T QTc = 424.0 ms
WT-Q576H QTc = 576.1 ms
WT-R582L QTc = 494.3 ms
WT-R582P QTc = 556.6 ms
WT-I583V QTc = 359.7 ms
WT-W585C QTc > 900 ms Strong LQT with abornal ECG morphology
WT-D591H QTc = 469.1 ms
WT-I593V QTc = 458.6 ms
WT-P596T QTc = 853.4 ms
WT-P596S QTc = 540.3 ms
WT-G601S QTc = 476.4 ms
WT-G603S QTc = 457.5 ms
WT-G604S QTc > 900 ms Strong LQT with abornal ECG morphology

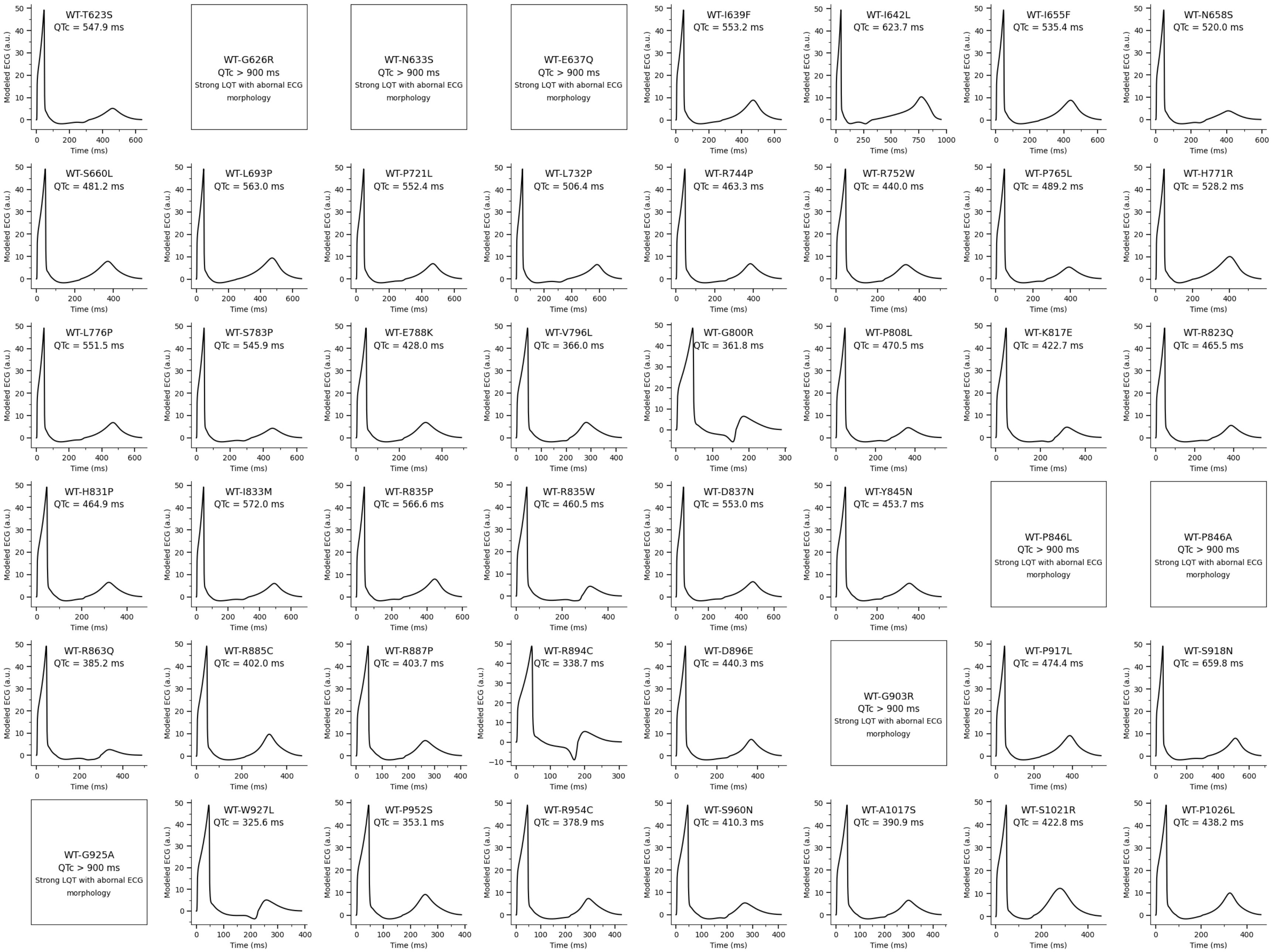
WT-T623S
QTc = 547.9 ms
WT-G626R
QTc > 900 ms
Strong LQT with abornal ECG morphology
WT-N633S
QTc > 900 ms
Strong LQT with abornal ECG morphology
WT-E637Q
QTc > 900 ms
Strong LQT with abornal ECG morphology
WT-I639F
QTc = 553.2 ms
WT-I642L
QTc = 623.7 ms
WT-I655F
QTc = 535.4 ms
WT-N658S
QTc = 520.0 ms
WT-S660L
QTc = 481.2 ms
WT-L693P
QTc = 563.0 ms
WT-P721L
QTc = 552.4 ms
WT-L732P
QTc = 506.4 ms
WT-R744P
QTc = 463.3 ms
WT-R752W
QTc = 440.0 ms
WT-P765L
QTc = 489.2 ms
WT-H771R
QTc = 528.2 ms
WT-L776P
QTc = 551.5 ms
WT-S783P
QTc = 545.9 ms
WT-E788K
QTc = 428.0 ms
WT-V796L
QTc = 366.0 ms
WT-G800R
QTc = 361.8 ms
WT-P808L
QTc = 470.5 ms
WT-K817E
QTc = 422.7 ms
WT-R823Q
QTc = 465.5 ms
WT-H831P
QTc = 464.9 ms
WT-I833M
QTc = 572.0 ms
WT-R835P
QTc = 566.6 ms
WT-R835W
QTc = 460.5 ms
WT-D837N
QTc = 553.0 ms
WT-Y845N
QTc = 453.7 ms
WT-P846L
QTc > 900 ms
Strong LQT with abornal ECG morphology
WT-P846A
QTc > 900 ms
Strong LQT with abornal ECG morphology
WT-R863Q
QTc = 385.2 ms
WT-R885C
QTc = 402.0 ms
WT-R887P
QTc = 403.7 ms
WT-R894C
QTc = 338.7 ms
WT-D896E
QTc = 440.3 ms
WT-G903R
QTc > 900 ms
Strong LQT with abornal ECG morphology
WT-P917L
QTc = 474.4 ms
WT-S918N
QTc = 659.8 ms
WT-G925A
QTc > 900 ms
Strong LQT with abornal ECG morphology
WT-W927L
QTc = 325.6 ms
WT-P952S
QTc = 353.1 ms
WT-R954C
QTc = 378.9 ms
WT-S960N
QTc = 410.3 ms
WT-A1017S
QTc = 390.9 ms
WT-S1021R
QTc = 422.8 ms
WT-P1026L
QTc = 438.2 ms
Modeled ECG (a.u.)
Time (ms)

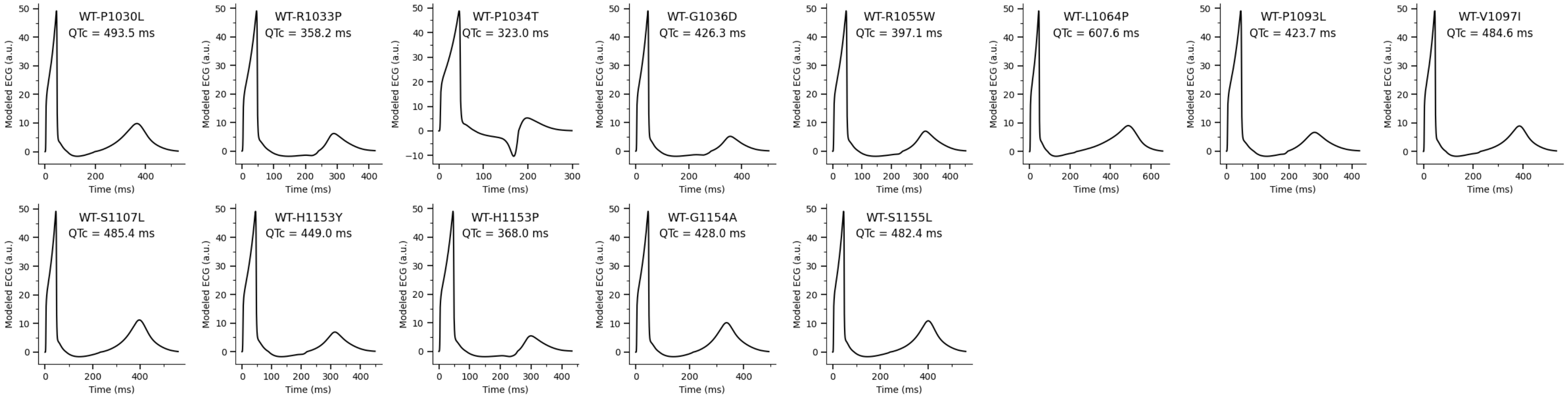


**Figure S9. ECGs modeled by MeTAL for *KCNH2* variants characterized under heterozygous conditions.** Representative ECG traces modeled by MeTAL for 157 *KCNH2* variants characterized under heterozygous conditions. For each variant, the trace corresponds to the ECG modeled from the mean values of the experimentally measured biophysical parameters in one of 1,100 virtual patients and illustrates the predicted electrophysiological phenotype. The QTc value modeled by MeTAL, calculated from the mean functional parameters across 1,100 virtual patients, is indicated for each variant. For 37 variants (19%), predominantly located in the pore region, no single ECG trace was considered representative because of marked inter-individual variability in ECG morphology across the virtual population. Despite this heterogeneity, the modeled ECGs consistently showed a prolonged-QT phenotype, in agreement with the mean MeTAL-predicted QTc.

*a*

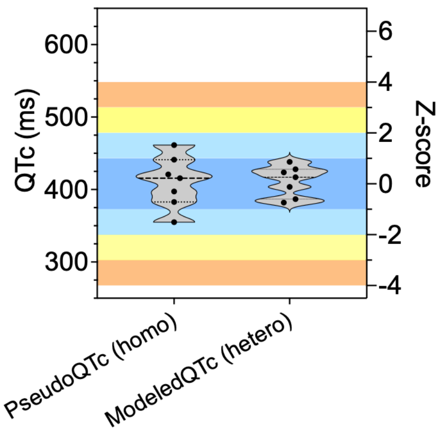


**Figure S10. Validation of control variants with normal pseudoQTc values in heterozygous condition.** Comparison of QTc values predicted from homozygous functional data (pseudoQTc) and QTc values predicted from heterozygous functional data (modeledQTc) for seven control variants selected on the basis of normal pseudoQTc values. All variants remained within the normal QTc range when evaluated using heterozygous functional data, with no significant change in the predicted QT phenotype. These results support the robustness of the MeTAL framework and demonstrate that variants predicted to have a normal QT phenotype from homozygous functional measurements remain correctly classified when assessed using heterozygous functional data.

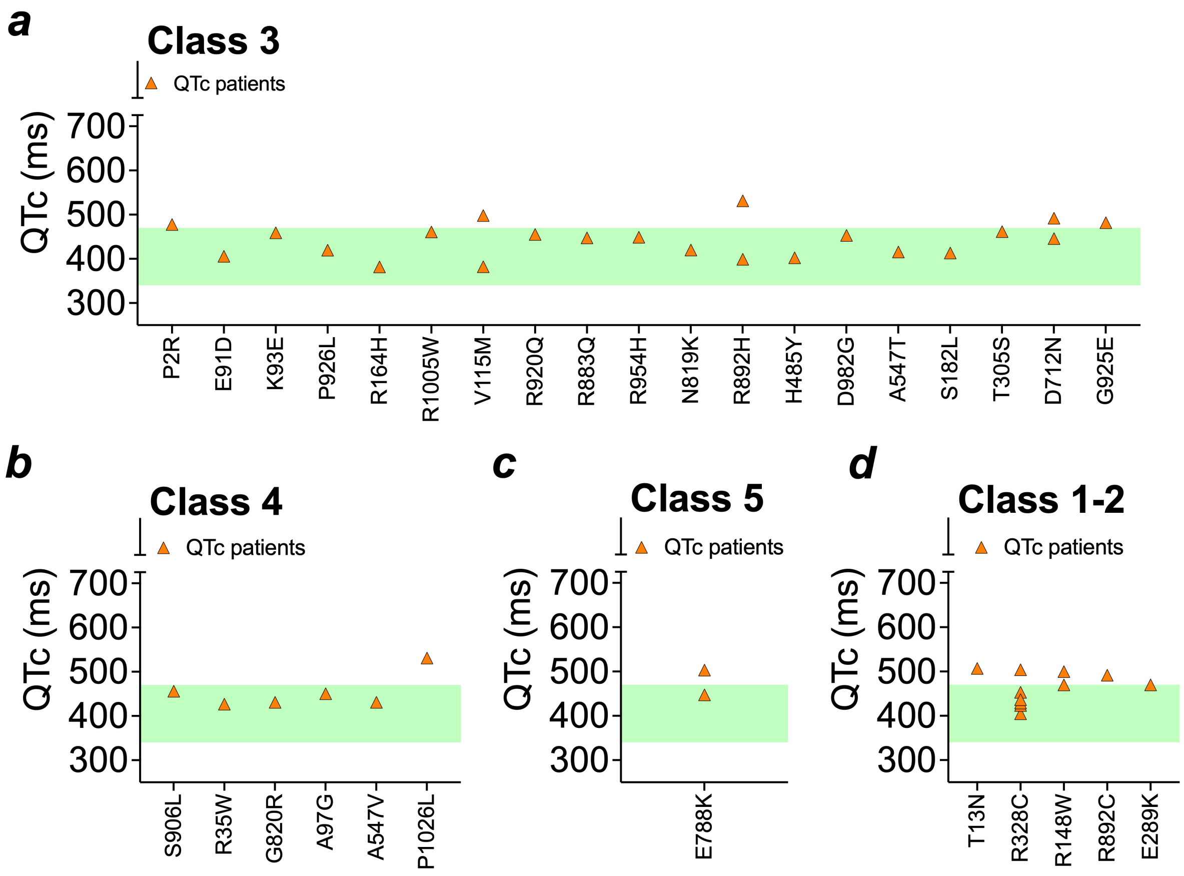


**Figure S11. Clinical validation of pseudoQTc predictions for variants with normal pseudoQTc values**. ***a-c***, Distribution of clinical QTc values for patients carrying hERG variants with normal pseudoQTc values belonging to ACMG classes 3 (***a***), 4 (***b***), and 5 (***c***). Each triangle represents one patient. The green shaded area indicates the normal QTc range (≤470 ms). The majority of patients carrying these variants exhibited normal QTc values, supporting the decision not to perform heterozygous functional modeling for variants predicted to have a normal phenotype. For variants associated with both normal and prolonged QTc values, representative patients with normal QTc were also identified (except P1026L), indicating incomplete clinical penetrance. ***d***, Clinical QTc values for ACMG class 1-2 variants with normal pseudoQTc values. Although most patients had normal QTc values, a subset presented prolonged QTc despite normal functional predictions, suggesting that factors other than the hERG variant likely contribute to the clinical phenotype.

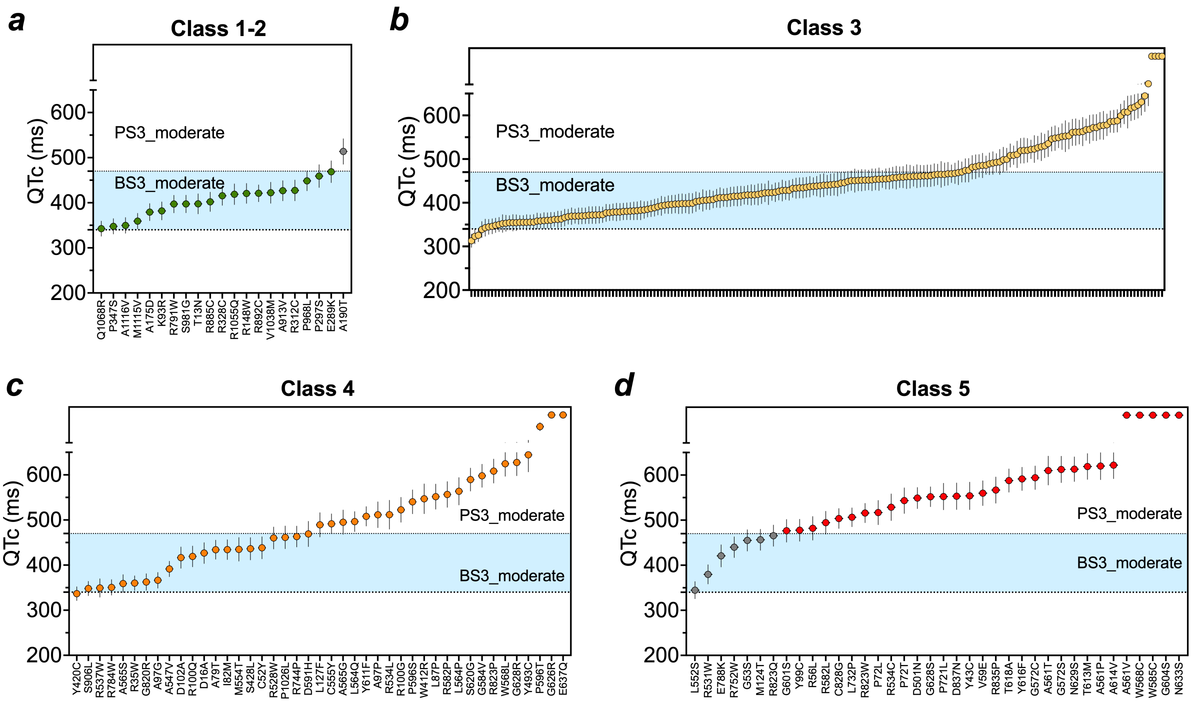


**Figure S12. Variant-level distribution of MeTAL-modeled QTc phenotypes underlying OddsPath functional evidence assignment.** Individual *KCNH2* variants are shown according to their ACMG classification (classes 1–2, 3, 4, and 5) and categorized on the basis of their MeTAL-modeled QTc phenotype as normal or abnormal. Normal modeled QTc phenotypes correspond to BS3_Moderate functional evidence, whereas abnormal phenotypes LQT correspond to PS3_Moderate functional evidence, according to the categorical OddsPath calibration described in Figure 5. Individual variant names are indicated for ACMG classes 1–2, 4, and 5; class 3 variants are shown without individual labels because of their larger number (n = 201).
